# The Therapy Intensity Level scale for traumatic brain injury: clinimetric assessment on neuro-monitored patients across 52 European intensive care units

**DOI:** 10.1101/2023.08.03.23293615

**Authors:** Shubhayu Bhattacharyay, Erta Beqiri, Patrick Zuercher, Lindsay Wilson, Ewout W Steyerberg, David W Nelson, Andrew I R Maas, David K Menon, Ari Ercole, the CENTER-TBI investigators and participants

## Abstract

Intracranial pressure (ICP) data from traumatic brain injury (TBI) patients in the intensive care unit (ICU) cannot be interpreted appropriately without accounting for the effect of administered therapy intensity level (TIL) on ICP. A 15-point scale was originally proposed in 1987 to quantify the hourly intensity of ICP-targeted treatment. This scale was subsequently modified – through expert consensus – during the development of TBI Common Data Elements to address statistical limitations and improve usability. The latest, 38-point scale (hereafter referred to as TIL) permits integrated scoring for a 24- hour period and has a five-category, condensed version (TIL^(Basic)^) based on qualitative assessment. Here, we perform a total- and component-score analysis of TIL and TIL^(Basic)^ to: (1) validate the scales across the wide variation in contemporary ICP management, (2) compare their performance against that of predecessors, and (3) derive guidelines for proper scale use. From the observational Collaborative European NeuroTrauma Effectiveness Research in TBI (CENTER-TBI) study, we extract clinical data from a prospective cohort of ICP-monitored TBI patients (*n*=873) from 52 ICUs across 19 countries. We calculate daily TIL and TIL^(Basic)^ scores (TIL_24_ and TIL^(Basic)^_24_, respectively) from each patient’s first week of ICU stay. We also calculate summary TIL and TIL^(Basic)^ scores by taking the first-week maximum (TIL_max_ and TIL^(Basic)^_max_) and first-week median (TIL_median_ and TIL^(Basic)^_median_) of TIL_24_ and TIL^(Basic)^_24_ scores for each patient. We find that, across all measures of construct and criterion validity, the latest TIL scale performs significantly greater than or similarly to all alternative scales (including TIL^(Basic)^) and integrates the widest range of modern ICP treatments. TIL_median_ outperforms both TIL_max_ and summarised ICP values in detecting refractory intracranial hypertension (RICH) during ICU stay. The RICH detection thresholds which maximise the sum of sensitivity and specificity are TIL_median_≥7.5 and TIL_max_≥14. The TIL_24_ threshold which maximises the sum of sensitivity and specificity in the detection of surgical ICP control is TIL_24_≥9. The median scores of each TIL component therapy over increasing TIL_24_ reflect a credible staircase approach to treatment intensity escalation, from head positioning to surgical ICP control, as well as considerable variability in the use of cerebrospinal fluid drainage and decompressive craniectomy. Since TIL^(Basic)^_max_ suffers from a strong statistical ceiling effect and only covers 17% (95% CI: 16–18%) of the information in TIL_max_, TIL^(Basic)^ should not be used instead of TIL for rating maximum treatment intensity. TIL^(Basic)^_24_ and TIL^(Basic)^_median_ can be suitable replacements for TIL_24_ and TIL_median_, respectively (with up to 33% [95% CI: 31–35%] information coverage) when TIL assessment is infeasible. Accordingly, we derive numerical ranges for categorising TIL_24_ scores into TIL^(Basic)^_24_ scores. In conclusion, our results validate TIL across a spectrum of ICP management and monitoring approaches. TIL is a more sensitive surrogate for pathophysiology than ICP and thus can be considered an intermediate outcome after TBI.

## Introduction

Elevated intracranial pressure (ICP) following traumatic brain injury (TBI) may impede the potential recovery of injured brain tissue and damage initially unaffected brain regions.^1^ Therefore, for TBI patients admitted to the intensive care unit (ICU), clinicians often monitor ICP and apply a wide range of ICP-reducing treatments.^2^ The selective use of these treatments typically follows a staircase approach, in which therapeutic intensity – defined by the risk and complexity of each treatment – is incrementally escalated until adequate ICP control is achieved.^3–5^ Thus, therapeutic intensity must be considered when interpreting ICP. Even if two TBI patients have comparable ICP values, a difference in the intensity of their ICP-directed therapies likely indicates a difference in pathophysiological severity.

Several versions of the Therapy Intensity Level (TIL) scale have been developed to rate and compare the overall intensity of ICP management amongst TBI patients. TIL scales assign a relative intensity score to each ICP-targeting therapy and return either the sum or the maximum value of the scores of simultaneously applied therapies. In 1987, Maset *et al.* produced the original, 15-point TIL scale (TIL^(1987)^) to be assessed once every four hours.^6^ In 2006, Shore *et al.* published the 38-point Paediatric Intensity Level of Therapy (PILOT) scale,^7^ revising TIL^(1987)^ to: (1) represent updated paediatric TBI management practices, (2) have a more practical, daily assessment frequency, and (3) resolve a statistical ceiling effect. In 2011, the interagency TBI Common Data Elements (CDE) scheme developed the most recent, 38-point TIL scale (hereafter referred to as TIL) as well as a condensed, five-category TIL^(Basic)^ scale through expert consensus.^8^ The TIL scale revised PILOT to integrate additional ICP-directed therapies and to be applicable to adult TBI management. Moreover, TIL^(Basic)^ was proposed as a simple, categorical measure to use when full TIL assessment would be infeasible. Since Zuercher *et al.* reported the validity and reliability of TIL in a two-centre cohort (*n*=31) in 2016,^9^ the scale has become a popular research metric for quantifying ICP treatment intensity.^10–13^

However, several critical questions regarding TIL remain unanswered. It is uncertain whether the validity of TIL, reported in a relatively small population, can be generalised across the wide variation of ICP management, monitoring, and data acquisition (i.e., intermittent chart recording or high-resolution storage^14^) strategies practised in contemporary intensive care.^11,12,15,16^ Furthermore, the scoring configuration of TIL has never been tested against alternatives (e.g., TIL^(1987)^ and PILOT), and the relative contribution of TIL’s component therapies towards the total score is unknown. It is unclear how TIL^(Basic)^ numerically relates to TIL and if the former captures the essential information of the latter. In this work, we aimed to answer these questions by performing a comprehensive assessment of TIL on a large, contemporary population of ICP-monitored TBI patients across European ICUs.

## Materials and Methods

### Therapy intensity level (TIL) and alternative scales

TIL refers to the 38-point scale developed by the CDE scheme for TBI.^8^ The domain or construct (i.e., targeted concept of a scale) of TIL is the therapeutic intensity of ICP management. The TIL scale has twelve items, each representing a distinct ICP-targeting treatment from one of eight modalities, as defined in Table 1. TIL was developed by an international expert panel which discussed: (1) the relevant ICP-treatment modalities of modern intensive care, (2) the relative risk and efficacy of individual therapies to derive scores, and (3) practical and statistical limitations of previous TIL scores.^8^ In this way, TIL is a formative measurement model, in which the construct (i.e., ICP treatment intensity) is not unidimensional but rather defined by the combination of items (i.e., ICP-targeting treatments).^17^ TIL was shown to have high interrater and intrarater reliability by Zuercher *et al*.^9^ If a decompressive craniectomy was performed as a last resort for refractory intracranial hypertension, its score was included in the day of the operation and in every subsequent day of ICU stay. TIL scores can be calculated as frequently as clinically desired. For our analysis, we calculated the following TIL scores from the first seven days of ICU stay:

- TIL_24_, the daily TIL score based on the sum of the highest scores per item per calendar day,
- TIL_max_, the maximum TIL_24_ over the first week of a patient’s ICU stay,
- TIL_median_, the median TIL_24_ over the first week of a patient’s ICU stay.

**Table 1.**
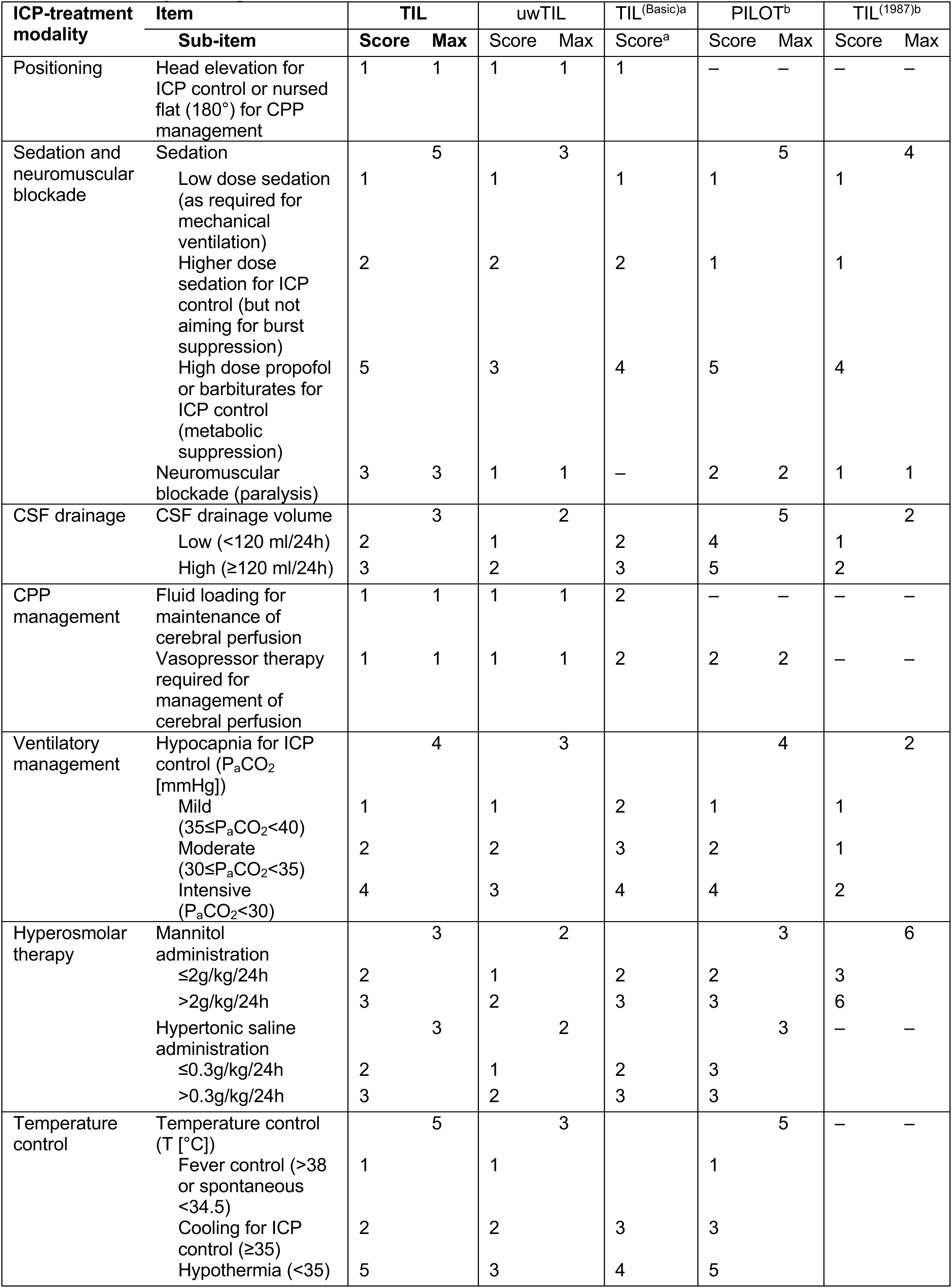
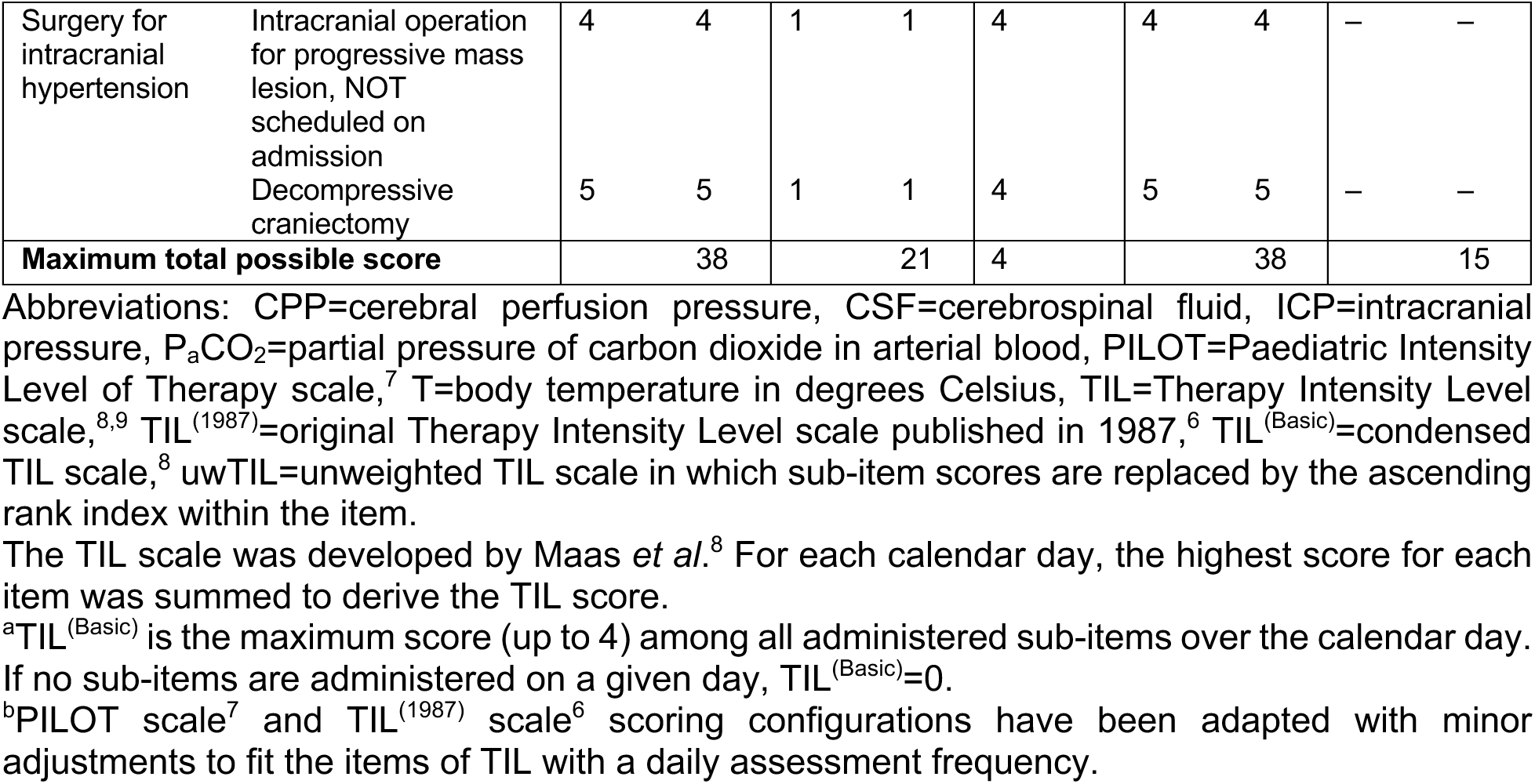
Scoring configurations for TIL and alternative scales.

We also calculated scores from four other therapeutic intensity scales to compare with TIL scores. The 21-point, unweighted TIL (uwTIL) scale replaces each sub-item score in TIL with its ascending rank index (i.e., 1, 2, 3, …) within each item (Table 1). The five- category TIL^(Basic)^ was also developed by the CDE scheme for TBI and takes the maximum score, from zero (i.e., no ICP-related intervention) to four, amongst all included sub-items over the calendar day.^8^ We adapted the 38-point PILOT^7^ and 15-point TIL^(1987)^ scales^6^ with minor adjustments to fit the items of TIL with a daily assessment frequency. PILOT was also shown to have high interrater and intrarater reliability by Shore *et al*.^7^ For the four alternative scales, daily (i.e., uwTIL_24_, TIL^(Basic)^_24_, PILOT_24_, and TIL^(1987)^_24_), maximum (i.e., uwTIL_max_, TIL^(Basic)^_max_, PILOT_max_, and TIL^(1987)^_max_), and median (i.e., uwTIL_median_, TIL^(Basic)^_median_, PILOT_median_, and TIL^(1987)^_median_) scores were calculated in the same way as TIL_24_, TIL_max_, and TIL_median_, respectively.

### Study design and populations

Our study population was prospectively recruited for the Collaborative European NeuroTrauma Effectiveness Research in Traumatic Brain Injury (CENTER-TBI) core and high-resolution studies. CENTER-TBI is a longitudinal, observational cohort study (NCT02210221) involving 65 medical centres across 18 European countries and Israel. Patients were recruited between 19 December 2014 and 17 December 2017 if they met the following criteria: (1) presentation within 24 hours of a TBI, (2) clinical indication for a CT scan, and (3) no severe pre-existing neurological disorder. In accordance with relevant laws of the European Union and the local country, ethical approval was obtained for each site, and written informed consent by the patient or legal representative was documented electronically. The list of sites, ethical committees, approval numbers, and approval dates can be found online: https://www.center-tbi.eu/project/ethical-approval. The project objectives and design of CENTER-TBI have been described in detail previously.^18,19^

In this work, we applied the following inclusion criteria in addition to those of CENTER-TBI (Fig. 1): (1) primary admission to the ICU, (2) at least 16 years old at ICU admission, (3) invasive ICP monitoring, (4) no decision to withdraw life-sustaining therapies (WLST) on the first day of ICU stay, and (5) daily assessment of TIL.

**Fig. 1.**
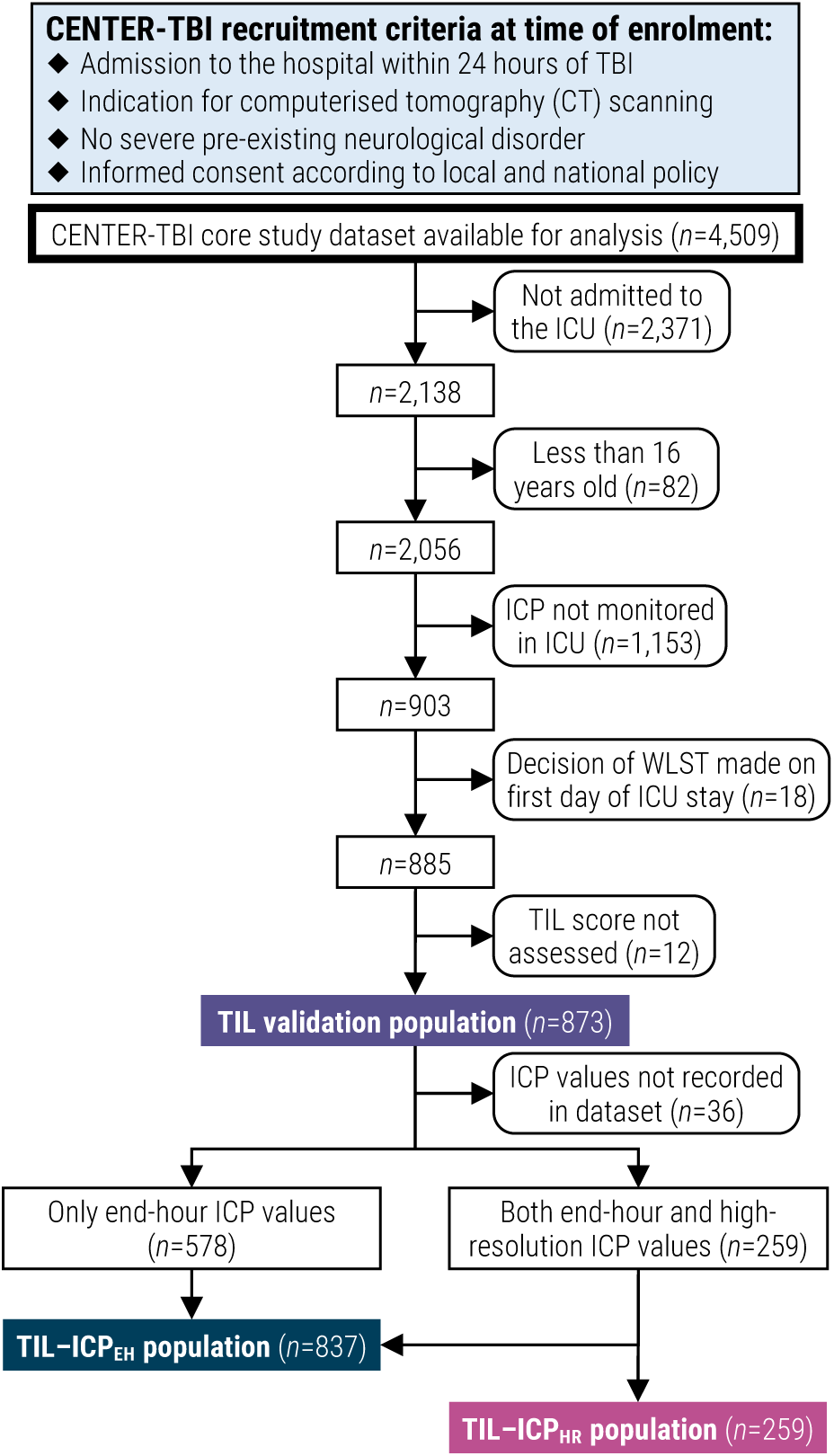
Flow diagram for patient enrolment and validation population assignment. Abbreviations: CENTER-TBI=Collaborative European NeuroTrauma Effectiveness Research in TBI, ICP=intracranial pressure, ICPEH=end-hour ICP, ICPHR=high-resolution ICP, ICU=intensive care unit, TBI=traumatic brain injury, TIL=Therapy Intensity Level scale,^8,9^ WLST=withdrawal of life-sustaining therapies.

For our sub-studies evaluating the association between TIL and ICP-derived values, we created two sub-populations based on the type of ICP values available. Patients with end- hour ICP (ICP_EH_) values, which were recorded by clinicians at the end of every other hour, constituted the TIL-ICP_EH_ sub-population. Patients with high-resolution ICP values (ICP_HR_), which were automatically stored with monitoring software, constituted the TIL- ICP_HR_ sub-population. All patients in the TIL-ICP_HR_ sub-population were also members of the TIL-ICP_EH_ sub-population (Fig. 1).

### Data collection

Data for the CENTER-TBI study was collected through the QuesGen electronic case report form system (QuesGen Systems Inc, Burlingame, CA, USA) hosted on the International Neuroinformatics Coordinating Facility (INCF) platform (INCF, Stockholm, Sweden). All data for the validation populations, except high-resolution signals, were extracted from the CENTER-TBI core study^19^ (v3.0, ICU stratum) using Opal database software.^20^

#### ICP management data for TIL calculation

Since TIL_24_ was found to be a reliable summary of hourly TIL,^9^ clinical data pertinent to the component items of TIL (i.e., ICP-guided treatments, Table 1) were recorded daily through the first week of ICU stay. We extracted all daily TIL item values for our population, and calculated TIL_24_, uwTIL_24_, TIL^(Basic)^_24_, PILOT_24_, and TIL^(1987)^_24_ as defined in Table 1. For patients who underwent WLST after the first day of ICU stay, we only extracted TIL item information from before the documented date of WLST decision.

#### ICP_EH_ and related values

End-hour ICP (ICP_EH_), systolic blood pressure (SBP_EH_), and diastolic blood pressure (DBP_EH_) were recorded by clinicians every two hours for the TIL-ICP_EH_ sub-population. Mean arterial pressure (MAP_EH_) was calculated as MAP_EH_ = (SBP_EH_ + 2DBP_EH_)/3, and cerebral perfusion pressure (CPP_EH_) was calculated as CPP_EH_ = MAP_EH_ – ICP_EH_. From ICP_EH_ and CPP_EH_, we calculated the following values:

- ICP_24_ or CPP_24_, the mean ICP or CPP value over a calendar day of ICU stay,
- ICP_max_ or CPP_min_, the maximum ICP_24_ or minimum CPP_24_ value over the first week of a patient’s ICU stay,
- ICP_median_ or CPP_median_, the median ICP_24_ or CPP_24_ value over the first week of a patient’s ICU stay.

#### ICP_HR_ and related values

High-resolution signals were collected using either ICM+ software (Cambridge Enterprise Ltd, Cambridge, UK; http://icmplus.neurosurg.cam.ac.uk), Moberg CNS monitor (Moberg Research Inc, Ambler, PA, USA; https://www.moberg.com), or both. Blood pressure was obtained through arterial lines connected to pressure transducers. High-resolution ICP (ICP_HR_) was acquired from either an intraparenchymal strain gauge probe (Codman ICP MicroSensor, Codman & Shurtleff Inc, Raynham, MA, USA), a parenchymal fibre optic pressure sensor (Camino ICP Monitor, Integra Life Sciences, Plainsboro, NJ, USA; https://www.integralife.com/), or an external ventricular drain. Detailed data collection and pre-processing methods (i.e., artefact cleaning and down-sampling to ten-second averaged time series) applied to high resolution signals in our study have been described previously.^21^ Ten-second averaged ICP (ICP_HR_10sec_) and CPP (CPP_HR_10sec_) time-series were retrieved for this analysis, and, from ICP_HR_10sec_ and CPP_HR_10s_, we calculated ICP_24_/CPP_24_, ICP_max_/CPP_min_, and ICP_median_/CPP_median_ as described above.

#### Physician impressions

Attending ICU physicians were asked to record their daily concerns with the patient’s ICP and CPP, separately, on a scale from one (not concerned) to ten (most concerned). Moreover, on each patient’s ICU discharge summary, physicians were asked to record whether the patient experienced refractory intracranial hypertension during his or her ICU stay. Refractory intracranial hypertension was defined as recurrent, sustained (i.e., of at least ten minutes) increases of ICP above 20 mmHg despite medical ICP management. We extracted the daily ICP/CPP concern ratings and refractory intracranial hypertension impressions which coincided with the ICU stays of our population.

#### Baseline characteristics, prognosis, and outcome

We extracted baseline demographic characteristics, Marshall CT classifications,^22^ and Glasgow Coma Scale (GCS)^23^ scores from ICU admission.^24^ We also extracted Glasgow Outcome Scale – Extended (GOSE) functional outcome scores at six months post-injury,^25^ with imputation of missing values as previously described.^26^ Finally, we extracted ordinal functional outcome prognosis scores, calculated from a tokenised embedding of all available clinical information in the first 24 hours of ICU stay, as described previously.^27^

### Validation

We appraised the validity of TIL according to recommendations of best practice from clinimetric literature.^28^ Based on the identified domain of TIL, we evaluated the construct and criterion validities of TIL. Our qualitative and quantitative assessments of TIL were performed against those of alternative scoring configurations (Table 1) for comparison.

#### Construct validity

Construct validity is the extent to which a clinical scale matches expectations of associations with parameters within or outside the identified domain. Construct validity is further broken down into convergent validity (i.e., associations with similar constructs), discriminant validity (i.e., associations with divergent constructs), and differentiation by known groups. In this work, statistical associations between study variables were measured with:

- Spearman’s correlation coefficients (*ρ*) for static (i.e., measured once) variables,
- repeated measures correlation coefficients (*r_rm_*)^29^ – interpreted as within-individual strength of association – for longitudinal (i.e., measured over time) variables,
- linear mixed effects regression (LMER) coefficients (*β_LMER_*) of daily scale scores (e.g., TIL_24_) when regressing ICP_24_ or CPP_24_ on daily scale scores and the day of ICU stay (Day_ICU_), accounting for inter-patient variability with random intercepts. Therefore, *β_LMER_* were interpreted as the expected difference in ICP_24_ or CPP_24_ per unit increase of daily scale score, independent of time since ICU admission or inter-patient variation.

For convergent validity, we expected therapeutic intensity to correlate at least mildly (i.e., |*ρ*|≥0.2, |*r_rm_*|≥0.2, |*β_LMER_*|>0) with markers of injury severity (i.e., baseline GCS and baseline outcome prognoses), functional outcome (i.e., six-month GOSE), clinical concerns of ICP status, and ICP itself. Accordingly, we calculated: (1) *ρ* values between TIL_max_ and GCS, ordinal prognosis scores, GOSE, and ICP_max_, (2) *ρ* values between TIL_median_ and GCS, ordinal prognosis scores, GOSE, and ICP_median_, (3) *r_rm_* values between TIL_24_ and physician concern of ICP and ICP_24_, and (4) *β_LMER_* of TIL_24_ when regressing ICP_24_ on Day_ICU_ and TIL_24_ (i.e., ICP_24_∼Day_ICU_+TIL_24_), accounting for inter-patient variability with random intercepts.

For discriminant validity, we expected therapeutic intensity to be more strongly correlated with ICP and physician concerns of ICP than with CPP and physician concerns of CPP, respectively. Even though CPP control through fluid loading or vasopressor therapy is a component modality of TIL (Table 1), we expected TIL to capture ICP management (i.e., the construct) more accurately than CPP management. We compared: (1) *ρ* values of TIL_max_ vs. CPP_min_ to those of TIL_max_ vs. ICP_max_, (2) *ρ* values of TIL_median_ vs. CPP_median_ to those of TIL_median_ vs. ICP_median_, (3) *r_rm_* values of TIL_24_ vs. CPP_24_ to those of TIL_24_ vs. ICP_24_, and (4) the *β_LMER_* of TIL_24_ when regressing CPP_24_∼Day_ICU_+TIL_24_ to the *β_LMER_* of TIL_24_ when regressing ICP_24_∼Day_ICU_+TIL_24_.

For differentiation by known groups, we expected TIL_max_ and TIL_median_ to effectively discriminate patients who experienced refractory intracranial hypertension during ICU stay from those who did not. We calculated the area under the receiver operating characteristic curve (AUC), which, in our case, was interpreted as the probability of a randomly selected patient with refractory intracranial hypertension having a higher TIL_max_ or TIL_median_ score than one without it. We also compared the AUCs of TIL_max_ and TIL_median_ to ICP_max_ and ICP_median_ and determined the sensitivity and specificity of refractory intracranial hypertension detection at each threshold of TIL_max_ and TIL_median_.

#### Criterion validity

Criterion (or concurrent) validity is the degree to which there is an association between a clinical scale and other scales measuring the same construct, particularly a gold standard assessment. Since there is no extant “gold standard” for assessing ICP management intensity, we tested the concurrent criterion validity of TIL by calculating its associations with its predecessors (i.e., PILOT and TIL^(1987)^), mindful of their limitations as described above. More specifically, we calculated: (1) *ρ* values between TIL_max_ and prior scale maximum scores (i.e., PILOT_max_ and TIL^(1987)^_max_), (2) *ρ* values between TIL_median_ and prior scale median scores (i.e., PILOT_median_ and TIL^(1987)^_median_), and (3) *r_rm_* between TIL_24_ and prior scale daily scores (i.e., PILOT_24_ and TIL^(1987)^_24_).

### Component item analysis

We evaluated inter-item (i.e., inter-treatment) and adjusted item-total associations of TIL_24_, uwTIL_24_, PILOT_24_, and TIL^(1987)^_24_ by calculating *r_rm_* values. Item-total correlations were adjusted by subtracting the tested item score from the total score prior to calculating the correlation. We measured Cronbach’s alpha (*α*) to assess internal reliability amongst scale items at each day of ICU stay. Moreover, we calculated the median score contribution of each item per total TIL_24_ score. The association between each TIL_24_ item score and ICP_24_, CPP_24_, physician concern of ICP, and physician concern of CPP was calculated with *r_rm_* values. Finally, we trained LMER models regressing ICP_24_ and CPP_24_ on all TIL items (with categorical dummy encoding) and Day_ICU_ concurrently. The *β_LMER_* values from these models were interpreted as the average change in ICP_24_ or CPP_24_ associated with each treatment when accounting for all other ICP-guided treatments, time since ICU admission, and inter-patient variability with random intercepts.

### TIL^(Basic)^ information coverage

We examined the distributions of TIL^(Basic)^_24_ per TIL_24_ and TIL_24_ per TIL^(Basic)^_24_ to derive thresholds for categorising TIL_24_ into TIL^(Basic)^_24_. We also calculated the information coverage (IC) of TIL^(Basic)^ by dividing the mutual information (MI) of TIL^(Basic)^ and TIL by the entropy of TIL. IC was calculated with TIL^(Basic)^_24_ and TIL_24_ for days one through seven of ICU stay, with TIL^(Basic)^_max_ and TIL_max_, and with TIL^(Basic)^_median_ and TIL_median_.

### Statistical analysis

#### Multiple imputation of missing values

Five of the static study variables had missing values for some of the patients in our study: GCS, GOSE, baseline prognosis scores, Marshall CT classifications, and refractory intracranial hypertension status. We assessed the patterns of missingness (Supplementary Fig. S1) and multiply imputed (*m*=100) these variables with independent, stochastic predictive mean matching functions using the *mice* package^30^ (v3.9.0) in R (v4.2.3). We assumed these variables to be missing-at-random (MAR) (as previously reported on CENTER-TBI data^31^) and supported this assumption by training imputation models on all study measures as well as correlated auxiliary variables (e.g., raised ICP during ICU stay).

For daily longitudinal study variables, we considered a value to be missing if the patient was still in the ICU and WLST had not been decided on or before that day. We assessed the longitudinal patterns of missingness (Supplementary Fig. S2) and multiply imputed (*m*=100) these variables with the multivariate, time-series algorithm from the *Amelia II* package^32^ (v1.7.6) in R over the first week of ICU stay. The algorithm exploits both between-variable and within-variable correlation structures over time to stochastically impute missing time series values in independently trained runs. We validated the MAR assumption by identifying characteristics significantly associated with longitudinal variable missingness (Supplementary Table S1) and included auxiliary information associated with value missingness (e.g., reasons for stopping ICP monitoring) in the imputation model.

#### Statistical inference

We calculated 95% confidence intervals (CI) for *ρ*, *r_rm_*, *β_LMER_*, AUC, sensitivity, specificity, *α*, and IC values using bootstrapping with 1,000 resamples of unique patients. For each resample, one of the 100 missing value imputations was randomly chosen. Therefore, confidence intervals represented the uncertainty due to patient resampling and missing value imputation.

#### Code

All statistical analyses were performed in Python (v3.8.2) or R, and all visualisations were created in R. All scripts used in this study are publicly available on GitHub: https://github.com/sbhattacharyay/CENTER-TBI_TIL.

## Results

### Study population

Of the 4,509 patients available for analysis in the CENTER-TBI core study, 873 patients from 52 ICUs met the additional inclusion criteria of this work. Amongst them, 837 constituted the TIL-ICP_EH_ sub-population and 259 constituted the TIL-ICP_HR_ sub- population (Fig. 1). Summary characteristics of the overall population as well as those of the TIL-ICP_EH_ and TIL-ICP_HR_ sub-populations are detailed in Table 2. Apart from two of the prognosis scores pertaining to the probability of returning to pre-injury life roles (i.e., Pr(GOSE>5) and Pr(GOSE>6)), none of the tested characteristics were significantly different between patients in the TIL-ICP_HR_ sub-population and those outside of it (Table 2).

**Table 2.**
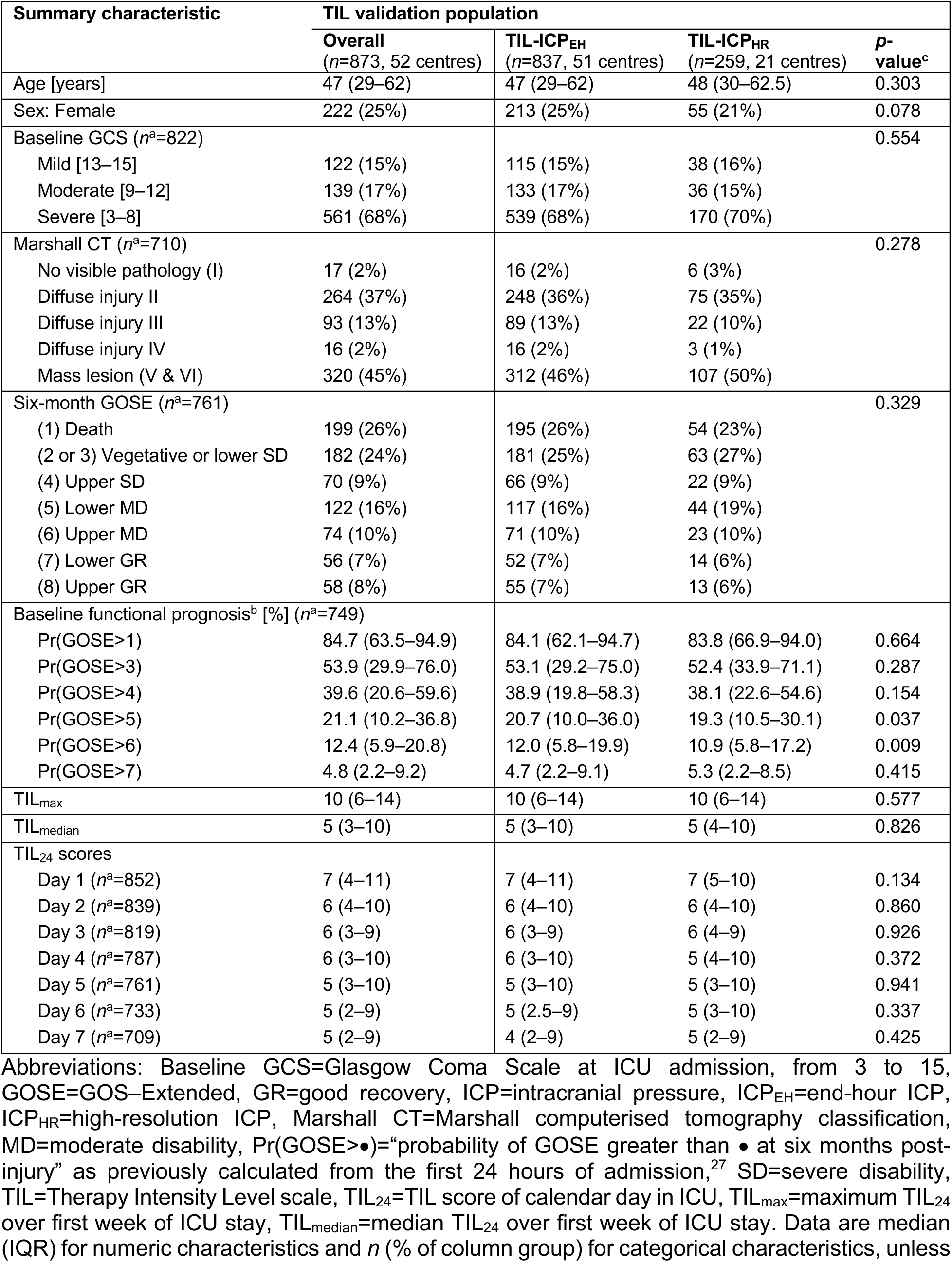
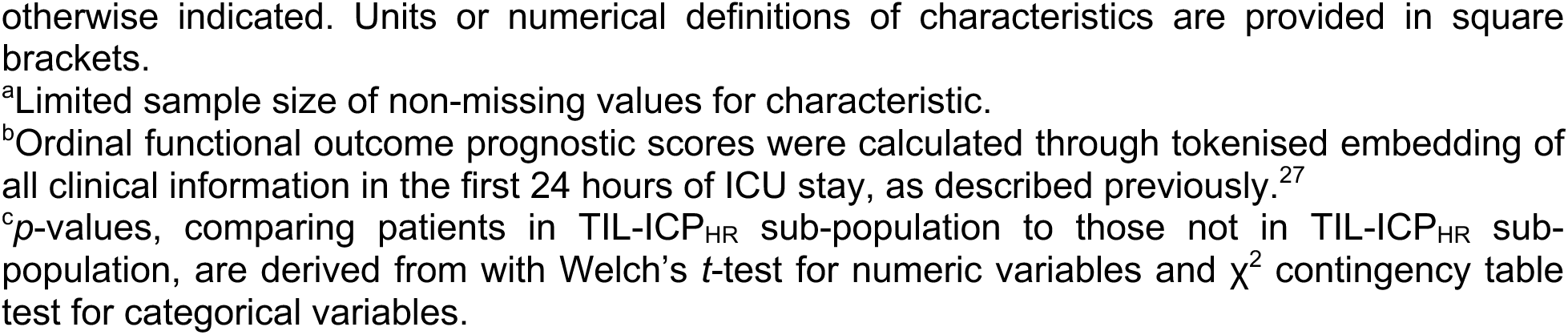
Summary characteristics of study validation populations.

| Summary characteristic | TIL validation population |  |  |  |
| --- | --- | --- | --- | --- |
|  | Overall<br>(*n*=873, 52 centres) | TIL-ICP<sub>EH</sub><br>(*n*=837, 51 centres) | TIL-ICP<sub>HR</sub><br>(*n*=259, 21 centres) | *p*-value<sup>c</sup> |
| Age [years] | 47 (29–62) | 47 (29–62) | 48 (30–62.5) | 0.303 |
| Sex: Female | 222 (25%) | 213 (25%) | 55 (21%) | 0.078 |
| Baseline GCS (*n*<sup>a</sup>=822) |  |  |  | 0.554 |
| Mild [13–15] | 122 (15%) | 115 (15%) | 38 (16%) |  |
| Moderate [9–12] | 139 (17%) | 133 (17%) | 36 (15%) |  |
| Severe [3–8] | 561 (68%) | 539 (68%) | 170 (70%) |  |
| Marshall CT (*n*<sup>a</sup>=710) |  |  |  | 0.278 |
| No visible pathology (I) | 17 (2%) | 16 (2%) | 6 (3%) |  |
| Diffuse injury II | 264 (37%) | 248 (36%) | 75 (35%) |  |
| Diffuse injury III | 93 (13%) | 89 (13%) | 22 (10%) |  |
| Diffuse injury IV | 16 (2%) | 16 (2%) | 3 (1%) |  |
| Mass lesion (V & VI) | 320 (45%) | 312 (46%) | 107 (50%) |  |
| Six-month GOSE (*n*<sup>a</sup>=761) |  |  |  | 0.329 |
| (1) Death | 199 (26%) | 195 (26%) | 54 (23%) |  |
| (2 or 3) Vegetative or lower SD | 182 (24%) | 181 (25%) | 63 (27%) |  |
| (4) Upper SD | 70 (9%) | 66 (9%) | 22 (9%) |  |
| (5) Lower MD | 122 (16%) | 117 (16%) | 44 (19%) |  |
| (6) Upper MD | 74 (10%) | 71 (10%) | 23 (10%) |  |
| (7) Lower GR | 56 (7%) | 52 (7%) | 14 (6%) |  |
| (8) Upper GR | 58 (8%) | 55 (7%) | 13 (6%) |  |
| Baseline functional prognosis<sup>b</sup> [%] (*n*<sup>a</sup>=749) |  |  |  |  |
| Pr(GOSE>1) | 84.7 (63.5–94.9) | 84.1 (62.1–94.7) | 83.8 (66.9–94.0) | 0.664 |
| Pr(GOSE>3) | 53.9 (29.9–76.0) | 53.1 (29.2–75.0) | 52.4 (33.9–71.1) | 0.287 |
| Pr(GOSE>4) | 39.6 (20.6–59.6) | 38.9 (19.8–58.3) | 38.1 (22.6–54.6) | 0.154 |
| Pr(GOSE>5) | 21.1 (10.2–36.8) | 20.7 (10.0–36.0) | 19.3 (10.5–30.1) | 0.037 |
| Pr(GOSE>6) | 12.4 (5.9–20.8) | 12.0 (5.8–19.9) | 10.9 (5.8–17.2) | 0.009 |
| Pr(GOSE>7) | 4.8 (2.2–9.2) | 4.7 (2.2–9.1) | 5.3 (2.2–8.5) | 0.415 |
| TIL<sub>max</sub> | 10 (6–14) | 10 (6–14) | 10 (6–14) | 0.577 |
| TIL<sub>median</sub> | 5 (3–10) | 5 (3–10) | 5 (4–10) | 0.826 |
| TIL<sub>24</sub> scores |  |  |  |  |
| Day 1 (*n*<sup>a</sup>=852) | 7 (4–11) | 7 (4–11) | 7 (5–10) | 0.134 |
| Day 2 (*n*<sup>a</sup>=839) | 6 (4–10) | 6 (4–10) | 6 (4–10) | 0.860 |
| Day 3 (*n*<sup>a</sup>=819) | 6 (3–9) | 6 (3–9) | 6 (4–9) | 0.926 |
| Day 4 (*n*<sup>a</sup>=787) | 6 (3–10) | 6 (3–10) | 5 (4–10) | 0.372 |
| Day 5 (*n*<sup>a</sup>=761) | 5 (3–10) | 5 (3–10) | 5 (3–10) | 0.941 |
| Day 6 (*n*<sup>a</sup>=733) | 5 (2–9) | 5 (2.5–9) | 5 (3–10) | 0.337 |
| Day 7 (*n*<sup>a</sup>=709) | 5 (2–9) | 4 (2–9) | 5 (2–9) | 0.425 |

Abbreviations: Baseline GCS=Glasgow Coma Scale at ICU admission, from 3 to 15, GOSE=GOS–Extended, GR=good recovery, ICP=intracranial pressure, ICP<sub>EH</sub>=end-hour ICP, ICP<sub>HR</sub>=high-resolution ICP, Marshall CT=Marshall computerised tomography classification, MD=moderate disability, Pr(GOSE>•)=“probability of GOSE greater than • at six months post-injury” as previously calculated from the first 24 hours of admission,<sup>27</sup> SD=severe disability, TIL=Therapy Intensity Level scale, TIL<sub>24</sub>=TIL score of calendar day in ICU, TIL<sub>max</sub>=maximum TIL<sub>24</sub> over first week of ICU stay, TIL<sub>median</sub>=median TIL<sub>24</sub> over first week of ICU stay. Data are median (IQR) for numeric characteristics and *n* (% of column group) for categorical characteristics, unless
otherwise indicated. Units or numerical definitions of characteristics are provided in square brackets.
<sup>a</sup>Limited sample size of non-missing values for characteristic.
<sup>b</sup>Ordinal functional outcome prognostic scores were calculated through tokenised embedding of all clinical information in the first 24 hours of ICU stay, as described previously.<sup>27</sup>
<sup>c</sup>*p*-values, comparing patients in TIL-ICP<sub>HR</sub> sub-population to those not in TIL-ICP<sub>HR</sub> sub-population, are derived from with Welch's *t*-test for numeric variables and $\chi^2$ contingency table test for categorical variables.

The median ICU stay duration of our population was 14 days (IQR: 7.8–23 days), and 83% (*n*=726) stayed through at least seven calendar days. At each day of ICU stay, less than 2.4% of the expected TIL scores were missing (Supplementary Fig. S2). Each TIL component item (Table 1) is represented by at least 17% (*n*=147, intracranial surgery) and each sub-item is represented by at least 4.9% (*n*=43, high-dose mannitol) of the population (Supplementary Table S2). The distributions of TIL_max_, TIL_median_, and TIL_24_, juxtaposed against the scores of alternative scales (Table 1), are displayed in Fig. 2. The distributions of TIL and PILOT were visually similar, and TIL^(Basic)^_max_ had a strong ceiling effect (i.e., 57% of the population had the maximum score). Whilst there was no significant difference in TIL_24_ distribution over the first seven days, most patients had their highest TIL_24_ (i.e., TIL_max_) soon after ICU admission (median: day two, IQR: days one–three). The Spearman’s rank correlation coefficient (*ρ*) between TIL_max_ and TIL_median_ was 0.80 (95% CI: 0.77–0.82), and the median TIL_median_:TIL_max_ ratio was 0.65 (IQR: 0.45–0.80).

**Fig. 2.**
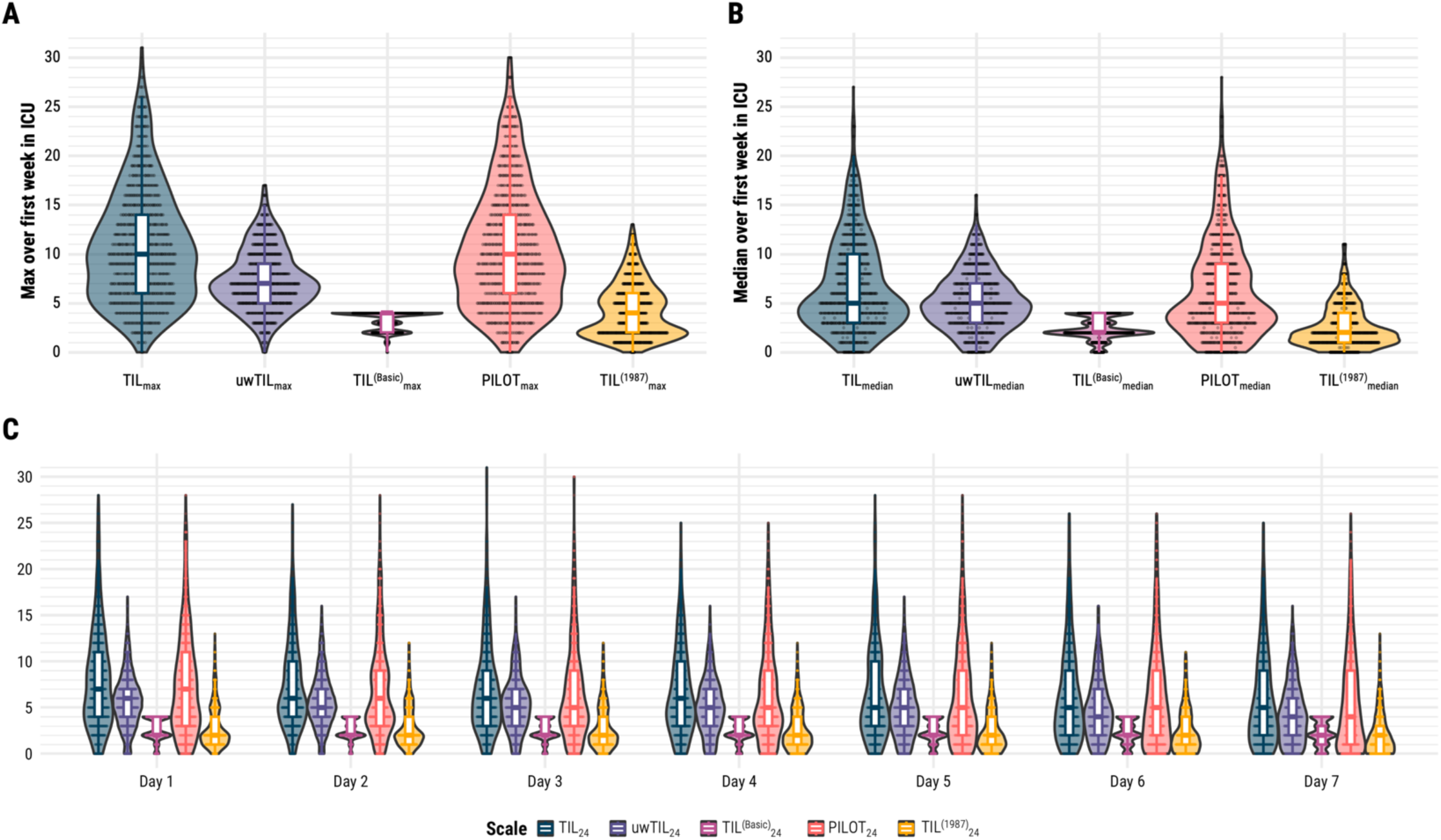
Distributions of TIL and alternative scales. Abbreviations: ICU=intensive care unit, PILOT=Paediatric Intensity Level of Therapy scale,^7^ TIL=Therapy Intensity Level scale,^8,9^ TIL^(1987)^=original Therapy Intensity Level scale published in 1987,^6^ TIL^(Basic)^=condensed TIL scale,^8^ uwTIL=unweighted TIL scale in which sub-item scores are replaced by the ascending rank index within the item. The numeric definition of each scale is listed in Table 1. (**A**) Distributions of maximum scores of TIL (i.e., TILmax) and alternative scales (i.e., uwTILmax, TIL^(Basic)^max, PILOTmax, and TIL^(1987)^max) over the first week of ICU stay. (**B**) Distribution of median scores of TIL (i.e., TILmedian) and alternative scales (i.e., uwTILmedian, TIL^(Basic)^median, PILOTmedian, and TIL^(1987)^median) over the first week of ICU stay. (**C**) Distributions of daily scores of TIL (i.e., TIL24) and alternative scales (i.e., uwTIL24, TIL^(Basic)^24, PILOT24, and TIL^(1987)^24) over the first week of ICU stay.

### Validation of TIL

The 95% CIs of *ρ* values, repeated measures correlation coefficients (*r_rm_*), and linear mixed effect regression coefficients (*β_LMER_*) of TIL with other study measures are visualised in Fig. 3. Both TIL_max_ and TIL_median_ had mildly negative correlations (- 0.26<*ρ*_mean_<-0.19) with baseline GCS, six-month GOSE, and functional outcome prognoses (Fig. 3A–B). The within-individual association of TIL_24_ with physician concerns of ICP was moderately positive (*r_rm_=*0.35 [95% CI: 0.31–0.38]) and significantly higher than that of TIL^(Basic)^_24_ (Fig. 3C). The association between ICP_median_ and TIL_median_ was moderately positive (0.35<*ρ*_mean_<0.45) with both ICP_EH_ and ICP_HR_ values, and the association between ICP_max_ and TIL_max_ was moderately positive (*ρ=*0.41 [95% CI: 0.33– 0.46]) with ICP_EH_ values. The ICP_max_ vs. TIL_max_ correlation was not significant (*ρ=*0.01 [95% CI: -0.16–0.17]) with ICP_HR_ values; however, without imputing missing ICP_HR_ values, the *ρ* was 0.43 (95% CI: 0.35–0.50). This suggests that the longitudinal missingness of ICP_HR_ (Supplementary Fig. S2) for our sample size made the ICP_max_ estimation significantly imprecise. Additionally, the within-individual association with ICP_24_ was either weak or not significant for any daily scale score according to *r_rm_* (Fig. 3C) and *β_LMER_* (Fig. 3D) values. On average, a single point increase in TIL_24_ was associated with a 0.22 (95% CI: 0.15–0.30) mmHg increase in daily mean ICP_EH_ and a 0.19 (95% CI: - 0.06–0.43) mmHg increase in daily mean ICP_HR_. These results mostly affirm the convergent validity of TIL but highlight the broad intra-patient variability between ICP and therapeutic intensity. From the distribution of ICP_24_ values at each TIL_24_ score (Fig. 4A), we observed both considerable ICP_24_ overlap across each TIL_24_ score and an overall positive relationship between TIL_24_ and ICP_24_, particularly for TIL_24_≥8.

**Fig. 3.**
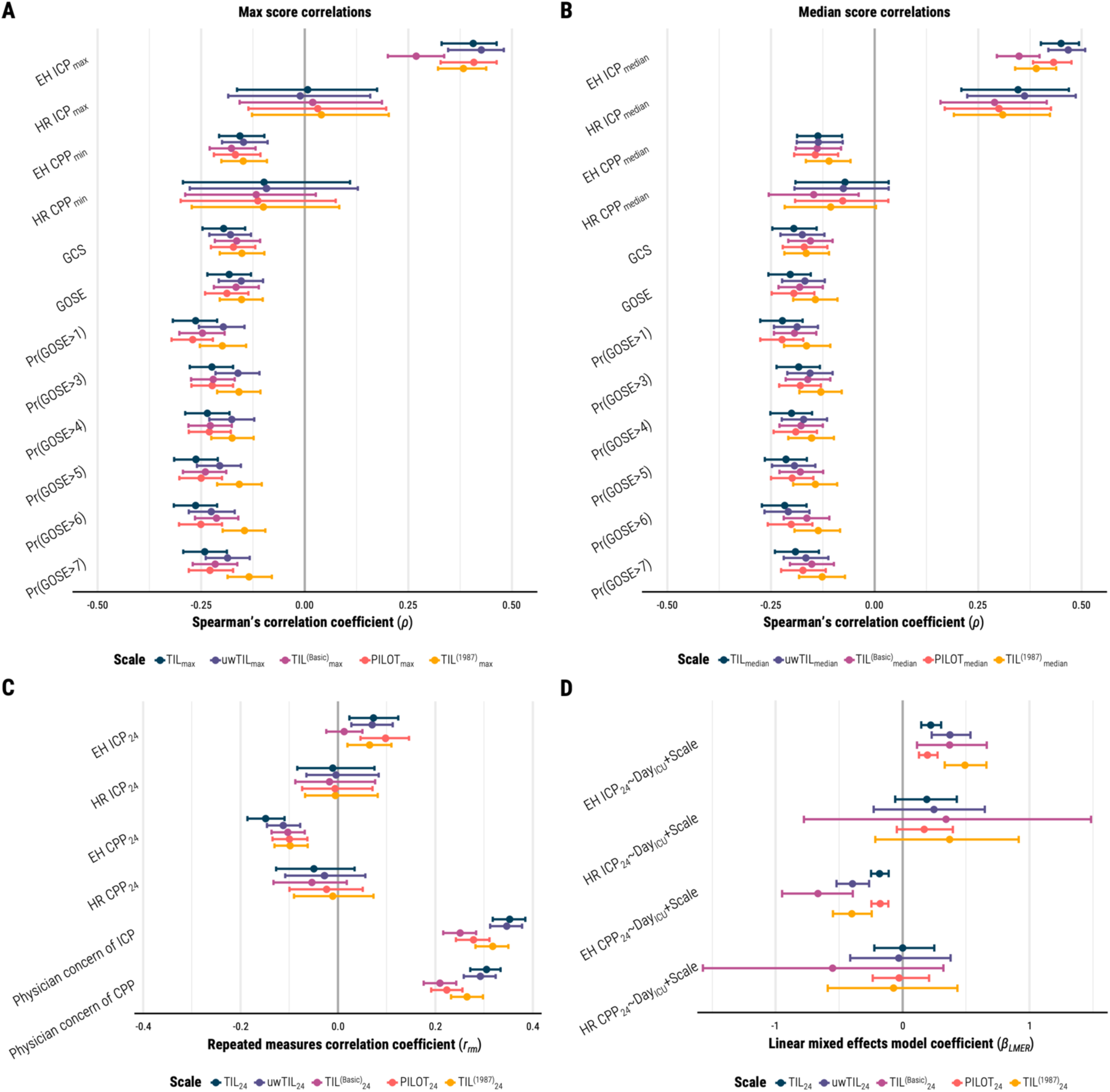
Associations of TIL and alternative scales with other clinical measures. Abbreviations: DayICU=variable representing day (from 1 to 7) of ICU stay, EH=end-hour, CPP=cerebral perfusion pressure, GCS=Glasgow Coma Scale at ICU admission, GOSE=Glasgow Outcome Scale–Extended at six months post-injury, HR=high-resolution, ICP=intracranial pressure, ICU=intensive care unit, PILOT=Paediatric Intensity Level of Therapy scale,^7^ Pr(GOSE>•)=“probability of GOSE greater than • at six months post-injury” as previously calculated from the first 24 hours of admission,^27^ TIL=Therapy Intensity Level scale,^8,9^ TIL^(1987)^=original Therapy Intensity Level scale published in 1987,^6^ TIL^(Basic)^=condensed TIL scale,^8^ uwTIL=unweighted TIL scale in which sub-item scores are replaced by the ascending rank index within the item. The numeric definition of each scale is listed in Table 1, and the calculation of daily (e.g., TIL24), maximum (e.g., TILmax), and median (e.g., TILmedian) scores are described in the Methods. The bars represent 95% confidence intervals derived from bootstrapping with 1,000 resamples of unique patients over 100 missing value imputations. (**A**) Spearman’s correlation coefficients (*ρ*) between maximum scale scores over first week of ICU stay (i.e., TILmax, uwTILmax, TIL^(Basic)^max, PILOTmax, and TIL^(1987)^max) and other clinical measures. (**B**) Spearman’s correlation coefficients (*ρ*) between median scale scores over first week of ICU stay (i.e., TILmedian, uwTILmedian, TIL^(Basic)^median, PILOTmedian, and TIL^(1987)^median) and other clinical measures. (**C**) Repeated measures correlation coefficients (*rrm*, from -1 to 1) are interpreted as the strength and direction of association between two variables after accounting for inter-patient variation. (**D**) Linear mixed effects model coefficients (*βLMER*) are interpreted as the expected difference in dependent variable (e.g., EH ICP24) per unit increase of daily scale score (e.g., TIL24) after accounting for time since ICU admission (i.e., DayICU) and inter-patient variation.

**Fig 4.**
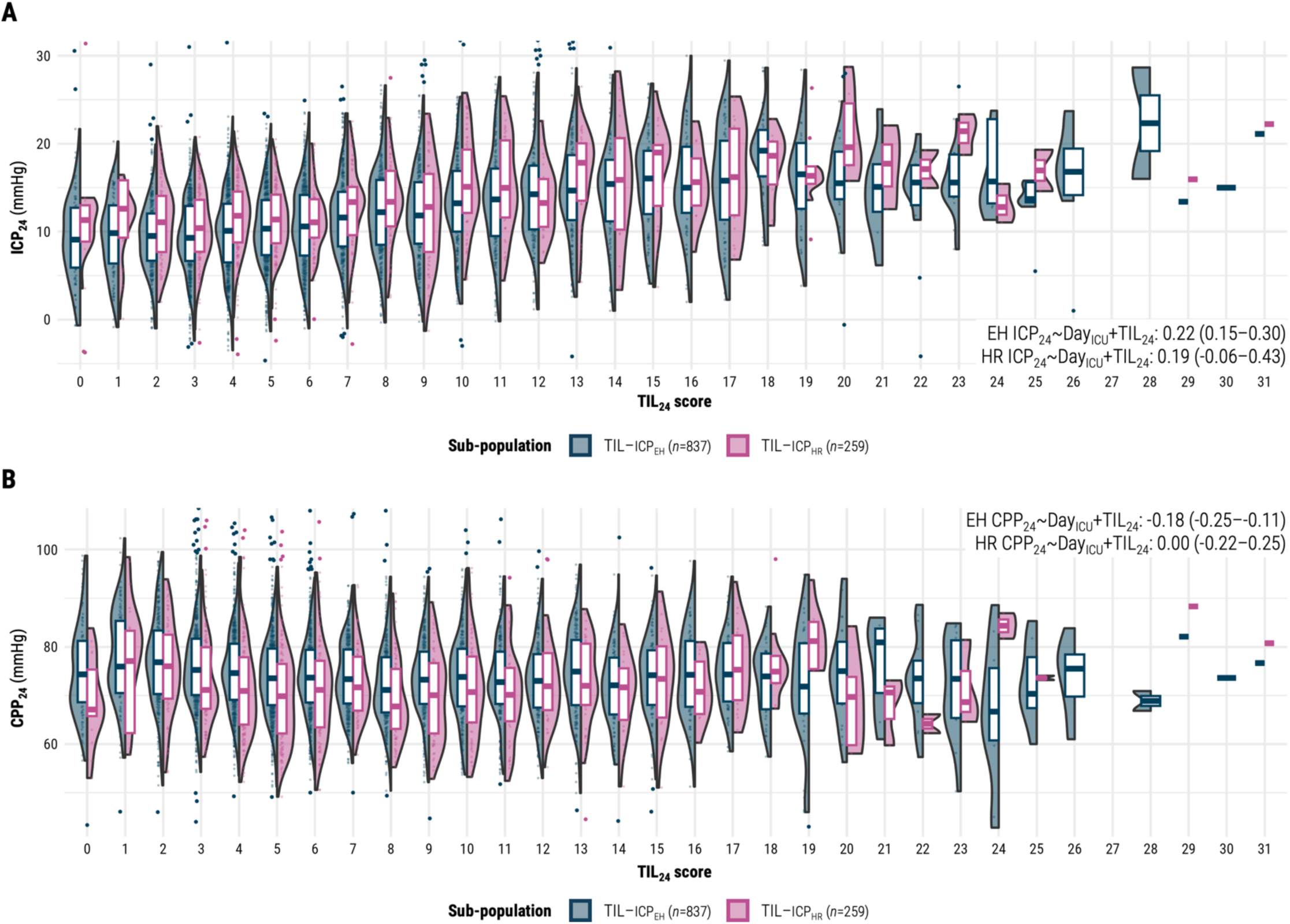
Distributions of daily intracranial pressure and cerebral perfusion pressure means per daily TIL score. Abbreviations: CPP=cerebral perfusion pressure, CPP24=mean CPP over calendar day, DayICU=variable representing day (from 1 to 7) of ICU stay, EH=end-hour, HR=high- resolution, ICP=intracranial pressure, ICP24=mean ICP over calendar day, TIL=Therapy Intensity Level scale,^8,9^ TIL24=TIL score of calendar day, TIL-ICPEH=end-hour ICP sub-population, TIL- ICPHR=high-resolution ICP sub-population. The values in each panel are the linear mixed effects model coefficients (*βLMER*) of TIL24 with 95% confidence intervals derived from bootstrapping with 1,000 resamples of unique patients over 100 missing value imputations. The width of violin plots is scaled for each population, but the width of the points inside them demonstrates relative frequency across the populations. The violin plots do not encompass outliers based on 1.5 times the interquartile range. (**A**) Distributions of ICP24 vs. TIL24 for both sub-populations. (**B**) Distributions of CPP24 vs. TIL24 for both sub-populations.

The correlation between TIL and both prior scales (i.e., PILOT and TIL^(1987)^) was positively strong for maximum, median, and daily scores (Supplementary Fig. S3), establishing the criterion validity of TIL. According to 95% CIs, the association of TIL with prior scales was stronger than that of uwTIL or TIL^(Basic)^ (Supplementary Fig. S3).

According to *ρ*, *r_rm_*, and *β_LMER_* values (Fig. 3), the associations of TIL with CPP and of TIL with physician concerns of CPP were weaker than or not significantly different from the corresponding associations with ICP. Moreover, the trend of CPP_24_ distributions over different TIL_24_ scores is not as visually apparent as that of ICP_24_ distributions over different TIL_24_ scores (Fig. 4B). These results support the discriminant validity of TIL.

In our population, 157 patients (18% of 864 assessed) were reported to experience refractory intracranial hypertension during ICU stay. TIL_max_ correctly discriminated these patients from the others 81% (95% CI: 78–84%) of the time (Fig. 5A), and TIL_median_ did so 83% (95% CI: 80–86%) of the time (Fig. 5B). This performance of TIL was significantly greater than or similar to that of all alternative scales (Fig. 5A–B). Furthermore, TIL_median_ had significantly greater discrimination performance than ICP_max_ (Fig. 5C) and ICP_median_ (Fig. 5D), respectively. The sensitivity and specificity of refractory intracranial hypertension detection at each threshold of TIL_max_, TIL_median_, TIL^(Basic)^_max_, and TIL^(Basic)^_median_ are listed in Supplementary Table S3 and visualised in Fig. 5C–D. The thresholds which maximised the sum of sensitivity and specificity were TIL_max_≥14 (sensitivity: 68% [95% CI: 62–74%], specificity: 79% [95% CI: 77–81%]) and TIL_median_≥7.5 (sensitivity: 81% [95% CI: 77–87%], specificity: 72% [95% CI: 70–75%]) (Table 3).

**Fig 5.**
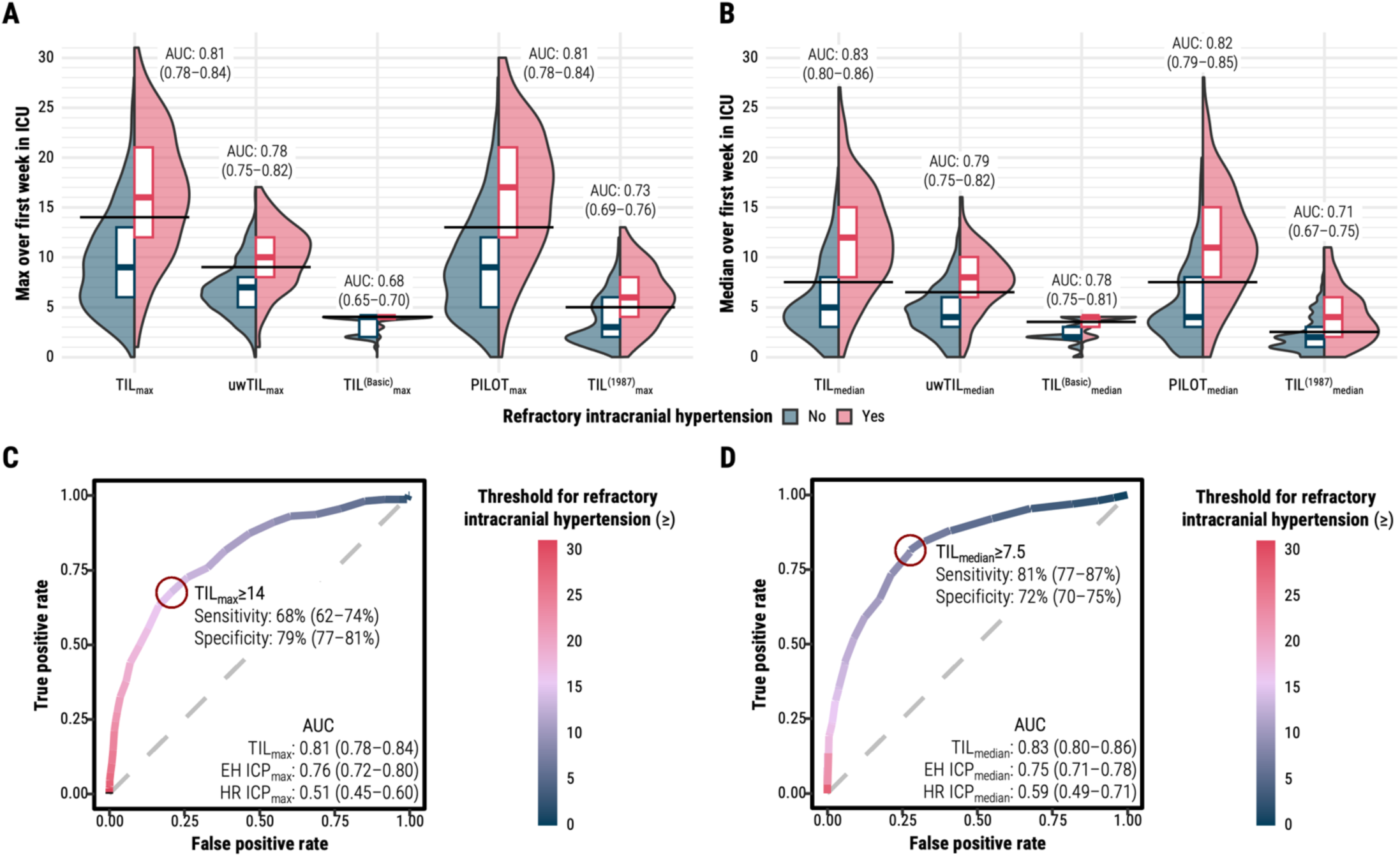
Discrimination of refractory intracranial hypertension status by TIL and alternative scale summary scores. Abbreviations: AUC=area under the receiver operating characteristic curve, EH=end-hour, HR=high-resolution, ICP=intracranial pressure, ICPmax=maximum calendar day mean of ICP over first week of ICU stay, ICPmedian=median calendar day mean of ICP over first week of ICU stay, ICU=intensive care unit, PILOT=Paediatric Intensity Level of Therapy scale,^7^ TIL=Therapy Intensity Level scale,^8,9^ TIL^(1987)^=original Therapy Intensity Level scale published in 1987,^6^ TIL^(Basic)^=condensed TIL scale,^8^ uwTIL=unweighted TIL scale in which sub-item scores are replaced by the ascending rank index within the item. The 95% confidence intervals of AUC were derived from bootstrapping with 1,000 resamples of unique patients over 100 missing value imputations. (**A**) Distributions of maximum scores of TIL (i.e., TILmax) and alternative scales (i.e., uwTILmax, TIL^(Basic)^max, PILOTmax, and TIL^(1987)^max) stratified by refractory intracranial hypertension status. The horizontal black line segments represent the thresholds which maximised the sum of sensitivity and specificity for each scale. (**B**) Distributions of median scores of TIL (i.e., TILmedian) and alternative scales (i.e., uwTILmedian, TIL^(Basic)^median, PILOTmedian, and TIL^(1987)^median) stratified by refractory intracranial hypertension status. The horizontal black line segments represent the thresholds which maximised the sum of sensitivity and specificity for each scale. (**C**) Receiver operating characteristic curve of refractory intracranial hypertension detection with TILmax. The threshold which maximised the sum of sensitivity and specificity is highlighted with the dark red circle. (**D**) Receiver operating characteristic curve of refractory intracranial hypertension detection with TILmedian. The threshold which maximised the sum of sensitivity and specificity is highlighted with the dark red circle.

**Table 3.** Optimised ranges for TIL categorisation.

| Category | Derived ranges | Performance (95% confidence intervals) |  |  | Case counts<sup>c</sup> |  | Previously proposed ranges<sup>d</sup> |
| --- | --- | --- | --- | --- | --- | --- | --- |
|  |  | Sensitivity | Specificity | Accuracy | No | Yes |  |
| Refractory intracranial hypertension<sup>a</sup> | TIL<sub>max</sub>≥14 | 68% (62–74%) | 79% (77–81%) | 77% (75–79%) | 707 | 157 | TIL<sub>max</sub>≥11 |
|  | TIL<sub>median</sub>≥7.5 | 81% (77–87%) | 72% (70–75%) | 74% (72–76%) |  |  | – |
| Day of surgical ICP control<sup>b</sup> | TIL<sub>24</sub>≥9 | 87% (83–91%) | 74% (72–76%) | 76% (74–77%) | 4916 | 585 | – |
| TIL<sup>(Basic)</sup><sub>24</sub> |  |  |  | 72% (70–73%) |  |  |  |
| (1) Basic ICU care | 1≤TIL<sub>24</sub>≤2 |  |  |  | 4932 | 568 | 1≤TIL<sub>24</sub>≤3 |
| (2) Mild | 3≤TIL<sub>24</sub>≤6 |  |  |  | 3294 | 2206 | 4≤TIL<sub>24</sub>≤7 |
| (3) Moderate | 7≤TIL<sub>24</sub>≤8 |  |  |  | 4709 | 791 | 8≤TIL<sub>24</sub>≤10 |
| (4) Extreme | TIL<sub>24</sub>≥9 |  |  |  | 3919 | 1581 | TIL<sub>24</sub>≥11 |

Abbreviations: ICP=intracranial pressure, ICU=intensive care unit, TIL=Therapy Intensity Level scale,<sup>8,9</sup> TIL<sup>(Basic)</sup>=condensed TIL scale.<sup>8</sup> The numeric definition of each scale is listed in Table 1, and the calculation of daily (e.g., TIL<sub>24</sub>), maximum (e.g., TIL<sub>max</sub>), and median (e.g., TIL<sub>median</sub>) scores is described in the Methods. The 95% confidence intervals of performance metrics were derived from bootstrapping with 1,000 resamples of unique patients over 100 missing value imputations.
<sup>a</sup>Refractory intracranial hypertension was defined as recurrent, sustained (i.e., of at least ten minutes) increases of ICP above 20 mmHg despite medical ICP management during ICU stay. This information was recorded by attending physicians in patient discharge summaries.
<sup>b</sup>If a decompressive craniectomy was performed as a last resort for refractory intracranial hypertension, each of the days following the operation were also considered days of surgical ICP control.
<sup>c</sup>For refractory intracranial hypertension, case counts represent the number of patients (with non-missing values) without (i.e., No) and with (i.e., Yes) refractory intracranial hypertension. For day of surgical ICP control and TIL<sup>(Basic)</sup><sub>24</sub>, case counts represent the number of non-missing TIL assessments not in (i.e., No) and in (i.e., Yes) the given category.
<sup>d</sup>Thresholds were previously proposed by the interagency panel which developed TIL based on expert opinion.<sup>8</sup>

### TIL component items

Whilst there was wide variation in item combinations per TIL_24_ score (i.e., sum of median scores was often under diagonal line in Fig. 6A), the average order of therapeutic escalation was fairly consistent: position, sedation, CPP management, ventilatory management, neuromuscular blockade, hyperosmolar therapy, temperature control, and then surgery for refractory ICP. Surgical control of ICP occurred in over 50% of reported cases at each TIL_24_ above 18 (Fig. 6A), but the threshold which maximised the sum of sensitivity and specificity in detecting surgical ICP control was TIL_24_≥9 (Table 3, performance at each threshold is listed in Supplementary Table S4). The inter-item *r_rm_* values of TIL_24_ (Supplementary Fig. S4) were mostly positive except for cerebrospinal fluid (CSF) drainage, which did not correlate significantly with most other items, and decompressive craniectomy, which did not correlate significantly with CSF, ventilatory, or temperature control. Consistent with Fig. 6A, this result suggested that CSF drainage and decompressive craniectomy were the most variably applied therapies across study ICUs. The Cronbach’s alpha (*α*) value of TIL_24_ was, at best, 0.65 (95% CI: 0.62–0.68) and lower (albeit, not significantly) than that of uwTIL_24_ at each day of ICU stay (Supplementary Fig. S5). However, since TIL is a formative scale (i.e., the construct is multidimensional and defined by the items), high inter-item correlation and *α* values are not necessary for item validation.^17^ Amongst all TIL_24_ items, sedation was most strongly correlated with adjusted TIL_24_ scores and physician concerns of ICP (Fig. 6B). From 10≤TIL_24_≤20, a plateau effect of high-dose sedation combined with neuromuscular blockade was observed in most cases (Fig. 6A). When accounting for all other TIL_24_ sub-items, time since ICU admission, as well as inter-patient variability, ventilation, mannitol administration, and hypertonic saline administration were most strongly associated with ICP_24_ and vasopressors were most strongly associated with CPP_24_ (Fig. 6C).

**Fig 6.**
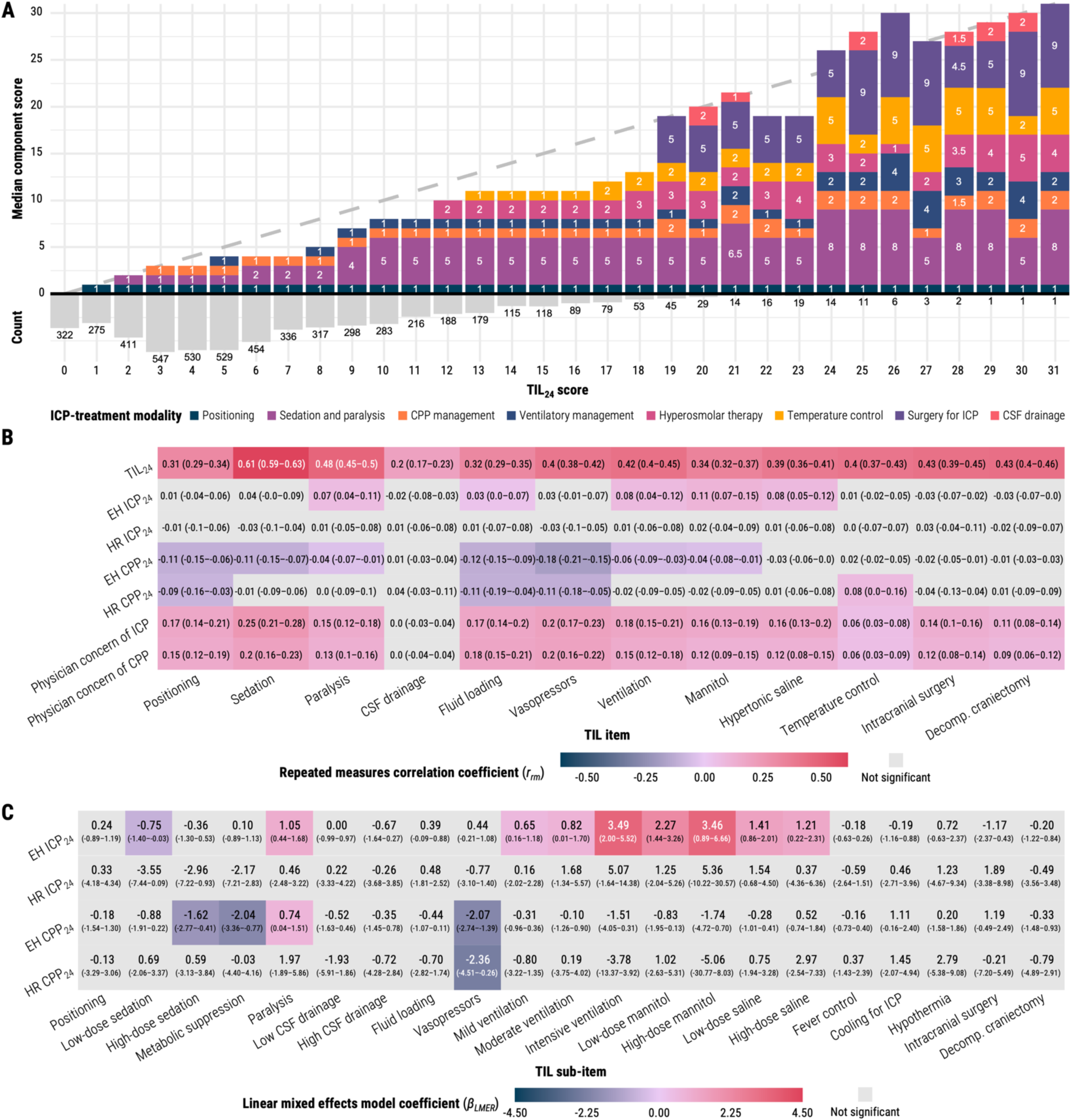
Association of TIL component items with TIL24 and other study measures. Abbreviations: CPP=cerebral perfusion pressure, CPP24=mean CPP over calendar day, CSF=cerebrospinal fluid, EH=end-hour, HR=high-resolution, ICP=intracranial pressure, ICP24=mean ICP over calendar day, ICU=intensive care unit, TIL=Therapy Intensity Level scale,^8,9^ TIL24=TIL score of calendar day. The 95% confidence intervals of *rrm* and *βLMER* values were derived from bootstrapping with 1,000 resamples of unique patients over 100 missing value imputations. (**A**) Median component score of each ICP-treatment modality (Table 1) per each TIL24 score. The histogram under the *x*-axis represents the relative frequency and count of each TIL24 score in the population, and diagonal dashed line represents the TIL24 score on both axes. If the sum of median item scores does not equal the corresponding TIL24 score, this can be interpreted as high variability in the combination of simultaneously applied therapies at that TIL24 score. (**B**) The repeated measures correlation coefficients (*rrm*, from -1 to 1) are interpreted as the strength and direction of association between two variables after accounting for inter-patient variation. The component score of each item (Table 1, *x*-axis) was subtracted from the TIL24 score (top row on *y*-axis) before calculating their *rrm* values. (**C**) Linear mixed effects model coefficients (*βLMER*) are interpreted as the expected difference in the dependent variable (*y*-axis) associated with the given TIL24 sub-item treatment (Table 1) after accounting for all other TIL24 sub-items, time since ICU admission, and inter-patient variation.

### _TIL_(Basic)

Based on the median TIL^(Basic)^_24_ score at each TIL_24_ score (Fig. 7A), we derived the ranges for mapping TIL_24_ onto TIL^(Basic)^_24_ in Table 3. There is, however, considerable overlap of TIL_24_ scores across TIL^(Basic)^_24_ scores (Fig. 7B), particularly in the range of 6≤TIL_24_≤10. TIL^(Basic)^_24_=3 was not the most represented score at any TIL_24_ score (Fig. 7A). TIL^(Basic)^_24_ covered up to 33% (95% CI: 31–34%) of the information (i.e., entropy) in TIL_24_, and TIL^(Basic)^_median_ covered up to 28% (95% CI: 27–30%) of the information in TIL_median_ (Fig. 7C). TIL^(Basic)^_max_ only covered 17% (95% CI: 16–18%) of the information in TIL_max_ (Fig. 7C).

**Fig 7.**
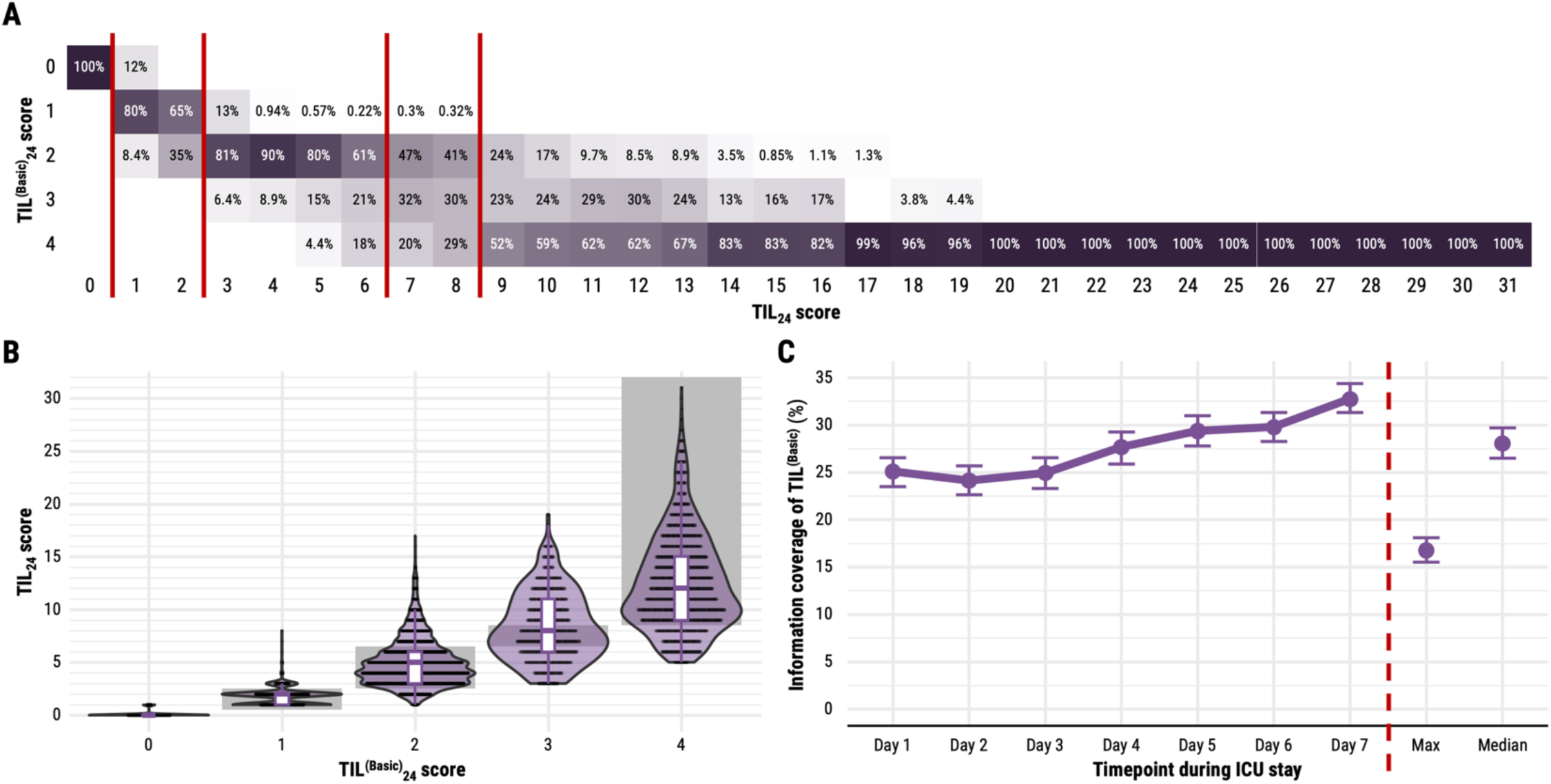
Relationship between TIL and TIL^(Basic)^. Abbreviations: AUC=area under the receiver operating characteristic curve, ICU=intensive care unit, TIL=Therapy Intensity Level scale,^8,9^ TIL^(Basic)^=condensed TIL scale.^8^ The numeric definition of each scale is listed in Table 1, and the calculation of daily (e.g., TIL24), maximum (e.g., TILmax), and median (e.g., TILmedian) scores are described in the Methods. The 95% confidence intervals of information coverage were derived from bootstrapping with 1,000 resamples of unique patients over 100 missing value imputations. (**A**) Distribution of corresponding TIL^(Basic)^24 scores per each TIL24 score. The values in each cell represent the percent of assessments at a given TIL24 score (i.e., column) corresponding to a TIL^(Basic)^24 score (i.e., row). The vertical, dark red lines represent cut-offs across which the median corresponding TIL^(Basic)^24 score per TIL24 score changes. (**B**) Distribution of corresponding TIL24 scores per each TIL^(Basic)^24 score. The width of violin plots is scaled for each TIL^(Basic)^24 score, but the width of the points inside them demonstrates relative frequency across the TIL^(Basic)^24 scores. The grey, shaded zones represent the range of TIL24 scores with corresponding median TIL^(Basic)^24 scores on the *x*-axis, as determined in panel (A). (**C**) The information of TIL24, TILmax, and TILmedian covered by TIL^(Basic)^24, TIL^(Basic)^max, and TIL^(Basic)^median, respectively. Information coverage is defined as the mutual information of TIL24 and TIL^(Basic)^24 (or TILmax and TIL^(Basic)^max or TILmedian and TIL^(Basic)^median) divided by the entropy of TIL24 (or TILmax or TILmedian).

## Discussion

In this work, we performed a large-scale (*n*=873), multicentre (52 ICUs, 19 countries), and prospective validation study of TIL and TIL^(Basic)^ against alternative scales. Our results support the validity of TIL as a metric for scoring ICP-directed therapeutic intensity. The dataset we used, as part of the CENTER-TBI study, not only reflects the modern variation in ICP-directed therapeutic intensity (Fig. 2 and Fig. 6A) but also illustrates the practical feasibility of daily TIL assessment: out of 885 eligible patients, 873 (99%) had daily TIL scores (Fig. 1) with less than 2.4% daily missingness (Supplementary Fig. S2).

We scrutinised and validated the use of TIL as a metric for scoring ICP-directed therapeutic intensity and for marking pathophysiological severity. The statistical construct and criterion validity measures of TIL were significantly greater or similar to those of alternative scales (Fig. 3 and Fig. 5), and TIL integrated the widest range of modern ICP treatments (Table 1). Summarised TIL scores outperformed summarised ICP values in detecting refractory intracranial hypertension. Our analysis yielded empirical ranges for interpreting TIL in terms of refractory intracranial hypertension, surgical intervention, and the condensed, TIL^(Basic)^ scores (Table 3). On a component level (Fig. 6A), TIL_24_ reflected a pattern of treatment intensity escalation consistent with clinical algorithms^2,3,5^ as well as a wide variation in treatment combinations, particularly in the use of CSF drainage and decompressive craniectomy. This finding is consistent with a previous CENTER-TBI study – which revealed inter-centre variation in TIL treatment selection and time to administration^12^ – and encourages an investigation of differences in TIL and long-term outcome between centres with known differences in ICP management strategies. In summary, our results support the use of TIL as an intermediate outcome for treatment effect, as done in previous studies.^33–35^

Due to a strong ceiling effect (Fig. 2A and Fig. 5A), TIL^(Basic)^ should not be used instead of TIL for rating maximum treatment intensity. TIL^(Basic)^_24_ and TIL_median_ covered up to 33% of the information in TIL_24_ (Fig. 7C), but the TIL^(Basic)^_24_ associations with physician concerns of ICP were significantly worse than those of TIL_24_ (Fig. 3C). TIL should always be preferred to TIL^(Basic)^, but we believe daily or median TIL^(Basic)^ can be a suitable alternative when daily or median TIL assessment is infeasible.

Moreover, we evaluated TIL with both end-hour (ICP_EH_) and high-resolution (ICP_HR_) ICP values. ICP_HR_, if available, should be considered the gold standard in terms of precision and granularity of the information provided, and neuromonitoring-related results from the ICP_HR_ population should generally take precedence.^14^ However, 67% of expected ICP_HR_ values were missing on day one of ICU stay (Supplementary Fig. S2), likely due to the time required to arrange high-resolution data collection. Consequently, estimates of high- resolution ICP_max_ were significantly affected by missing value imputation and became imprecise at our sample size (Fig. 3A). In these cases, results from the ICP_EH_ population served as a valuable reference on a substantially larger sample size (Table 2) since ICP_EH_ and CPP_EH_ have been shown to be fair end-hour representations of ICP_HR_ and CPP_HR_, respectively, in CENTER-TBI.^14^ The considerable overlap of ICP_24_ values across TIL_24_ scores (both at low and high levels of ICP, Fig. 4A) and the insignificant-to-weak, within-individual association between ICP_24_ and TIL_24_ (Fig. 3C–D) highlight the need to account for therapeutic intensity when interpreting ICP. Additionally, the higher median ICP_24_ values for TIL_24_≥8 (Fig. 4A) may suggest that clinicians accept a slightly higher ICP when balancing the risks of elevating therapeutic intensity against those of intracranial hypertension.

We see three main opportunities to improve TIL. First, the item scores of TIL and its predecessors (i.e., PILOT and TIL^(1987)^) were not derived empirically. Data-driven techniques, such as confirmatory factor analysis,^28^ can be used to derive scoring configurations which optimise a defined objective (e.g., maximal separation of patients). However, data-driven scores do not necessarily reflect the intended construct (i.e., treatment risk and complexity),^36^ and, in general, item scoring does not have an appreciable impact on overall scale performance.^28^ Second, the items of TIL must evolve as therapeutic approaches to ICP management evolve. TIL discriminated refractory intracranial hypertension status significantly better than TIL^(1987)^ (Fig. 5A–B) because TIL updated TIL^(1987)^ with six additional items (Table 1). We recommend updating and re-evaluating TIL each time ICP-treatment modalities or their perceived risks change. Finally, the development of TIL was largely informed by the perspective of ICU practices in high-income countries.^8^ Likewise, this assessment was performed in a cohort of patients across Europe and Israel. Especially given the disproportionately higher burden of TBI in low- and middle-income countries,^37^ it is imperative to test and, if necessary, adapt TIL to a more inclusive, global population of TBI.

By design, TIL does not encompass all facets of modern intensive care for TBI patients. Brain tissue oxygen tension (PbtO_2_),^38^ cerebral microdialysis,^39^ and brain temperature^40^ have emerged as multimodal, neuromonitoring targets which may affect ICU management in addition to ICP or CPP. Therefore, TIL should be interpreted not as general treatment intensity but rather as the intensity of ICP-directed therapy specifically. We encourage the development and validation of clinical scales assessing the intensity of TBI treatments directed at other physiological targets. Since treatments for other targets often overlap with those for ICP or CPP (e.g., vasopressors target both PbtO_2_ and CPP),^2^ we also promote a consolidation of all TBI treatments in an overall therapeutic intensity scale which considers the effect of each treatment on multiple physiological targets.

We recognise several limitations of our analysis. Whilst numerous investigators assessed TIL across the study ICUs, each TIL score was only assessed once. Therefore, we could not evaluate the interrater reliability of TIL. Similarly, data needed to calculate the full TIL score was only recorded once a day, so we could not determine if a daily assessment frequency was sufficient. Since the prior TIL validation study reported a high interrater reliability and recommended a daily assessment frequency,^9^ we assumed both to be true. The results from the Randomised Evaluation of Surgery with Craniectomy for Uncontrollable Elevation of Intracranial Pressure (RESCUEicp) trial^41^ – published amidst CENTER-TBI patient recruitment in 2016 – have likely changed the global frequency and perceived intensity of decompressive craniectomy for TBI. Therefore, we recognise the potentially confounding effect of the trial results on treatment decision making for some patients in the CENTER-TBI population and encourage a potential reappraisal of the therapeutic intensity of decompressive craniectomy through expert discussion and statistical validation. The physician impressions (i.e., physician concerns of ICP and CPP and refractory intracranial hypertension status) were subjective, and we did not have enough information to account for interrater variability. Therefore, these scores and labels should be considered unrefined. Finally, because of limited dosage data for numerical treatments (i.e., CSF drainage, ventilation, hyperosmolar therapy, and temperature control), we did not test alternative sub-item categorisations.

## Conclusion

TIL is a valid, generalisable measurement of ICP management amongst neuro-monitored TBI patients in the ICU. On all validation metrics, TIL performs at least as well as its alternatives and considers the widest range of modern treatment strategies. TIL’s component scores over increasing TIL reflect a clinically credible order of treatment escalation, from head positioning to ICP-directed surgery. TIL^(Basic)^ is not suitable for evaluating maximum treatment intensity, but daily TIL^(Basic)^ and median TIL^(Basic)^ can cover up to a third of the information in TIL. In the setting of clinical ICP management, TIL is a more sensitive marker of pathophysiological severity than ICP and can be considered an intermediate outcome after TBI.

## Data Availability

Individual participant data are available online, conditional to approved online study proposal, with no end date at https://www.center-tbi.eu/data. Signed confirmation of a data access agreement is required, and all access must comply with regulatory restrictions imposed on the original study. All analytic code used to perform the statistical analyses are publicly available online at: https://github.com/sbhattacharyay/CENTER-TBI_TIL.

https://github.com/sbhattacharyay/CENTER-TBI_TIL

## Transparency, Rigor and Reproducibility Summary

The CENTER-TBI study was pre-registered at clinicaltrials.gov (NCT02210221, https://clinicaltrials.gov/ct2/show/NCT02210221). The analysis plan was registered after beginning data collection but before data analysis at https://www.center-tbi.eu/data/approved-proposals (#491), and the lead author with primary responsibility for the analysis certifies that the analysis plan was pre-specified. A sample size of 903 patients was planned based on availability of critically ill, ICP-monitored, adult TBI patients recruited for CENTER-TBI. Actual sample size was 873, as 18 patients had a documented decision to WLST on the first day of ICU stay and 12 additional patients did not have daily TIL scores assessed. A patient inclusion diagram is provided (Fig. 1). TIL scoring and clinical data entry was performed by investigators who were aware of relevant characteristics of the participants. Participants were recruited between 19 December 2014 and 17 December 2017, and data (including follow-up results) were collected until 31 March 2021. High-resolution waveforms were stored directly from bedside monitoring software, as described in the Methods and Materials. Variability amongst different TIL assessors is not expected to be significant based on the established high interrater reliability of TIL.^9^ All equipment and software used to perform imaging and preprocessing are widely available from commercial sources or open source repositories. The clinimetric validation procedure and the primary clinical metric (TIL) are established standards in the field, based on previously published results^9,28^ and this study. The assumption of bootstrapping-derived confidence intervals is that the sample is representative of the population. This study is, itself, an external validation, and internal replication by the study group was performed. Individual participant data are available online, conditional to approved online study proposal, with no end date at https://www.center-tbi.eu/data. Signed confirmation of a data access agreement is required, and all access must comply with regulatory restrictions imposed on the original study. All analytic code used to perform the statistical analyses are publicly available online at: https://github.com/sbhattacharyay/CENTER-TBI_TIL. This paper will be published under a Creative Commons Open Access license, and upon publication, will be freely available at https://www.liebertpub.com/loi/neu.

## Acknowledgments

We are grateful to the patients and families of our study for making our efforts to improve TBI care possible. S.B. would like to thank Kathleen Mitchell-Fox (Princeton University) for offering comments on the manuscript.

## Author contributions

S.B. co-conceptualised the aims, developed the methodology and design, curated, analysed, and visualised the data, acquired funding, and wrote the manuscript. E.B. curated and analysed data, acquired funding, and reviewed the manuscript. P.Z. and L.W. curated data, aided in the development of methodology, and reviewed the manuscript. EWS and DWN curated data, acquired funding, advised statistical analysis, and reviewed the manuscript. A.I.R.M. and D.K.M. curated data, acquired funding, co-conceptualised the aims, co-developed the methodology, and reviewed the manuscript. A.E. served as principal investigator, curated data, conceptualised the aims, co-developed the methodology, and reviewed the manuscript. All authors read and approved the final manuscript.

## The CENTER-TBI investigators and participants

The co-lead investigators of CENTER-TBI are designated with an asterisk (*), and their contact email addresses are listed below.

Cecilia Åkerlund^1^, Krisztina Amrein^2^, Nada Andelic^3^, Lasse Andreassen^4^, Audny Anke^5^, Anna Antoni^6^, Gérard Audibert^7^, Philippe Azouvi^8^, Maria Luisa Azzolini^9^, Ronald Bartels^10^, Pál Barzó^11^, Romuald Beauvais^12^, Ronny Beer^13^, Bo-Michael Bellander^14^, Antonio Belli^15^, Habib Benali^16^, Maurizio Berardino^17^, Luigi Beretta^9^, Morten Blaabjerg^18^, Peter Bragge^19^, Alexandra Brazinova^20^, Vibeke Brinck^21^, Joanne Brooker^22^, Camilla Brorsson^23^, Andras Buki^24^, Monika Bullinger^25^, Manuel Cabeleira^26^, Alessio Caccioppola^27^, Emiliana Calappi^27^, Maria Rosa Calvi^9^, Peter Cameron^28^, Guillermo Carbayo Lozano^29^, Marco Carbonara^27^, Simona Cavallo^17^, Giorgio Chevallard^30^, Arturo Chieregato^30^, Giuseppe Citerio^31,32^, Hans Clusmann^33^, Mark Coburn^34^, Jonathan Coles^35^, Jamie D. Cooper^36^, Marta Correia^37^, Amra Čović^38^, Nicola Curry^39^, Endre Czeiter^24^, Marek Czosnyka^26^, Claire Dahyot-Fizelier^40^, Paul Dark^41^, Helen Dawes^42^, Véronique De Keyser^43^, Vincent Degos^16^, Francesco Della Corte^44^, Hugo den Boogert^10^, Bart Depreitere^45^, Đula Đilvesi^46^, Abhishek Dixit^47^, Emma Donoghue^22^, Jens Dreier^48^, Guy-Loup Dulière^49^, Ari Ercole^47^, Patrick Esser^42^, Erzsébet Ezer^50^, Martin Fabricius^51^, Valery L. Feigin^52^, Kelly Foks^53^, Shirin Frisvold^54^, Alex Furmanov^55^, Pablo Gagliardo^56^, Damien Galanaud^16^, Dashiell Gantner^28^, Guoyi Gao^57^, Pradeep George^58^, Alexandre Ghuysen^59^, Lelde Giga^60^, Ben Glocker^61^, Jagoš Golubovic^46^, Pedro A. Gomez^62^, Johannes Gratz^63^, Benjamin Gravesteijn^64^, Francesca Grossi^44^, Russell L. Gruen^65^, Deepak Gupta^66^, Juanita A. Haagsma^64^, Iain Haitsma^67^, Raimund Helbok^13^, Eirik Helseth^68^, Lindsay Horton^69^, Jilske Huijben^64^, Peter J. Hutchinson^70^, Bram Jacobs^71^, Stefan Jankowski^72^, Mike Jarrett^21^, Ji-yao Jiang^58^, Faye Johnson^73^, Kelly Jones^52^, Mladen Karan^46^, Angelos G. Kolias^70^, Erwin Kompanje^74^, Daniel Kondziella^51^, Evgenios Kornaropoulos^47^, Lars-Owe Koskinen^75^, Noémi Kovács^76^, Ana Kowark^77^, Alfonso Lagares^62^, Linda Lanyon^58^, Steven Laureys^78^, Fiona Lecky^79,80^, Didier Ledoux^78^, Rolf Lefering^81^, Valerie Legrand^82^, Aurelie Lejeune^83^, Leon Levi^84^, Roger Lightfoot^85^, Hester Lingsma^64^, Andrew I.R. Maas^43,86,^*, Ana M. Castaño-León^62^, Marc Maegele^87^, Marek Majdan^20^, Alex Manara^88^, Geoffrey Manley^89^, Costanza Martino^90^, Hugues Maréchal^49^, Julia Mattern^91^, Catherine McMahon^92^, Béla Melegh^93^, David Menon^47,^*, Tomas Menovsky^43,86^, Ana Mikolic^64^, Benoit Misset^78^, Visakh Muraleedharan^58^, Lynnette Murray^28^, Ancuta Negru^94^, David Nelson^1^, Virginia Newcombe^47^, Daan Nieboer^64^, József Nyirádi^2^, Otesile Olubukola^79^, Matej Oresic^95^, Fabrizio Ortolano^27^, Aarno Palotie^96,97,98^, Paul M. Parizel^99^, Jean-François Payen^100^, Natascha Perera^12^, Vincent Perlbarg^16^, Paolo Persona^101^, Wilco Peul^102^, Anna Piippo- Karjalainen^103^, Matti Pirinen^96^, Dana Pisica^64^, Horia Ples^94^, Suzanne Polinder^64^, Inigo Pomposo^29^, Jussi P. Posti^104^, Louis Puybasset^105^, Andreea Radoi^106^, Arminas Ragauskas^107^, Rahul Raj^103^, Malinka Rambadagalla^108^, Isabel Retel Helmrich^64^, Jonathan Rhodes^109^, Sylvia Richardson^110^, Sophie Richter^47^, Samuli Ripatti^96^, Saulius Rocka^107^, Cecilie Roe^111^, Olav Roise^112,113^, Jonathan Rosand^114^, Jeffrey V. Rosenfeld^115^, Christina Rosenlund^116^, Guy Rosenthal^55^, Rolf Rossaint^77^, Sandra Rossi^101^, Daniel Rueckert^61^ Martin Rusnák^117^, Juan Sahuquillo^106^, Oliver Sakowitz^91,118^, Renan Sanchez-Porras^118^, Janos Sandor^119^, Nadine Schäfer^81^, Silke Schmidt^120^, Herbert Schoechl^121^, Guus Schoonman^122^, Rico Frederik Schou^123^, Elisabeth Schwendenwein^6^, Charlie Sewalt^64^, Ranjit D. Singh^102^, Toril Skandsen^124,125^, Peter Smielewski^26^, Abayomi Sorinola^126^, Emmanuel Stamatakis^47^, Simon Stanworth^39^, Robert Stevens^127^, William Stewart^128^, Ewout W. Steyerberg^64,129^, Nino Stocchetti^130^, Nina Sundström^131^, Riikka Takala^132^, Viktória Tamás^126^, Tomas Tamosuitis^133^, Mark Steven Taylor^20^, Aurore Thibaut^78^, Braden Te Ao^52^, Olli Tenovuo^104^, Alice Theadom^52^, Matt Thomas^88^, Dick Tibboel^134^, Marjolein Timmers^74^, Christos Tolias^135^, Tony Trapani^28^, Cristina Maria Tudora^94^, Andreas Unterberg^91^, Peter Vajkoczy^136^, Shirley Vallance^28^, Egils Valeinis^60^, Zoltán Vámos^50^, Mathieu van der Jagt^137^, Gregory Van der Steen^43^, Joukje van der Naalt^71^, Jeroen T.J.M. van Dijck^102^, Inge A. M. van Erp^102^, Thomas A. van Essen^102^, Wim Van Hecke^138^, Caroline van Heugten^139^, Ernest van Veen^64^, Thijs Vande Vyvere^140^, Roel P. J. van Wijk^102^, Alessia Vargiolu^32^, Emmanuel Vega^83^, Kimberley Velt^64^, Jan Verheyden^138^, Paul M. Vespa^141^, Anne Vik^124,142^, Rimantas Vilcinis^133^, Victor Volovici^67^, Nicole von Steinbüchel^38^, Daphne Voormolen^64^, Petar Vulekovic^46^, Kevin K.W. Wang^143^, Daniel Whitehouse^47^, Eveline Wiegers^64^, Guy Williams^47^, Lindsay Wilson^69^, Stefan Winzeck^47^, Stefan Wolf^144^, Zhihui Yang^114^, Peter Ylén^145^, Alexander Younsi^91^, Frederick A. Zeiler^47,146^, Veronika Zelinkova^20^, Agate Ziverte^60^, Tommaso Zoerle^27^

^1^Department of Physiology and Pharmacology, Section of Perioperative Medicine and Intensive Care, Karolinska Institutet, Stockholm, Sweden

^2^János Szentágothai Research Centre, University of Pécs, Pécs, Hungary

^3^Division of Clinical Neuroscience, Department of Physical Medicine and Rehabilitation, Oslo University Hospital and University of Oslo, Oslo, Norway

^4^Department of Neurosurgery, University Hospital Northern Norway, Tromso, Norway

^5^Department of Physical Medicine and Rehabilitation, University Hospital Northern Norway, Tromso, Norway

^6^Trauma Surgery, Medical University Vienna, Vienna, Austria

^7^Department of Anesthesiology & Intensive Care, University Hospital Nancy, Nancy, France

^8^Raymond Poincare hospital, Assistance Publique – Hopitaux de Paris, Paris, France

^9^Department of Anesthesiology & Intensive Care, S Raffaele University Hospital, Milan, Italy

^10^Department of Neurosurgery, Radboud University Medical Center, Nijmegen, The Netherlands

^11^Department of Neurosurgery, University of Szeged, Szeged, Hungary

^12^International Projects Management, ARTTIC, Munchen, Germany

^13^Department of Neurology, Neurological Intensive Care Unit, Medical University of Innsbruck, Innsbruck, Austria

^14^Department of Neurosurgery & Anesthesia & intensive care medicine, Karolinska University Hospital, Stockholm, Sweden

^15^NIHR Surgical Reconstruction and Microbiology Research Centre, Birmingham, UK

^16^Anesthesie-Réanimation, Assistance Publique – Hopitaux de Paris, Paris, France

^17^Department of Anesthesia & ICU, AOU Città della Salute e della Scienza di Torino - Orthopedic and Trauma Center, Torino, Italy

^18^Department of Neurology, Odense University Hospital, Odense, Denmark

^19^BehaviourWorks Australia, Monash Sustainability Institute, Monash University, Victoria, Australia

^20^Department of Public Health, Faculty of Health Sciences and Social Work, Trnava University, Trnava, Slovakia

^21^Quesgen Systems Inc., Burlingame, California, USA

^22^Australian & New Zealand Intensive Care Research Centre, Department of Epidemiology and Preventive Medicine, School of Public Health and Preventive Medicine, Monash University, Melbourne, Australia

^23^Department of Surgery and Perioperative Science, Umeå University, Umeå, Sweden

^24^Department of Neurosurgery, Medical School, University of Pécs, Hungary and Neurotrauma Research Group, János Szentágothai Research Centre, University of Pécs, Hungary

^25^Department of Medical Psychology, Universitätsklinikum Hamburg-Eppendorf, Hamburg, Germany

^26^Brain Physics Lab, Division of Neurosurgery, Dept of Clinical Neurosciences, University of Cambridge, Addenbrooke’s Hospital, Cambridge, UK

^27^Neuro ICU, Fondazione IRCCS Cà Granda Ospedale Maggiore Policlinico, Milan, Italy

^28^ANZIC Research Centre, Monash University, Department of Epidemiology and Preventive Medicine, Melbourne, Victoria, Australia

^29^Department of Neurosurgery, Hospital of Cruces, Bilbao, Spain

^30^NeuroIntensive Care, Niguarda Hospital, Milan, Italy

^31^School of Medicine and Surgery, Università Milano Bicocca, Milano, Italy

^32^NeuroIntensive Care Unit, Department Neuroscience, IRCCS Fondazione San Gerardo dei Tintori, Monza, Italy

^33^Department of Neurosurgery, Medical Faculty RWTH Aachen University, Aachen, Germany

^34^Department of Anesthesiology and Intensive Care Medicine, University Hospital Bonn, Bonn, Germany

^35^Department of Anesthesia & Neurointensive Care, Cambridge University Hospital NHS Foundation Trust, Cambridge, UK

^36^School of Public Health & PM, Monash University and The Alfred Hospital, Melbourne, Victoria, Australia

^37^Radiology/MRI department, MRC Cognition and Brain Sciences Unit, Cambridge, UK

^38^Institute of Medical Psychology and Medical Sociology, Universitätsmedizin Göttingen, Göttingen, Germany

^39^Oxford University Hospitals NHS Trust, Oxford, UK

^40^Intensive Care Unit, CHU Poitiers, Potiers, France

^41^University of Manchester NIHR Biomedical Research Centre, Critical Care Directorate, Salford Royal Hospital NHS Foundation Trust, Salford, UK

^42^Movement Science Group, Faculty of Health and Life Sciences, Oxford Brookes University, Oxford, UK

^43^Department of Neurosurgery, Antwerp University Hospital, Edegem, Belgium

^44^Department of Anesthesia & Intensive Care, Maggiore Della Carità Hospital, Novara, Italy

^45^Department of Neurosurgery, University Hospitals Leuven, Leuven, Belgium

^46^Department of Neurosurgery, Clinical centre of Vojvodina, Faculty of Medicine, University of Novi Sad, Novi Sad, Serbia

^47^Division of Anaesthesia, University of Cambridge, Addenbrooke’s Hospital, Cambridge, UK

^48^Center for Stroke Research Berlin, Charité – Universitätsmedizin Berlin, corporate member of Freie Universität Berlin, Humboldt-Universität zu Berlin, and Berlin Institute of Health, Berlin, Germany

^49^Intensive Care Unit, CHR Citadelle, Liège, Belgium

^50^Department of Anaesthesiology and Intensive Therapy, University of Pécs, Pécs, Hungary

^51^Departments of Neurology, Clinical Neurophysiology and Neuroanesthesiology, Region Hovedstaden Rigshospitalet, Copenhagen, Denmark

^52^National Institute for Stroke and Applied Neurosciences, Faculty of Health and Environmental Studies, Auckland University of Technology, Auckland, New Zealand

^53^Department of Neurology, Erasmus MC, Rotterdam, the Netherlands

^54^Department of Anesthesiology and Intensive care, University Hospital Northern Norway, Tromso, Norway

^55^Department of Neurosurgery, Hadassah-hebrew University Medical center, Jerusalem, Israel

^56^Fundación Instituto Valenciano de Neurorrehabilitación (FIVAN), Valencia, Spain

^57^Department of Neurosurgery, Shanghai Renji hospital, Shanghai Jiaotong University/school of medicine, Shanghai, China

^58^Karolinska Institutet, INCF International Neuroinformatics Coordinating Facility, Stockholm, Sweden

^59^Emergency Department, CHU, Liège, Belgium

^60^Neurosurgery clinic, Pauls Stradins Clinical University Hospital, Riga, Latvia

^61^Department of Computing, Imperial College London, London, UK

^62^Department of Neurosurgery, Hospital Universitario 12 de Octubre, Madrid, Spain

^63^Department of Anesthesia, Critical Care and Pain Medicine, Medical University of Vienna, Austria

^64^Department of Public Health, Erasmus Medical Center-University Medical Center, Rotterdam, The Netherlands

^65^College of Health and Medicine, Australian National University, Canberra, Australia

^66^Department of Neurosurgery, Neurosciences Centre & JPN Apex trauma centre, All India Institute of Medical Sciences, New Delhi-110029, India

^67^Department of Neurosurgery, Erasmus MC, Rotterdam, the Netherlands

^68^Department of Neurosurgery, Oslo University Hospital, Oslo, Norway

^69^Division of Psychology, University of Stirling, Stirling, UK

^70^Division of Neurosurgery, Department of Clinical Neurosciences, Addenbrooke’s Hospital & University of Cambridge, Cambridge, UK

^71^Department of Neurology, University of Groningen, University Medical Center Groningen, Groningen, Netherlands

^72^Neurointensive Care, Sheffield Teaching Hospitals NHS Foundation Trust, Sheffield, UK

^73^Salford Royal Hospital NHS Foundation Trust Acute Research Delivery Team, Salford, UK

^74^Department of Intensive Care and Department of Ethics and Philosophy of Medicine, Erasmus Medical Center, Rotterdam, The Netherlands

^75^Department of Clinical Neuroscience, Neurosurgery, Umeå University, Umeå, Sweden

^76^Hungarian Brain Research Program - Grant No. KTIA_13_NAP-A-II/8, University of Pécs, Pécs, Hungary

^77^Department of Anaesthesiology, University Hospital of Aachen, Aachen, Germany

^78^Cyclotron Research Center, University of Liège, Liège, Belgium

^79^Centre for Urgent and Emergency Care Research (CURE), Health Services Research Section, School of Health and Related Research (ScHARR), University of Sheffield, Sheffield, UK

^80^Emergency Department, Salford Royal Hospital, Salford UK

^81^Institute of Research in Operative Medicine (IFOM), Witten/Herdecke University, Cologne, Germany

^82^VP Global Project Management CNS, ICON, Paris, France

^83^Department of Anesthesiology-Intensive Care, Lille University Hospital, Lille, France

^84^Department of Neurosurgery, Rambam Medical Center, Haifa, Israel

^85^Department of Anesthesiology & Intensive Care, University Hospitals Southhampton NHS Trust, Southhampton, UK

^86^Department of Translational Neuroscience, Faculty of Medicine and Health Science, University of Antwerp, Antwerp, Belgium

^87^Cologne-Merheim Medical Center (CMMC), Department of Traumatology, Orthopedic Surgery and Sportmedicine, Witten/Herdecke University, Cologne, Germany

^88^Intensive Care Unit, Southmead Hospital, Bristol, Bristol, UK

^89^Department of Neurological Surgery, University of California, San Francisco, California, USA

^90^Department of Anesthesia & Intensive Care,M. Bufalini Hospital, Cesena, Italy

^91^Department of Neurosurgery, University Hospital Heidelberg, Heidelberg, Germany

^92^Department of Neurosurgery, The Walton centre NHS Foundation Trust, Liverpool, UK

^93^Department of Medical Genetics, University of Pécs, Pécs, Hungary

^94^Department of Neurosurgery, Emergency County Hospital Timisoara, Timisoara, Romania

^95^School of Medical Sciences, Örebro University, Örebro, Sweden

^96^Institute for Molecular Medicine Finland, University of Helsinki, Helsinki, Finland

^97^Analytic and Translational Genetics Unit, Department of Medicine; Psychiatric & Neurodevelopmental Genetics Unit, Department of Psychiatry; Department of Neurology, Massachusetts General Hospital, Boston, MA, USA

^98^Program in Medical and Population Genetics; The Stanley Center for Psychiatric Research, The Broad Institute of MIT and Harvard, Cambridge, MA, USA

^99^Department of Radiology, University of Antwerp, Edegem, Belgium

^100^Department of Anesthesiology & Intensive Care, University Hospital of Grenoble, Grenoble, France

^101^Department of Anesthesia & Intensive Care, Azienda Ospedaliera Università di Padova, Padova, Italy

^102^Dept. of Neurosurgery, Leiden University Medical Center, Leiden, The Netherlands and Dept. of Neurosurgery, Medical Center Haaglanden, The Hague, The Netherlands

^103^Department of Neurosurgery, Helsinki University Central Hospital

^104^Division of Clinical Neurosciences, Department of Neurosurgery and Turku Brain Injury Centre, Turku University Hospital and University of Turku, Turku, Finland

^105^Department of Anesthesiology and Critical Care, Pitié -Salpêtrière Teaching Hospital, Assistance Publique, Hôpitaux de Paris and University Pierre et Marie Curie, Paris, France

^106^Neurotraumatology and Neurosurgery Research Unit (UNINN), Vall d’Hebron Research Institute, Barcelona, Spain

^107^Department of Neurosurgery, Kaunas University of technology and Vilnius University, Vilnius, Lithuania

^108^Department of Neurosurgery, Rezekne Hospital, Latvia

^109^Department of Anaesthesia, Critical Care & Pain Medicine NHS Lothian & University of Edinburg, Edinburgh, UK

^110^Director, MRC Biostatistics Unit, Cambridge Institute of Public Health, Cambridge, UK

^111^Department of Physical Medicine and Rehabilitation, Oslo University Hospital/University of Oslo, Oslo, Norway

^112^Division of Orthopedics, Oslo University Hospital, Oslo, Norway

^113^Institue of Clinical Medicine, Faculty of Medicine, University of Oslo, Oslo, Norway

^114^Broad Institute, Cambridge MA Harvard Medical School, Boston MA, Massachusetts General Hospital, Boston MA, USA

^115^National Trauma Research Institute, The Alfred Hospital, Monash University, Melbourne, Victoria, Australia

^116^Department of Neurosurgery, Odense University Hospital, Odense, Denmark

^117^International Neurotrauma Research Organisation, Vienna, Austria

^118^Klinik für Neurochirurgie, Klinikum Ludwigsburg, Ludwigsburg, Germany

^119^Division of Biostatistics and Epidemiology, Department of Preventive Medicine, University of Debrecen, Debrecen, Hungary

^120^Department Health and Prevention, University Greifswald, Greifswald, Germany

^121^Department of Anaesthesiology and Intensive Care, AUVA Trauma Hospital, Salzburg, Austria

^122^Department of Neurology, Elisabeth-TweeSteden Ziekenhuis, Tilburg, the Netherlands

^123^Department of Neuroanesthesia and Neurointensive Care, Odense University Hospital, Odense, Denmark

^124^Department of Neuromedicine and Movement Science, Norwegian University of

Science and Technology, NTNU, Trondheim, Norway

^125^Department of Physical Medicine and Rehabilitation, St.Olavs Hospital, Trondheim University Hospital, Trondheim, Norway

^126^Department of Neurosurgery, University of Pécs, Pécs, Hungary

^127^Division of Neuroscience Critical Care, John Hopkins University School of Medicine, Baltimore, USA

^128^Department of Neuropathology, Queen Elizabeth University Hospital and University of Glasgow, Glasgow, UK

^129^Dept. of Department of Biomedical Data Sciences, Leiden University Medical Center, Leiden, The Netherlands

^130^Department of Pathophysiology and Transplantation, Milan University, and Neuroscience ICU, Fondazione IRCCS Cà Granda Ospedale Maggiore Policlinico, Milano, Italy

^131^Department of Radiation Sciences, Biomedical Engineering, Umeå University, Umeå, Sweden

^132^Perioperative Services, Intensive Care Medicine and Pain Management, Turku

University Hospital and University of Turku, Turku, Finland

^133^Department of Neurosurgery, Kaunas University of Health Sciences, Kaunas, Lithuania

^134^Intensive Care and Department of Pediatric Surgery, Erasmus Medical Center, Sophia Children’s Hospital, Rotterdam, The Netherlands

^135^Department of Neurosurgery, Kings college London, London, UK

^136^Neurologie, Neurochirurgie und Psychiatrie, Charité – Universitätsmedizin Berlin, Berlin, Germany

^137^Department of Intensive Care Adults, Erasmus MC– University Medical Center Rotterdam, Rotterdam, the Netherlands

^138^icoMetrix NV, Leuven, Belgium

^139^Movement Science Group, Faculty of Health and Life Sciences, Oxford Brookes University, Oxford, UK

^140^Radiology Department, Antwerp University Hospital, Edegem, Belgium

^141^Director of Neurocritical Care, University of California, Los Angeles, USA

^142^Department of Neurosurgery, St.Olavs Hospital, Trondheim University Hospital, Trondheim, Norway

^143^Department of Emergency Medicine, University of Florida, Gainesville, Florida, USA

^144^Department of Neurosurgery, Charité – Universitätsmedizin Berlin, corporate member of Freie Universität Berlin, Humboldt-Universität zu Berlin, and Berlin Institute of Health, Berlin, Germany

^145^VTT Technical Research Centre, Tampere, Finland

^146^Section of Neurosurgery, Department of Surgery, Rady Faculty of Health Sciences, University of Manitoba, Winnipeg, MB, Canada

*Co-lead investigators: (AIRM) and (DM)

## References

1. Stocchetti N, Maas AIR. Traumatic Intracranial Hypertension. N Engl J Med 2014;370(22):2121–2130; doi: 10.1056/NEJMra1208708.

2. Meyfroidt G, Bouzat P, Casaer MP, et al. Management of moderate to severe traumatic brain injury: an update for the intensivist. Intensive Care Med 2022;48(6):649–666; doi: 10.1007/s00134-022-06702-4.

3. Menon DK, Ercole A. Chapter 14 - Critical care management of traumatic brain injury. Wijdicks EFM, Kramer AH. eds. Handb Clin Neurol 2017;140(Journal Article):239–274; doi: 10.1016/B978-0-444-63600-3.00014-3.

4. Stocchetti N, Carbonara M, Citerio G, et al. Severe traumatic brain injury: targeted management in the intensive care unit. Lancet Neurol 2017;16(6):452–464; doi: 10.1016/S1474-4422(17)30118-7.

5. Hawryluk GWJ, Aguilera S, Buki A, et al. A management algorithm for patients with intracranial pressure monitoring: the Seattle International Severe Traumatic Brain Injury Consensus Conference (SIBICC). Intensive Care Med 2019;45(12):1783– 1794; doi: 10.1007/s00134-019-05805-9.

6. Maset AL, Marmarou A, Ward JD, et al. Pressure-volume index in head injury. J Neurosurg 1987;67(6):832–840; doi: 10.3171/jns.1987.67.6.0832.

7. Shore PM, Hand LL, Roy L, et al. Reliability and validity of the Pediatric Intensity Level of Therapy (PILOT) scale: A measure of the use of intracranial pressure– directed therapies. Crit Care Med 2006;34(7):1981; doi: 10.1097/01.CCM.0000220765.22184.ED.

8. Maas AIR, Harrison-Felix CL, Menon D, et al. Standardizing Data Collection in Traumatic Brain Injury. J Neurotrauma 2011;28(2):177–187; doi: 10.1089/neu.2010.1617.

9. Zuercher P, Groen JL, Aries MJH, et al. Reliability and Validity of the Therapy Intensity Level Scale: Analysis of Clinimetric Properties of a Novel Approach to Assess Management of Intracranial Pressure in Traumatic Brain Injury. J Neurotrauma 2016;33(19):1768–1774; doi: 10.1089/neu.2015.4266.

10. Robba C, Graziano F, Guglielmi A, et al. Treatments for intracranial hypertension in acute brain-injured patients: grading, timing, and association with outcome. Data from the SYNAPSE-ICU study. Intensive Care Med 2023;49(1):50–61; doi: 10.1007/s00134-022-06937-1.

11. Huijben JA, Wiegers EJA, Lingsma HF, et al. Changing care pathways and between-center practice variations in intensive care for traumatic brain injury across Europe: a CENTER-TBI analysis. Intensive Care Med 2020;46(5):995–1004; doi: 10.1007/s00134-020-05965-z.

12. Huijben JA, Dixit A, Stocchetti N, et al. Use and impact of high intensity treatments in patients with traumatic brain injury across Europe: a CENTER-TBI analysis. Crit Care 2021;25(1):78; doi: 10.1186/s13054-020-03370-y.

13. Bhattacharyay S, Caruso PF, Åkerlund C, et al. Mining the contribution of intensive care clinical course to outcome after traumatic brain injury. Npj Digit Med 2023;6(1):154; doi: 10.1038/s41746-023-00895-8.

14. Zoerle T, Birg T, Carbonara M, et al. Accuracy of Manual Intracranial Pressure Recording Compared to a Computerized High-Resolution System: A CENTER-TBI Analysis. Neurocrit Care 2023; doi: 10.1007/s12028-023-01697-2.

15. Maas AIR, Menon DK, Manley GT, et al. Traumatic brain injury: progress and challenges in prevention, clinical care, and research. Lancet Neurol 2022;21(11):1004–1060; doi: 10.1016/S1474-4422(22)00309-X.

16. Cnossen MC, Huijben JA, van der Jagt M, et al. Variation in monitoring and treatment policies for intracranial hypertension in traumatic brain injury: a survey in 66 neurotrauma centers participating in the CENTER-TBI study. Crit Care 2017;21(1):233; doi: 10.1186/s13054-017-1816-9.

17. Avila ML, Stinson J, Kiss A, et al. A critical review of scoring options for clinical measurement tools. BMC Res Notes 2015;8(1):612; doi: 10.1186/s13104-015-1561-6.

18. Maas AIR, Menon DK, Steyerberg EW, et al. Collaborative European NeuroTrauma Effectiveness Research in Traumatic Brain Injury (CENTER-TBI): A Prospective Longitudinal Observational Study. Neurosurgery 2015;76(1):67–80; doi: 10.1227/NEU.0000000000000575.

19. Steyerberg EW, Wiegers E, Sewalt C, et al. Case-mix, care pathways, and outcomes in patients with traumatic brain injury in CENTER-TBI: a European prospective, multicentre, longitudinal, cohort study. Lancet Neurol 2019;18(10):923– 934; doi: 10.1016/S1474-4422(19)30232-7.

20. Doiron D, Marcon Y, Fortier I, et al. Software Application Profile: Opal and Mica: open-source software solutions for epidemiological data management, harmonization and dissemination. Int J Epidemiol 2017;46(5):1372–1378; doi: 10.1093/ije/dyx180.

21. Zeiler FA, Ercole A, Cabeleira M, et al. Patient-specific ICP Epidemiologic Thresholds in Adult Traumatic Brain Injury: A CENTER-TBI Validation Study. J Neurosurg Anesthesiol 2021;33(1):28–38; doi: 10.1097/ANA.0000000000000616.

22. Marshall LF, Marshall SB, Klauber MR, et al. The diagnosis of head injury requires a classification based on computed axial tomography. J Neurotrauma 1992;9 Suppl 1:S287–292.

23. Teasdale G, Maas A, Lecky F, et al. The Glasgow Coma Scale at 40 years: standing the test of time. The LancetNeurology 2014;13(8):844–854; doi: 10.1016/S1474-4422(14)70120-6.

24. Ercole A, Dixit A, Nelson DW, et al. Imputation strategies for missing baseline neurological assessment covariates after traumatic brain injury: A CENTER-TBI study. PLoS ONE 2021;16(8):e0253425.

25. McMillan T, Wilson L, Ponsford J, et al. The Glasgow Outcome Scale - 40 years of application and refinement. Nat Rev Neurol 2016;12(8):477–485; doi: 10.1038/nrneurol.2016.89.

26. Kunzmann K, Wernisch L, Richardson S, et al. Imputation of Ordinal Outcomes: A Comparison of Approaches in Traumatic Brain Injury. J Neurotrauma 2021;38(4):455–463; doi: 10.1089/neu.2019.6858.

27. Bhattacharyay S, Milosevic I, Wilson L, et al. The leap to ordinal: Detailed functional prognosis after traumatic brain injury with a flexible modelling approach. PLoS ONE 2022;17(7):e0270973; doi: 10.1371/journal.pone.0270973.

28. Boateng GO, Neilands TB, Frongillo EA, et al. Best Practices for Developing and Validating Scales for Health, Social, and Behavioral Research: A Primer. Front Public Health 2018;6:149; doi: 10.3389/fpubh.2018.00149.

29. Bakdash JZ, Marusich LR. Repeated Measures Correlation. Front Psychol 2017;8:456; doi: 10.3389/fpsyg.2017.00456.

30. van Buuren S, Groothuis-Oudshoorn CGM. mice: Multivariate Imputation by Chained Equations in R. J Stat Softw 2011;45(3):1–67; doi: 10.18637/jss.v045.i03.

31. Gravesteijn BY, Sewalt CA, Venema E, et al. Missing Data in Prediction Research: A Five-Step Approach for Multiple Imputation, Illustrated in the CENTER- TBI Study. J Neurotrauma 2021;38(13):1842–1857; doi: 10.1089/neu.2020.7218.

32. Honaker J King, G, Blackwell,M. Amelia II: A Program for Missing Data. J Stat Softw 2011;45(7); doi: 10.18637/jss.v045.i07.

33. Beqiri E, Smielewski P, Robba C, et al. Feasibility of individualised severe traumatic brain injury management using an automated assessment of optimal cerebral perfusion pressure: the COGiTATE phase II study protocol. BMJ Open 2019;9(9):e030727; doi: 10.1136/bmjopen-2019-030727.

34. Tas J, Beqiri E, van Kaam RC, et al. Targeting Autoregulation-Guided Cerebral Perfusion Pressure after Traumatic Brain Injury (COGiTATE): A Feasibility Randomized Controlled Clinical Trial. J Neurotrauma 2021;38(20):2790–2800; doi: 10.1089/neu.2021.0197.

35. van Erp IAM, van Essen TA, Fluiter K, et al. Safety and efficacy of C1-inhibitor in traumatic brain injury (CIAO@TBI): study protocol for a randomized, placebo- controlled, multi-center trial. Trials 2021;22(1):874; doi: 10.1186/s13063-021-05833-1.

36. Schreiber JB. Issues and recommendations for exploratory factor analysis and principal component analysis. Res Soc Adm Pharm 2021;17(5):1004–1011; doi: 10.1016/j.sapharm.2020.07.027.

37. Clark D, Joannides A, Adeleye AO, et al. Casemix, management, and mortality of patients receiving emergency neurosurgery for traumatic brain injury in the Global Neurotrauma Outcomes Study: a prospective observational cohort study. Lancet Neurol 2022;21(5):438–449; doi: 10.1016/S1474-4422(22)00037-0.

38. Okonkwo DO, Shutter LA, Moore C, et al. Brain Oxygen Optimization in Severe Traumatic Brain Injury Phase-II: A Phase II Randomized Trial*. Crit Care Med 2017;45(11):1907; doi: 10.1097/CCM.0000000000002619.

39. Hutchinson PJ, Jalloh I, Helmy A, et al. Consensus statement from the 2014 International Microdialysis Forum. Intensive Care Med 2015;41(9):1517–1528; doi: 10.1007/s00134-015-3930-y.

40. Yokobori S, Yokota H. Targeted temperature management in traumatic brain injury. J Intensive Care 2016;4(1):28; doi: 10.1186/s40560-016-0137-4.

41. Hutchinson PJ, Kolias AG, Timofeev IS, et al. Trial of Decompressive Craniectomy for Traumatic Intracranial Hypertension. N Engl J Med 2016;375(12):1119–1130; doi: 10.1056/NEJMoa1605215.

