## Supplementary for "The Therapy Intensity Level scale for traumatic brain injury: clinimetric assessment on neuro-monitored patients across 52 European intensive care units"

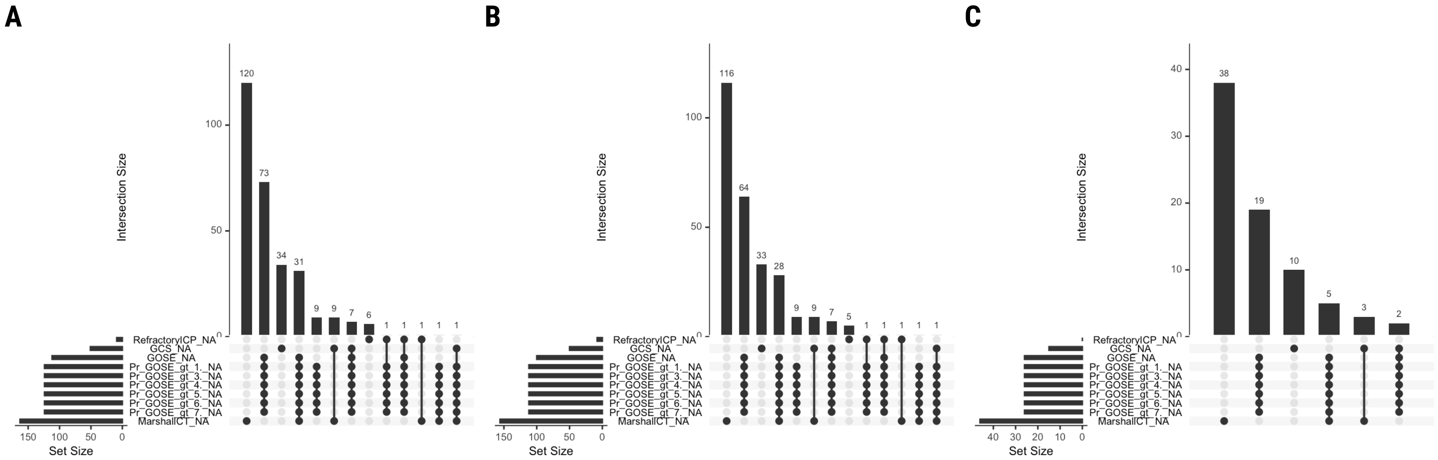


**Supplementary Figure S1. Missingness of static study measures.** Abbreviations: GCS_NA=indicator variable representing cases with missing values for Glasgow Coma Scale at ICU admission, GOSE_NA=indicator variable representing cases with missing values for Glasgow Outcome Scale–Extended at six months post-injury, ICU=intensive care unit, MarshallCT_NA=indicator variable representing cases with missing values for Marshall CT classification at ICU admission**,** Pr_GOSE_gt_**•**._NA=indicator variable representing cases with missing values for "probability of GOSE greater than • at six months post-injury" as previously calculated from the first 24 hours of admission,[27] RefractoryICP_NA=indicator variable representing cases with missing values for refractory intracranial hypertension during ICU stay. A separate missingness UpSet plot is shown for the (**A**) TIL validation population (*n*=873), (**B**) TIL-$ICP_{EH}$ sub-population (*n*=837), and (**C**) TIL-$ICP_{HR}$ sub-population (*n*=259), as defined in Figure 1.

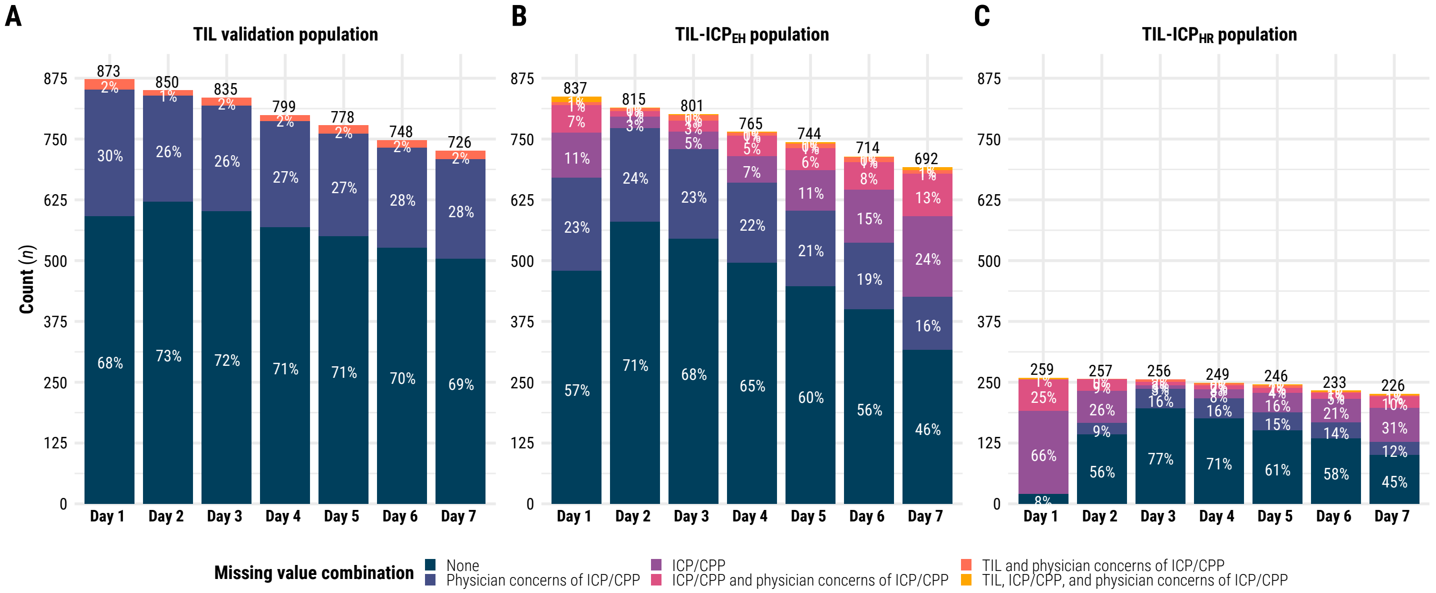


**Supplementary Figure S2. Study populations remaining in ICU over first week and missingness of longitudinal study measures.** Abbreviations: CPP=cerebral perfusion pressure, EH=end-hour, HR=high-resolution, ICP=intracranial pressure, ICU=intensive care unit, TIL=Therapy Intensity Level scale,[8,9] TIL-ICP$_{EH}$=end-hour ICP sub-population, TIL-ICP$_{HR}$=high-resolution ICP sub-population. The values above each stacked bar plot represent the number of patients remaining at each day of ICU stay (i.e., expected cases), excluding those who have already had a decision to withdraw life-sustaining therapies. The value in each component of the stacked bar plot represents the percent of expected cases per each day of ICU stay that falls into the corresponding missing value combination of that colour. A separate missingness stacked bar plot is shown for the (**A**) TIL validation population (*n*=873), (**B**) TIL-ICP$_{EH}$ sub-population (*n*=837), and (**C**) TIL-ICP$_{HR}$ sub-population (*n*=259), as defined in Figure 1.

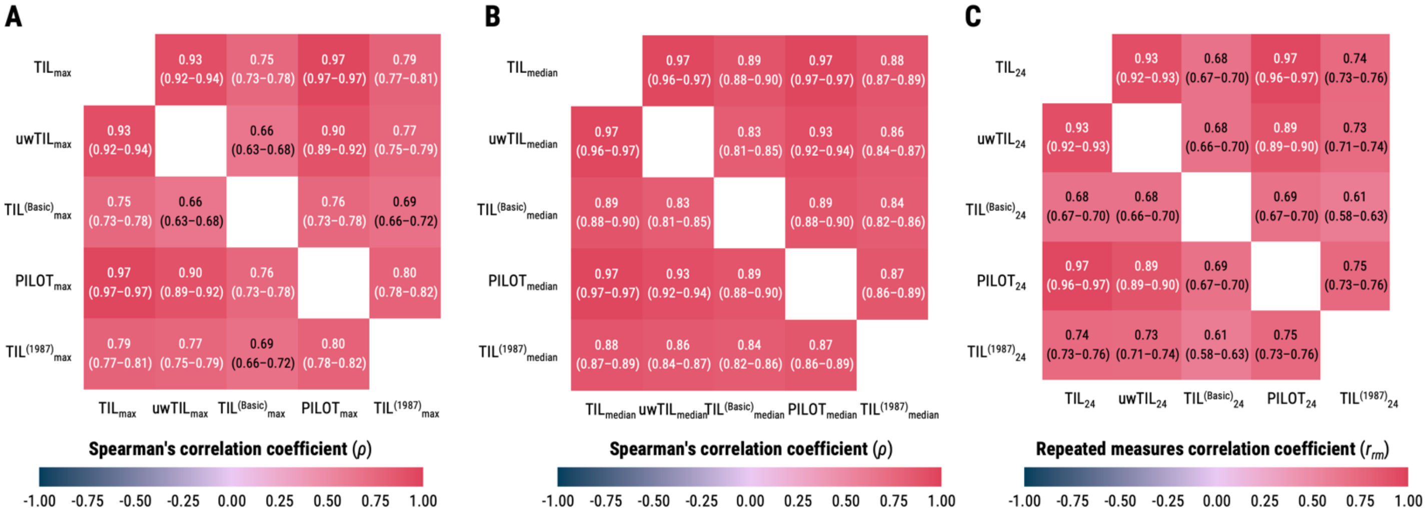


**Supplementary Figure S3. Correlation matrices between total scores of TIL and alternative scales.** Abbreviations: ICU=intensive care unit, PILOT=Paediatric Intensity Level of Therapy scale,[7] TIL=Therapy Intensity Level scale,[8,9] $TIL^{(1987)}$=original Therapy Intensity Level scale published in 1987,[6] $TIL^{(Basic)}$=condensed TIL scale,[8] uwTIL=unweighted TIL scale in which sub-item scores are replaced by the ascending rank index within the item. The numeric definition of each scale is listed in Table 1, and the calculation of daily (e.g., $TIL_{24}$), maximum (e.g., $TIL_{max}$), and median (e.g., $TIL_{median}$) scores are described in the Methods. The values in parentheses represent 95% confidence intervals derived from bootstrapping with 1,000 resamples of unique patients over 100 missing value imputations. (**A**) Spearman's correlation matrix between maximum total scores of TIL and alternative scales. (**B**) Spearman's correlation matrix between median total scores of TIL and alternative scales. (**C**) Repeated measures (i.e., within-individual) correlation matrix between daily total scores of TIL and alternative scales.

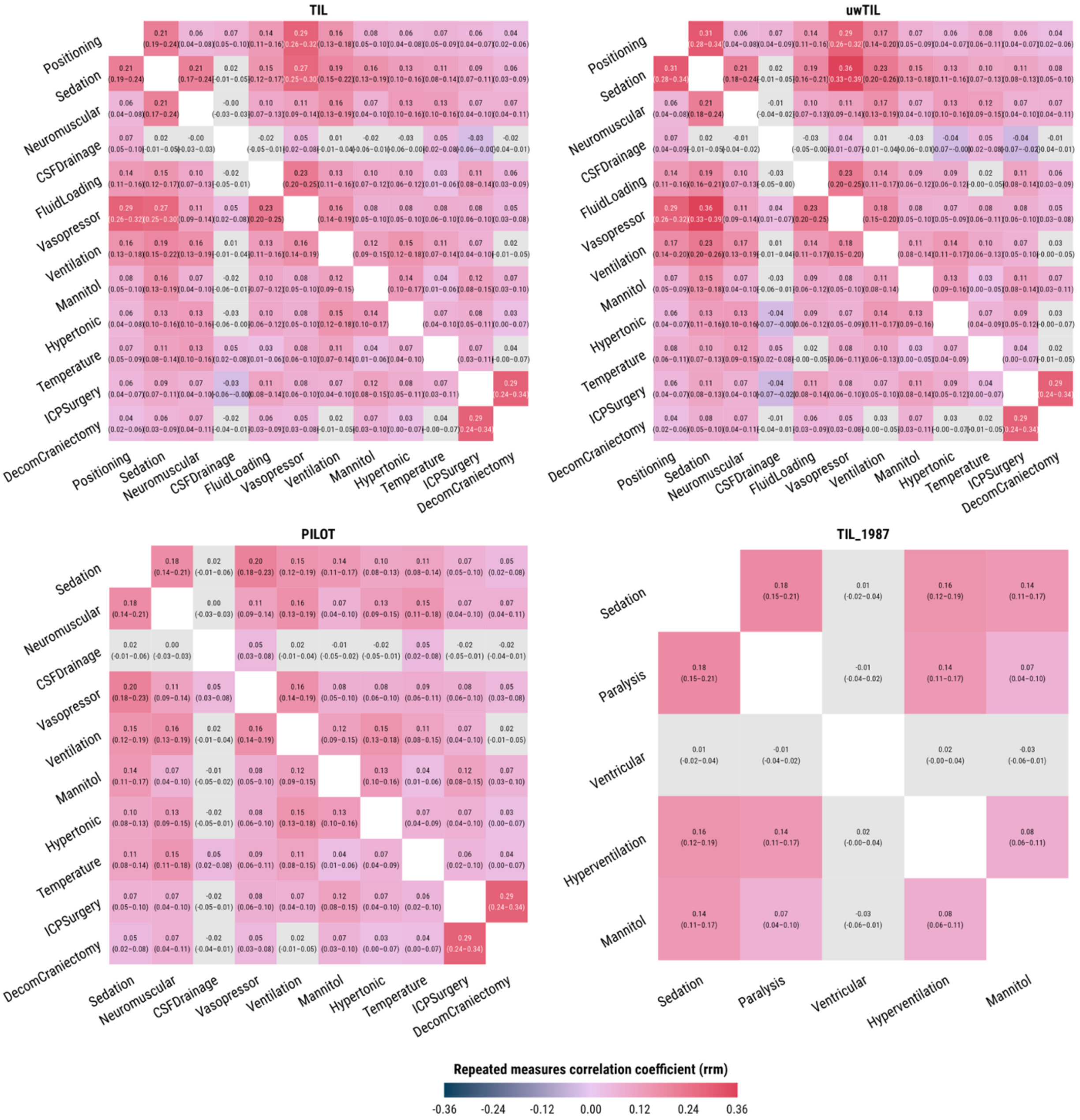


**Supplementary Figure S4. Inter-item correlation matrices for daily scores of TIL and alternative scales.** Abbreviations: ICU=intensive care unit, PILOT=Paediatric Intensity Level of Therapy scale,[7] TIL=Therapy Intensity Level scale,[8,9] TIL$^{(1987)}$=original Therapy Intensity Level scale published in 1987,[6] TIL$^{(Basic)}$=condensed TIL scale,[8] uwTIL=unweighted TIL scale in which sub-item scores are replaced by the ascending rank index within the item. The items and associated scores of each scale are described in Table 1. The values in parentheses represent 95% confidence intervals derived from bootstrapping with 1,000 resamples of unique patients over 100 missing value imputations, and light grey boxes designate statistically insignificant correlations.

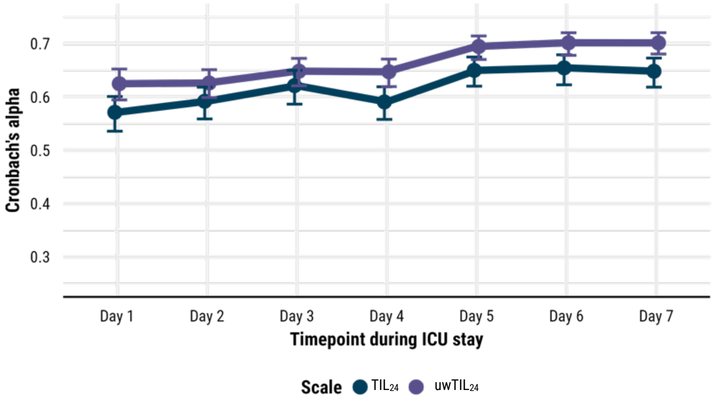


**Supplementary Figure S5. Internal reliability of TIL and uwTIL.** Abbreviations: ICU=intensive care unit, $TIL_{24}$=Therapy Intensity Level scale score of calendar day,[8,9] $uwTIL_{24}$=unweighted TIL scale score of calendar day in which sub-item scores are replaced by the ascending rank index within the item. The numeric definition of each scale is listed in Table 1, and the calculation of daily (e.g., $TIL_{24}$) scores are described in the Methods. The bars represent 95% confidence intervals derived from bootstrapping with 1,000 resamples of unique patients over 100 missing value imputations.

### Supplementary Table S1. Significant characteristic differences associated with longitudinal variable missingness.

| Validation population | Day in ICU | Variable | Nonmissing count | Missing count | Characteristic | Value | Nonmissing group | Missing group | p-value† |
|---|---|---|---|---|---|---|---|---|---|
| TIL | Day 1 | Physician concerns of ICP/CPP | 592 | 281 | Centre distribution* | | 46 | 35 | 0.000 |
| | | | 504 | 245 | Baseline functional prognosis | Pr(GOSE>4) | 37.2 (19.3–56.9) | 45.7 (22.2–64.6) | 0.004 |
| | | | 504 | 245 | Baseline functional prognosis | Pr(GOSE>5) | 19.6 (10.5–35.9) | 23.3 (9.5–39.0) | 0.039 |
| | | | 504 | 245 | Baseline functional prognosis | Pr(GOSE>6) | 11.4 (5.7–19.6) | 14.0 (6.3–21.9) | 0.013 |
| | | | 504 | 245 | Baseline functional prognosis | Pr(GOSE>7) | 4.4 (2.2–8.7) | 5.8 (2.5–10.2) | 0.005 |
| | | | 592 | 281 | TILmax | | 10.0 (7.0–15.0) | 10.0 (6.0–14.0) | 0.012 |
| | | | 592 | 281 | TILmedian | | 6.0 (3.0–10.0) | 5.0 (3.0–9.0) | 0.017 |
| | | | 592 | 260 | TIL24 | | 8.0 (5.0–12.0) | 6.0 (3.0–11.0) | 0.002 |
| TIL | Day 2 | TIL24 | 839 | 11 | Centre distribution* | | 51 | 11 | 0.000 |
| | | | 730 | 8 | Baseline functional prognosis | Pr(GOSE>6) | 12.4 (6.1–20.9) | 6.0 (3.3–9.7) | 0.021 |
| TIL | Day 2 | Physician concerns of ICP/CPP | 621 | 229 | Centre distribution* | | 45 | 37 | 0.000 |
| | | | 539 | 199 | Baseline functional prognosis | Pr(GOSE>4) | 38.2 (21.1–57.7) | 46.0 (20.7–63.1) | 0.038 |
| | | | 539 | 199 | Baseline functional prognosis | Pr(GOSE>6) | 11.9 (6.0–19.8) | 13.8 (6.3–22.4) | 0.033 |
| | | | 539 | 199 | Baseline functional prognosis | Pr(GOSE>7) | 4.6 (2.2–8.8) | 5.5 (2.4–10.5) | 0.020 |
| | | | 621 | 229 | TILmax | | 10.0 (7.0–15.0) | 9.0 (5.0–14.0) | 0.004 |
| | | | 621 | 229 | TILmedian | | 6.0 (3.0–10.0) | 5.0 (3.0–9.0) | 0.009 |
| | | | 621 | 218 | TIL24 | | 6.0 (4.0–11.0) | 6.0 (4.0–9.0) | 0.007 |
| TIL | Day 3 | TIL24 | 715 | 14 | Six-month GOSE | 1 | 164 (23%) | 12 (86%) | 0.000 |
| | | | 715 | 14 | Six-month GOSE | 2_or_3 | 180 (25%) | 1 (7%) | 0.000 |
| | | | 715 | | Six-month GOSE | 4 | 70 (10%) | | 0.000 |
| | | | 715 | 14 | Six-month GOSE | 5 | 121 (17%) | 1 (7%) | 0.000 |
| | | | 715 | | Six-month GOSE | 6 | 70 (10%) | | 0.000 |
| | | | 715 | | Six-month GOSE | 7 | 54 (8%) | | 0.000 |
| | | | 715 | | Six-month GOSE | 8 | 56 (8%) | | 0.000 |
| | | | 715 | 14 | Baseline functional prognosis | Pr(GOSE>1) | 85.5 (65.4–94.9) | 47.9 (24.0–69.0) | 0.003 |
| | | | 715 | 14 | Baseline functional prognosis | Pr(GOSE>3) | 55.2 (30.7–76.1) | 18.4 (7.7–43.2) | 0.003 |
| | | | 715 | 14 | Baseline functional prognosis | Pr(GOSE>4) | 40.6 (21.2–59.7) | 14.6 (5.0–30.7) | 0.007 |
| | | | 715 | 14 | Baseline functional prognosis | Pr(GOSE>5) | 21.6 (10.9–36.9) | 7.6 (2.5–18.0) | 0.007 |
| | | | 715 | 14 | Baseline functional prognosis | Pr(GOSE>6) | 12.5 (6.3–20.9) | 4.2 (1.7–8.0) | 0.021 |
| | | | 715 | 14 | Baseline functional prognosis | Pr(GOSE>7) | 5.1 (2.4–9.2) | 1.2 (0.5–4.5) | 0.001 |
| | | | 819 | 16 | TILmedian | | 5.0 (3.0–9.0) | 9.0 (6.0–11.8) | 0.032 |
| TIL | Day 3 | Physician concerns of ICP/CPP | 602 | 233 | Centre distribution* | | 44 | 37 | 0.000 |
| | | | 602 | 233 | TILmax | | 10.0 (7.0–15.0) | 10.0 (5.0–14.0) | 0.021 |
| | | | 602 | 217 | TIL24 | | 6.0 (4.0–10.0) | 5.0 (3.0–8.0) | 0.001 |
| TIL | Day 4 | TIL24 | 642 | 10 | Marshall CT | 1 | 15 (2%) | 2 (20%) | 0.011 |
| | | | 642 | 10 | Marshall CT | 2 | 249 (39%) | 3 (30%) | 0.011 |
| | | | 642 | 10 | Marshall CT | 3 | 80 (12%) | 2 (20%) | 0.011 |
| | | | 642 | | Marshall CT | 4 | 14 (2%) | | 0.011 |
| | | | 642 | 10 | Marshall CT | 5_or_6 | 284 (44%) | 3 (30%) | 0.011 |
| | | | 787 | 12 | TILmax | | 10.0 (6.0–15.0) | 8.5 (4.5–11.0) | 0.048 |
| TIL | Day 4 | Physician concerns of ICP/CPP | 569 | 230 | Centre distribution* | | 45 | 36 | 0.000 |
| | | | 493 | 200 | Baseline functional prognosis | Pr(GOSE>7) | 4.8 (2.5–8.8) | 5.7 (2.6–10.3) | 0.026 |
| | | | 569 | 230 | TILmax | | 10.0 (7.0–15.0) | 10.0 (5.2–14.0) | 0.004 |
| | | | 569 | 230 | TILmedian | | 6.0 (3.0–10.0) | 5.0 (3.0–8.0) | 0.001 |
| | | | 569 | 218 | TIL24 | | 6.0 (3.0–10.0) | 5.0 (3.0–8.0) | 0.002 |
| TIL | Day 5 | TIL24 | 663 | 12 | Six-month GOSE | 1 | 138 (21%) | 9 (75%) | 0.001 |
| | | | 663 | 12 | Six-month GOSE | 2_or_3 | 176 (27%) | 2 (17%) | 0.001 |
| | | | 663 | | Six-month GOSE | 4 | 69 (10%) | | 0.001 |
| | | | 663 | | Six-month GOSE | 5 | 119 (18%) | | 0.001 |
| | | | 663 | | Six-month GOSE | 6 | 66 (10%) | | 0.001 |

| Validation population | Day in ICU | Variable | Nonmissing count | Missing count | Characteristic | Value | Nonmissing group | Missing group | p-value† |
|---|---|---|---|---|---|---|---|---|---|
| | | | 663 | | Six-month GOSE | 7 | 49 (7%) | | 0.001 |
| | | | 663 | 12 | Six-month GOSE | 8 | 46 (7%) | 1 (8%) | 0.001 |
| | | | 663 | 12 | Baseline functional prognosis | Pr(GOSE>1) | 85.7 (66.1–95.1) | 63.2 (38.3–72.6) | 0.022 |
| | | | 663 | 12 | Baseline functional prognosis | Pr(GOSE>3) | 55.5 (32.3–76.1) | 29.8 (11.4–40.0) | 0.005 |
| | | | 663 | 12 | Baseline functional prognosis | Pr(GOSE>4) | 41.0 (22.7–59.6) | 23.5 (7.2–29.8) | 0.011 |
| | | | 663 | 12 | Baseline functional prognosis | Pr(GOSE>5) | 21.8 (11.2–37.0) | 11.6 (4.0–16.5) | 0.046 |
| TIL | Day 5 | Physician concerns of ICP/CPP | 551 | 227 | Centre distribution* | | 42 | 36 | 0.000 |
| | | | 551 | 227 | TILmedian | | 6.0 (3.8–10.0) | 5.0 (3.0–8.0) | 0.004 |
| | | | 551 | 210 | TIL24 | | 6.0 (3.0–10.0) | 5.0 (2.0–9.0) | 0.000 |
| TIL | Day 6 | TIL24 | 633 | 11 | Six-month GOSE | 1 | 127 (20%) | 8 (73%) | 0.003 |
| | | | 633 | 11 | Six-month GOSE | 2_or_3 | 175 (28%) | 2 (18%) | 0.003 |
| | | | 633 | | Six-month GOSE | 4 | 68 (11%) | | 0.003 |
| | | | 633 | | Six-month GOSE | 5 | 112 (18%) | | 0.003 |
| | | | 633 | | Six-month GOSE | 6 | 64 (10%) | | 0.003 |
| | | | 633 | | Six-month GOSE | 7 | 44 (7%) | | 0.003 |
| | | | 633 | 11 | Six-month GOSE | 8 | 43 (7%) | 1 (9%) | 0.003 |
| | | | 633 | 11 | Baseline functional prognosis | Pr(GOSE>3) | 55.5 (33.4–75.6) | 28.5 (17.3–40.4) | 0.011 |
| | | | 633 | 11 | Baseline functional prognosis | Pr(GOSE>4) | 41.1 (23.5–59.0) | 22.2 (13.5–29.8) | 0.021 |
| | | | 733 | 15 | TILmax | | 10.0 (7.0–15.0) | 8.0 (4.0–12.0) | 0.022 |
| TIL | Day 6 | Physician concerns of ICP/CPP | 527 | 221 | Centre distribution* | | 42 | 35 | 0.000 |
| | | | 527 | 221 | TILmax | | 11.0 (7.0–15.0) | 10.0 (6.0–14.0) | 0.005 |
| | | | 527 | 221 | TILmedian | | 6.0 (4.0–10.0) | 5.0 (3.0–8.0) | 0.000 |
| | | | 527 | 206 | TIL24 | | 5.0 (3.0–10.0) | 4.5 (2.0–8.0) | 0.003 |
| TIL | Day 7 | TIL24 | 709 | 17 | Centre distribution* | | 51 | 12 | 0.001 |
| TIL | Day 7 | Physician concerns of ICP/CPP | 504 | 222 | Centre distribution* | | 41 | 39 | 0.000 |
| | | | 504 | 222 | TILmedian | | 6.0 (4.0–10.0) | 5.0 (3.0–8.0) | 0.003 |
| | | | 504 | 205 | TIL24 | | 5.0 (2.0–9.0) | 4.0 (2.0–7.0) | 0.009 |
| TIL-ICPEH | Day 1 | TIL24 | 820 | 17 | TILmax | | 10.0 (6.0–15.0) | 6.0 (4.0–10.0) | 0.012 |
| | | | 820 | 17 | TILmedian | | 5.5 (3.0–10.0) | 4.5 (2.0–6.0) | 0.047 |
| TIL-ICPEH | Day 1 | ICP/CPP | 677 | 160 | Centre distribution* | | 50 | 40 | 0.000 |
| | | | 555 | 126 | Marshall CT | 1 | 11 (2%) | 5 (4%) | 0.013 |
| | | | 555 | 126 | Marshall CT | 2 | 187 (34%) | 61 (48%) | 0.013 |
| | | | 555 | 126 | Marshall CT | 3 | 75 (14%) | 14 (11%) | 0.013 |
| | | | 555 | 126 | Marshall CT | 4 | 14 (3%) | 2 (2%) | 0.013 |
| | | | 555 | 126 | Marshall CT | 5_or_6 | 268 (48%) | 44 (35%) | 0.013 |
| | | | 593 | 132 | Baseline functional prognosis | Pr(GOSE>3) | 50.9 (27.3–73.6) | 63.2 (31.5–80.4) | 0.008 |
| | | | 593 | 132 | Baseline functional prognosis | Pr(GOSE>4) | 37.0 (18.7–56.7) | 47.5 (22.7–65.0) | 0.008 |
| | | | 593 | 132 | Baseline functional prognosis | Pr(GOSE>5) | 19.3 (9.5–33.7) | 27.7 (14.1–42.7) | 0.001 |
| | | | 593 | 132 | Baseline functional prognosis | Pr(GOSE>6) | 11.4 (5.6–18.9) | 14.0 (7.4–23.8) | 0.004 |
| | | | 593 | 132 | Baseline functional prognosis | Pr(GOSE>7) | 4.5 (2.2–8.9) | 5.9 (2.9–10.1) | 0.020 |
| | | | 671 | 149 | TIL24 | | 8.0 (5.0–12.0) | 5.0 (2.0–9.0) | 0.000 |
| TIL-ICPEH | Day 1 | Physician concerns of ICP/CPP | 572 | 265 | Centre distribution* | | 45 | 35 | 0.000 |
| | | | 490 | 235 | Baseline functional prognosis | Pr(GOSE>4) | 36.6 (18.9–55.9) | 44.0 (21.0–63.1) | 0.008 |
| | | | 490 | 235 | Baseline functional prognosis | Pr(GOSE>6) | 11.2 (5.6–19.1) | 13.7 (6.1–21.5) | 0.017 |
| | | | 490 | 235 | Baseline functional prognosis | Pr(GOSE>7) | 4.4 (2.2–8.7) | 5.5 (2.4–10.2) | 0.008 |
| | | | 572 | 265 | TILmax | | 10.0 (7.0–15.0) | 10.0 (6.0–14.0) | 0.012 |
| | | | 572 | 265 | TILmedian | | 6.0 (3.0–10.0) | 5.0 (3.0–9.0) | 0.016 |
| | | | 572 | 248 | TIL24 | | 8.0 (5.0–12.0) | 6.0 (3.0–11.0) | 0.009 |
| TIL-ICPEH | Day 2 | TIL24 | 807 | 8 | Centre distribution* | | 50 | 8 | 0.000 |
| | | | 707 | 7 | Baseline functional prognosis | Pr(GOSE>6) | 12.1 (6.0–19.9) | 5.0 (3.2–8.2) | 0.038 |
| TIL-ICPEH | Day 2 | ICP/CPP | 780 | 35 | Centre distribution* | | 51 | 21 | 0.003 |

| Validation population | Day in ICU | Variable | Nonmissing count | Missing count | Characteristic | Value | Nonmissing group | Missing group | p-value† |
|---|---|---|---|---|---|---|---|---|---|
| | | | 688 | 26 | Baseline functional prognosis | Pr(GOSE>3) | 52.9 (29.5–74.6) | 73.4 (46.3–85.3) | 0.044 |
| | | | 688 | 26 | Baseline functional prognosis | Pr(GOSE>4) | 38.6 (19.8–57.8) | 56.3 (30.8–70.9) | 0.030 |
| | | | 688 | 26 | Baseline functional prognosis | Pr(GOSE>5) | 20.3 (10.0–35.3) | 40.3 (13.1–47.8) | 0.018 |
| | | | 688 | 26 | Baseline functional prognosis | Pr(GOSE>6) | 11.8 (5.9–19.4) | 21.4 (7.5–32.4) | 0.019 |
| | | | 688 | 26 | Baseline functional prognosis | Pr(GOSE>7) | 4.6 (2.2–8.9) | 8.3 (2.8–15.5) | 0.041 |
| TIL-ICPEH | Day 2 | Physician concerns of ICP/CPP | 603 | 212 | Centre distribution* | | 45 | 36 | 0.000 |
| | | | 525 | 189 | Baseline functional prognosis | Pr(GOSE>7) | 4.6 (2.2–8.7) | 5.3 (2.3–10.5) | 0.041 |
| | | | 603 | 212 | TILmax | | 10.0 (7.0–15.0) | 9.0 (5.8–14.0) | 0.009 |
| | | | 603 | 212 | TILmedian | | 6.0 (3.0–10.0) | 5.0 (3.0–8.0) | 0.010 |
| | | | 603 | 204 | TIL24 | | 6.0 (4.0–11.0) | 6.0 (4.0–9.0) | 0.008 |
| TIL-ICPEH | Day 3 | TIL24 | 693 | 13 | Six-month GOSE | 1 | 161 (23%) | 11 (85%) | 0.000 |
| | | | 693 | 13 | Six-month GOSE | 2_or_3 | 179 (26%) | 1 (8%) | 0.000 |
| | | | 693 | | Six-month GOSE | 4 | 66 (10%) | | 0.000 |
| | | | 693 | 13 | Six-month GOSE | 5 | 116 (17%) | 1 (8%) | 0.000 |
| | | | 693 | | Six-month GOSE | 6 | 67 (10%) | | 0.000 |
| | | | 693 | | Six-month GOSE | 7 | 51 (7%) | | 0.000 |
| | | | 693 | | Six-month GOSE | 8 | 53 (8%) | | 0.000 |
| | | | 693 | 13 | Baseline functional prognosis | Pr(GOSE>1) | 85.0 (64.5–94.9) | 40.0 (23.4–67.9) | 0.003 |
| | | | 693 | 13 | Baseline functional prognosis | Pr(GOSE>3) | 54.6 (30.3–75.0) | 12.4 (7.4–39.6) | 0.003 |
| | | | 693 | 13 | Baseline functional prognosis | Pr(GOSE>4) | 40.3 (21.0–58.8) | 9.5 (4.9–29.9) | 0.006 |
| | | | 693 | 13 | Baseline functional prognosis | Pr(GOSE>5) | 21.2 (10.6–36.3) | 7.2 (2.2–15.5) | 0.010 |
| | | | 693 | 13 | Baseline functional prognosis | Pr(GOSE>6) | 12.4 (6.1–20.0) | 3.5 (1.6–7.1) | 0.032 |
| | | | 693 | 13 | Baseline functional prognosis | Pr(GOSE>7) | 4.9 (2.4–9.1) | 1.1 (0.5–4.4) | 0.001 |
| | | | 788 | 13 | TILmedian | | 5.0 (3.0–9.0) | 9.0 (7.0–14.0) | 0.007 |
| TIL-ICPEH | Day 3 | ICP/CPP | 740 | 61 | Centre distribution* | | 48 | 32 | 0.000 |
| | | | 657 | 49 | Six-month GOSE | 1 | 166 (25%) | 6 (12%) | 0.034 |
| | | | 657 | 49 | Six-month GOSE | 2_or_3 | 169 (26%) | 11 (22%) | 0.034 |
| | | | 657 | 49 | Six-month GOSE | 4 | 64 (10%) | 2 (4%) | 0.034 |
| | | | 657 | 49 | Six-month GOSE | 5 | 108 (16%) | 9 (18%) | 0.034 |
| | | | 657 | 49 | Six-month GOSE | 6 | 60 (9%) | 7 (14%) | 0.034 |
| | | | 657 | 49 | Six-month GOSE | 7 | 45 (7%) | 6 (12%) | 0.034 |
| | | | 657 | 49 | Six-month GOSE | 8 | 45 (7%) | 8 (16%) | 0.034 |
| | | | 657 | 49 | Baseline functional prognosis | Pr(GOSE>4) | 38.7 (20.8–57.1) | 58.3 (18.7–72.0) | 0.026 |
| | | | 657 | 49 | Baseline functional prognosis | Pr(GOSE>5) | 20.2 (10.2–34.4) | 36.1 (10.6–48.2) | 0.010 |
| | | | 657 | 49 | Baseline functional prognosis | Pr(GOSE>6) | 11.7 (6.0–18.9) | 16.1 (6.6–27.1) | 0.017 |
| | | | 657 | 49 | Baseline functional prognosis | Pr(GOSE>7) | 4.6 (2.3–8.8) | 7.2 (2.7–12.9) | 0.048 |
| | | | 740 | 61 | TILmax | | 10.0 (7.0–15.0) | 7.0 (4.0–12.0) | 0.005 |
| | | | 740 | 61 | TILmedian | | 6.0 (4.0–10.0) | 3.0 (1.0–5.0) | 0.000 |
| | | | 729 | 59 | TIL24 | | 6.0 (4.0–10.0) | 3.0 (0.0–5.5) | 0.000 |
| TIL-ICPEH | Day 3 | Physician concerns of ICP/CPP | 582 | 219 | Centre distribution* | | 44 | 36 | 0.000 |
| | | | 582 | 219 | TILmax | | 10.0 (7.0–15.0) | 10.0 (6.0–14.0) | 0.020 |
| | | | 582 | 219 | TILmedian | | 6.0 (3.5–10.0) | 5.0 (3.0–8.2) | 0.037 |
| | | | 582 | 206 | TIL24 | | 6.0 (4.0–10.0) | 5.0 (3.0–8.0) | 0.000 |
| TIL-ICPEH | Day 4 | ICP/CPP | 667 | 98 | Centre distribution* | | 48 | 37 | 0.000 |
| | | | 667 | 98 | Age | | 46.0 (29.0–61.0) | 53.0 (34.2–64.8) | 0.030 |
| | | | 589 | 81 | Six-month GOSE | 1 | 136 (23%) | 10 (12%) | 0.003 |
| | | | 589 | 81 | Six-month GOSE | 2_or_3 | 162 (28%) | 18 (22%) | 0.003 |
| | | | 589 | 81 | Six-month GOSE | 4 | 62 (11%) | 4 (5%) | 0.003 |
| | | | 589 | 81 | Six-month GOSE | 5 | 100 (17%) | 16 (20%) | 0.003 |
| | | | 589 | 81 | Six-month GOSE | 6 | 54 (9%) | 11 (14%) | 0.003 |
| | | | 589 | 81 | Six-month GOSE | 7 | 39 (7%) | 9 (11%) | 0.003 |

| Validation population | Day in ICU | Variable | Nonmissing count | Missing count | Characteristic | Value | Nonmissing group | Missing group | p-value† |
|---|---|---|---|---|---|---|---|---|---|
| | | | 589 | 81 | Six-month GOSE | 8 | 36 (6%) | 13 (16%) | 0.003 |
| | | | 589 | 81 | Baseline functional prognosis | Pr(GOSE>3) | 53.4 (30.8–73.6) | 64.1 (33.6–82.3) | 0.023 |
| | | | 589 | 81 | Baseline functional prognosis | Pr(GOSE>4) | 39.7 (21.3–56.5) | 49.4 (26.1–71.5) | 0.004 |
| | | | 589 | 81 | Baseline functional prognosis | Pr(GOSE>5) | 20.5 (10.6–34.2) | 30.9 (14.0–48.2) | 0.001 |
| | | | 589 | 81 | Baseline functional prognosis | Pr(GOSE>6) | 11.7 (6.1–18.7) | 16.5 (9.3–25.7) | 0.001 |
| | | | 589 | 81 | Baseline functional prognosis | Pr(GOSE>7) | 4.6 (2.5–8.7) | 8.1 (2.7–12.6) | 0.003 |
| | | | 667 | 98 | TILmax | | 11.0 (7.0–15.0) | 6.0 (4.0–10.0) | 0.000 |
| | | | 667 | 98 | TILmedian | | 6.0 (4.0–10.0) | 2.0 (1.0–3.5) | 0.000 |
| | | | 661 | 96 | TIL24 | | 6.0 (4.0–10.0) | 1.0 (0.0–3.0) | 0.000 |
| TIL-ICPEH | Day 4 | Physician concerns of ICP/CPP | 550 | 215 | Centre distribution* | | 45 | 35 | 0.000 |
| | | | 550 | 215 | TILmax | | 10.0 (7.0–15.0) | 10.0 (6.0–14.0) | 0.009 |
| | | | 550 | 215 | TILmedian | | 6.0 (3.5–10.0) | 5.0 (3.0–8.0) | 0.001 |
| | | | 550 | 207 | TIL24 | | 6.0 (3.0–10.0) | 5.0 (3.0–8.0) | 0.003 |
| TIL-ICPEH | Day 5 | TIL24 | 642 | 10 | Six-month GOSE | 1 | 135 (21%) | 8 (80%) | 0.002 |
| | | | 642 | 10 | Six-month GOSE | 2_or_3 | 175 (27%) | 2 (20%) | 0.002 |
| | | | 642 | | Six-month GOSE | 4 | 65 (10%) | | 0.002 |
| | | | 642 | | Six-month GOSE | 5 | 114 (18%) | | 0.002 |
| | | | 642 | | Six-month GOSE | 6 | 63 (10%) | | 0.002 |
| | | | 642 | | Six-month GOSE | 7 | 46 (7%) | | 0.002 |
| | | | 642 | | Six-month GOSE | 8 | 44 (7%) | | 0.002 |
| | | | 642 | 10 | Baseline functional prognosis | Pr(GOSE>1) | 85.7 (65.6–94.9) | 60.4 (35.0–69.0) | 0.016 |
| | | | 642 | 10 | Baseline functional prognosis | Pr(GOSE>3) | 55.1 (31.3–75.0) | 29.0 (9.5–38.0) | 0.000 |
| | | | 642 | 10 | Baseline functional prognosis | Pr(GOSE>4) | 40.7 (22.4–58.7) | 21.6 (6.0–28.6) | 0.000 |
| | | | 642 | 10 | Baseline functional prognosis | Pr(GOSE>5) | 21.4 (11.0–36.3) | 10.3 (2.9–15.0) | 0.000 |
| | | | 642 | 10 | Baseline functional prognosis | Pr(GOSE>6) | 12.4 (6.4–20.0) | 5.0 (2.1–8.0) | 0.001 |
| | | | 642 | 10 | Baseline functional prognosis | Pr(GOSE>7) | 5.1 (2.5–9.2) | 2.3 (0.5–4.5) | 0.002 |
| TIL-ICPEH | Day 5 | ICP/CPP | 613 | 131 | Centre distribution* | | 48 | 40 | 0.002 |
| | | | 503 | 103 | Marshall CT | 1 | 12 (2%) | 3 (3%) | 0.027 |
| | | | 503 | 103 | Marshall CT | 2 | 177 (35%) | 53 (51%) | 0.027 |
| | | | 503 | 103 | Marshall CT | 3 | 69 (14%) | 9 (9%) | 0.027 |
| | | | 503 | 103 | Marshall CT | 4 | 11 (2%) | 3 (3%) | 0.027 |
| | | | 503 | 103 | Marshall CT | 5_or_6 | 234 (47%) | 35 (34%) | 0.027 |
| | | | 609 | 128 | Refractory intracranial hypertension | | 113 (19%) | 9 (7%) | 0.002 |
| | | | 539 | 113 | Six-month GOSE | 1 | 129 (24%) | 14 (12%) | 0.036 |
| | | | 539 | 113 | Six-month GOSE | 2_or_3 | 146 (27%) | 31 (27%) | 0.036 |
| | | | 539 | 113 | Six-month GOSE | 4 | 58 (11%) | 7 (6%) | 0.036 |
| | | | 539 | 113 | Six-month GOSE | 5 | 88 (16%) | 26 (23%) | 0.036 |
| | | | 539 | 113 | Six-month GOSE | 6 | 49 (9%) | 14 (12%) | 0.036 |
| | | | 539 | 113 | Six-month GOSE | 7 | 36 (7%) | 10 (9%) | 0.036 |
| | | | 539 | 113 | Six-month GOSE | 8 | 33 (6%) | 11 (10%) | 0.036 |
| | | | 539 | 113 | Baseline functional prognosis | Pr(GOSE>3) | 53.1 (31.1–73.0) | 61.8 (30.8–81.9) | 0.031 |
| | | | 539 | 113 | Baseline functional prognosis | Pr(GOSE>4) | 39.2 (21.2–56.4) | 44.9 (24.8–69.1) | 0.011 |
| | | | 539 | 113 | Baseline functional prognosis | Pr(GOSE>5) | 19.9 (10.5–33.4) | 26.8 (12.7–47.0) | 0.001 |
| | | | 539 | 113 | Baseline functional prognosis | Pr(GOSE>6) | 11.6 (6.1–18.5) | 16.6 (7.8–25.5) | 0.000 |
| | | | 539 | 113 | Baseline functional prognosis | Pr(GOSE>7) | 4.5 (2.3–8.5) | 7.3 (2.7–12.0) | 0.002 |
| | | | 613 | 131 | TILmax | | 11.0 (7.0–15.0) | 7.0 (4.5–10.5) | 0.000 |
| | | | 613 | 131 | TILmedian | | 6.0 (4.0–10.0) | 3.0 (1.0–4.0) | 0.000 |
| | | | 603 | 128 | TIL24 | | 6.0 (4.0–10.5) | 2.0 (1.0–3.2) | 0.000 |
| TIL-ICPEH | Day 5 | Physician concerns of ICP/CPP | 531 | 213 | Centre distribution* | | 42 | 35 | 0.000 |
| | | | 531 | 213 | TILmedian | | 6.0 (4.0–10.0) | 5.0 (3.0–8.0) | 0.009 |
| | | | 531 | 200 | TIL24 | | 6.0 (3.0–10.0) | 5.0 (2.0–9.0) | 0.001 |

| Validation population | Day in ICU | Variable | Nonmissing count | Missing count | Characteristic | Value | Nonmissing group | Missing group | p-value† |
|---|---|---|---|---|---|---|---|---|---|
| TIL-ICPEH | Day 6 | TIL24 | 612 | 9 | Six-month GOSE | 1 | 124 (20%) | 7 (78%) | 0.005 |
| | | | 612 | 9 | Six-month GOSE | 2_or_3 | 174 (28%) | 2 (22%) | 0.005 |
| | | | 612 | | Six-month GOSE | 4 | 64 (10%) | | 0.005 |
| | | | 612 | | Six-month GOSE | 5 | 107 (17%) | | 0.005 |
| | | | 612 | | Six-month GOSE | 6 | 61 (10%) | | 0.005 |
| | | | 612 | | Six-month GOSE | 7 | 41 (7%) | | 0.005 |
| | | | 612 | | Six-month GOSE | 8 | 41 (7%) | | 0.005 |
| | | | 612 | 9 | Baseline functional prognosis | Pr(GOSE>1) | 85.7 (66.4–94.8) | 66.8 (38.8–75.1) | 0.046 |
| | | | 612 | 9 | Baseline functional prognosis | Pr(GOSE>3) | 55.1 (32.6–74.8) | 26.9 (10.7–39.6) | 0.001 |
| | | | 612 | 9 | Baseline functional prognosis | Pr(GOSE>4) | 40.8 (22.8–57.8) | 18.8 (8.5–29.8) | 0.000 |
| | | | 612 | 9 | Baseline functional prognosis | Pr(GOSE>5) | 21.4 (11.2–35.6) | 11.0 (4.2–15.5) | 0.000 |
| | | | 612 | 9 | Baseline functional prognosis | Pr(GOSE>6) | 12.4 (6.4–19.6) | 6.5 (2.1–8.2) | 0.005 |
| | | | 612 | 9 | Baseline functional prognosis | Pr(GOSE>7) | 5.0 (2.5–8.9) | 2.6 (0.7–4.4) | 0.005 |
| TIL-ICPEH | Day 6 | ICP/CPP | 547 | 167 | Centre distribution* | | 48 | 39 | 0.000 |
| | | | 547 | 167 | Age | | 44.0 (28.0–59.5) | 51.0 (34.0–65.5) | 0.004 |
| | | | 543 | 164 | Refractory intracranial hypertension | | 110 (20%) | 10 (6%) | 0.000 |
| | | | 480 | 141 | Six-month GOSE | 1 | 106 (22%) | 25 (18%) | 0.028 |
| | | | 480 | 141 | Six-month GOSE | 2_or_3 | 137 (29%) | 39 (28%) | 0.028 |
| | | | 480 | 141 | Six-month GOSE | 4 | 57 (12%) | 7 (5%) | 0.028 |
| | | | 480 | 141 | Six-month GOSE | 5 | 73 (15%) | 34 (24%) | 0.028 |
| | | | 480 | 141 | Six-month GOSE | 6 | 45 (9%) | 16 (11%) | 0.028 |
| | | | 480 | 141 | Six-month GOSE | 7 | 34 (7%) | 7 (5%) | 0.028 |
| | | | 480 | 141 | Six-month GOSE | 8 | 28 (6%) | 13 (9%) | 0.028 |
| | | | 480 | 141 | Baseline functional prognosis | Pr(GOSE>6) | 12.2 (6.1–18.6) | 12.9 (7.3–21.6) | 0.044 |
| | | | 547 | 167 | TILmax | | 11.0 (8.0–16.0) | 7.0 (5.0–11.0) | 0.000 |
| | | | 547 | 167 | TILmedian | | 7.0 (4.0–10.0) | 3.0 (2.0–5.0) | 0.000 |
| | | | 537 | 166 | TIL24 | | 6.0 (4.0–10.0) | 2.0 (1.0–3.0) | 0.000 |
| TIL-ICPEH | Day 6 | Physician concerns of ICP/CPP | 509 | 205 | Centre distribution* | | 42 | 34 | 0.000 |
| | | | 509 | 205 | TILmax | | 11.0 (7.0–15.0) | 10.0 (6.0–14.0) | 0.021 |
| | | | 509 | 205 | TILmedian | | 6.0 (4.0–10.0) | 5.0 (3.0–8.0) | 0.002 |
| | | | 509 | 194 | TIL24 | | 5.0 (3.0–10.0) | 5.0 (2.0–8.0) | 0.006 |
| TIL-ICPEH | Day 7 | TIL24 | 679 | 13 | Centre distribution* | | 50 | 9 | 0.008 |
| TIL-ICPEH | Day 7 | ICP/CPP | 433 | 259 | Centre distribution* | | 44 | 43 | 0.000 |
| | | | 359 | 208 | Marshall CT | 1 | 7 (2%) | 7 (3%) | 0.015 |
| | | | 359 | 208 | Marshall CT | 2 | 119 (33%) | 96 (46%) | 0.015 |
| | | | 359 | 208 | Marshall CT | 3 | 53 (15%) | 19 (9%) | 0.015 |
| | | | 359 | 208 | Marshall CT | 4 | 9 (3%) | 4 (2%) | 0.015 |
| | | | 359 | 208 | Marshall CT | 5_or_6 | 171 (48%) | 82 (39%) | 0.015 |
| | | | 430 | 255 | Refractory intracranial hypertension | | 98 (23%) | 17 (7%) | 0.000 |
| | | | 380 | 223 | Six-month GOSE | 1 | 89 (23%) | 33 (15%) | 0.001 |
| | | | 380 | 223 | Six-month GOSE | 2_or_3 | 112 (29%) | 62 (28%) | 0.001 |
| | | | 380 | 223 | Six-month GOSE | 4 | 44 (12%) | 20 (9%) | 0.001 |
| | | | 380 | 223 | Six-month GOSE | 5 | 49 (13%) | 56 (25%) | 0.001 |
| | | | 380 | 223 | Six-month GOSE | 6 | 37 (10%) | 22 (10%) | 0.001 |
| | | | 380 | 223 | Six-month GOSE | 7 | 30 (8%) | 11 (5%) | 0.001 |
| | | | 380 | 223 | Six-month GOSE | 8 | 19 (5%) | 19 (9%) | 0.001 |
| | | | 380 | 223 | Baseline functional prognosis | Pr(GOSE>1) | 82.5 (63.8–93.4) | 90.1 (73.2–96.4) | 0.000 |
| | | | 380 | 223 | Baseline functional prognosis | Pr(GOSE>3) | 51.4 (29.3–72.4) | 60.0 (38.9–78.4) | 0.000 |
| | | | 380 | 223 | Baseline functional prognosis | Pr(GOSE>4) | 37.3 (19.3–55.1) | 43.9 (28.1–63.5) | 0.000 |
| | | | 380 | 223 | Baseline functional prognosis | Pr(GOSE>5) | 19.6 (9.8–33.2) | 23.7 (13.7–38.4) | 0.004 |
| | | | 380 | 223 | Baseline functional prognosis | Pr(GOSE>6) | 11.4 (5.6–18.3) | 13.3 (7.9–21.6) | 0.004 |

| Validation population | Day in ICU | Variable | Nonmissing count | Missing count | Characteristic | Value | Nonmissing group | Missing group | p-value[†] |
|---|---|---|---|---|---|---|---|---|---|
| | | | 380 | 223 | Baseline functional prognosis | Pr(GOSE>7) | 4.4 (2.2–8.2) | 5.8 (3.2–10.4) | 0.005 |
| | | | 433 | 259 | TILmax | | 12.0 (9.0–16.0) | 8.0 (5.0–12.0) | 0.000 |
| | | | 433 | 259 | TILmedian | | 7.0 (5.0–11.0) | 4.0 (2.0–6.0) | 0.000 |
| | | | 426 | 253 | TIL24 | | 6.0 (4.0–11.0) | 2.0 (1.0–4.0) | 0.000 |
| TIL-ICPEH | Day 7 | Physician concerns of ICP/CPP | 483 | 209 | Centre distribution* | | 41 | 38 | 0.000 |
| | | | 483 | 209 | TILmedian | | 6.0 (4.0–10.0) | 5.0 (3.0–8.0) | 0.013 |
| | | | 483 | 196 | TIL24 | | 5.0 (2.0–9.0) | 4.0 (2.0–7.0) | 0.022 |
| TIL-ICPHR | Day 1 | TIL24 | 256 | 3 | Centre distribution* | | 21 | 2 | 0.006 |
| | | | 256 | 3 | TILmax | | 10.0 (6.0–14.0) | 6.0 (5.5–7.0) | 0.023 |
| TIL-ICPHR | Day 1 | ICP/CPP | 21 | 238 | Centre distribution* | | 5 | 21 | 0.000 |
| | | | 21 | 238 | Age | | 64.0 (51.0–70.0) | 46.5 (30.0–61.0) | 0.008 |
| | | | 20 | 213 | Baseline functional prognosis | Pr(GOSE>3) | 34.4 (24.8–49.7) | 54.2 (35.8–72.5) | 0.009 |
| | | | 20 | 213 | Baseline functional prognosis | Pr(GOSE>4) | 25.5 (14.1–37.5) | 39.8 (23.2–55.1) | 0.008 |
| | | | 20 | 213 | Baseline functional prognosis | Pr(GOSE>5) | 13.3 (6.9–22.2) | 19.6 (10.6–31.5) | 0.011 |
| TIL-ICPHR | Day 1 | Physician concerns of ICP/CPP | 192 | 67 | Centre distribution* | | 17 | 13 | 0.000 |
| | | | 174 | 59 | Baseline functional prognosis | Pr(GOSE>6) | 10.1 (5.2–16.4) | 13.7 (8.7–21.5) | 0.007 |
| | | | 174 | 59 | Baseline functional prognosis | Pr(GOSE>7) | 4.4 (2.0–7.8) | 7.4 (3.3–12.0) | 0.001 |
| TIL-ICPHR | Day 2 | ICP/CPP | 168 | 89 | Centre distribution* | | 19 | 19 | 0.000 |
| | | | 168 | 89 | Age | | 51.0 (32.0–64.0) | 43.0 (27.0–60.0) | 0.018 |
| | | | 167 | 89 | TIL24 | | 6.0 (4.0–9.0) | 6.0 (5.0–11.0) | 0.048 |
| TIL-ICPHR | Day 2 | Physician concerns of ICP/CPP | 209 | 48 | Centre distribution* | | 16 | 16 | 0.000 |
| | | | 191 | 40 | Baseline functional prognosis | Pr(GOSE>6) | 10.5 (5.6–16.7) | 13.5 (7.3–22.2) | 0.045 |
| | | | 191 | 40 | Baseline functional prognosis | Pr(GOSE>7) | 4.8 (2.2–8.2) | 6.4 (3.1–12.7) | 0.043 |
| | | | 209 | 48 | TILmax | | 10.0 (6.0–15.0) | 9.0 (5.8–11.0) | 0.017 |
| | | | 209 | 47 | TIL24 | | 6.0 (5.0–10.0) | 5.0 (3.0–8.5) | 0.003 |
| TIL-ICPHR | Day 3 | TIL24 | 226 | 5 | Six-month GOSE | 1 | 47 (21%) | 5 (100%) | 0.007 |
| | | | 226 | | Six-month GOSE | 2_or_3 | 63 (28%) | | 0.007 |
| | | | 226 | | Six-month GOSE | 4 | 22 (10%) | | 0.007 |
| | | | 226 | | Six-month GOSE | 5 | 44 (19%) | | 0.007 |
| | | | 226 | | Six-month GOSE | 6 | 23 (10%) | | 0.007 |
| | | | 226 | | Six-month GOSE | 7 | 14 (6%) | | 0.007 |
| | | | 226 | | Six-month GOSE | 8 | 13 (6%) | | 0.007 |
| | | | 226 | 5 | Baseline functional prognosis | Pr(GOSE>3) | 53.7 (35.2–72.2) | 33.2 (12.4–39.6) | 0.040 |
| | | | 226 | 5 | Baseline functional prognosis | Pr(GOSE>4) | 38.9 (23.3–55.0) | 25.2 (7.9–29.9) | 0.022 |
| | | | 226 | 5 | Baseline functional prognosis | Pr(GOSE>5) | 19.6 (10.7–31.0) | 12.9 (4.6–18.9) | 0.029 |
| | | | 226 | 5 | Baseline functional prognosis | Pr(GOSE>6) | 11.7 (6.0–17.5) | 7.1 (3.5–8.3) | 0.017 |
| | | | 226 | 5 | Baseline functional prognosis | Pr(GOSE>7) | 5.5 (2.3–8.6) | 3.8 (1.2–4.4) | 0.009 |
| | | | 251 | 5 | TILmedian | | 5.0 (3.2–10.0) | 9.0 (8.5–9.5) | 0.018 |
| TIL-ICPHR | Day 3 | ICP/CPP | 242 | 14 | Sex | F | 47 (19%) | 7 (50%) | 0.017 |
| | | | 237 | 14 | TIL24 | | 6.0 (4.0–9.0) | 3.0 (2.2–6.5) | 0.024 |
| TIL-ICPHR | Day 3 | Physician concerns of ICP/CPP | 203 | 53 | Centre distribution* | | 17 | 16 | 0.000 |
| | | | 185 | 46 | Baseline functional prognosis | Pr(GOSE>6) | 10.3 (5.5–16.5) | 13.6 (7.9–23.6) | 0.017 |
| | | | 185 | 46 | Baseline functional prognosis | Pr(GOSE>7) | 4.6 (2.2–8.1) | 6.8 (3.7–13.0) | 0.015 |
| TIL-ICPHR | Day 4 | TIL24 | 219 | 5 | Baseline functional prognosis | Pr(GOSE>3) | 53.9 (35.7–72.4) | 26.9 (12.4–33.2) | 0.008 |
| | | | 219 | 5 | Baseline functional prognosis | Pr(GOSE>4) | 39.2 (24.4–55.0) | 18.5 (7.9–25.2) | 0.008 |
| | | | 219 | 5 | Baseline functional prognosis | Pr(GOSE>5) | 20.2 (11.7–31.2) | 7.6 (4.6–12.9) | 0.007 |
| | | | 219 | 5 | Baseline functional prognosis | Pr(GOSE>6) | 11.9 (6.1–17.6) | 3.5 (3.5–8.3) | 0.012 |
| | | | 219 | 5 | Baseline functional prognosis | Pr(GOSE>7) | 5.6 (2.5–8.7) | 1.2 (0.8–4.4) | 0.006 |
| TIL-ICPHR | Day 4 | ICP/CPP | 221 | 28 | Centre distribution* | | 21 | 13 | 0.016 |
| | | | 221 | 28 | Sex | F | 41 (19%) | 11 (39%) | 0.022 |
| | | | 199 | 25 | Six-month GOSE | 1 | 41 (21%) | 5 (20%) | 0.043 |

| Validation population | Day in ICU | Variable | Nonmissing count | Missing count | Characteristic | Value | Nonmissing group | Missing group | p-value† |
|---|---|---|---|---|---|---|---|---|---|
| | | | 199 | 25 | Six-month GOSE | 2_or_3 | 57 (29%) | 6 (24%) | 0.043 |
| | | | 199 | | Six-month GOSE | 4 | 22 (11%) | | 0.043 |
| | | | 199 | 25 | Six-month GOSE | 5 | 38 (19%) | 5 (20%) | 0.043 |
| | | | 199 | 25 | Six-month GOSE | 6 | 21 (11%) | 2 (8%) | 0.043 |
| | | | 199 | 25 | Six-month GOSE | 7 | 12 (6%) | 2 (8%) | 0.043 |
| | | | 199 | 25 | Six-month GOSE | 8 | 8 (4%) | 5 (20%) | 0.043 |
| | | | 221 | 28 | TILmax | | 10.0 (7.0–14.0) | 5.0 (4.0–8.2) | 0.009 |
| | | | 221 | 28 | TILmedian | | 6.0 (4.0–10.0) | 2.0 (1.0–4.2) | 0.001 |
| | | | 217 | 27 | TIL24 | | 6.0 (4.0–10.0) | 2.0 (0.5–5.0) | 0.020 |
| TIL-ICPHR | Day 4 | Physician concerns of ICP/CPP | 195 | 54 | Centre distribution* | | 17 | 14 | 0.000 |
| TIL-ICPHR | Day 5 | TIL24 | 215 | 6 | Six-month GOSE | 1 | 41 (19%) | 5 (83%) | 0.020 |
| | | | 215 | 6 | Six-month GOSE | 2_or_3 | 61 (28%) | 1 (17%) | 0.020 |
| | | | 215 | | Six-month GOSE | 4 | 22 (10%) | | 0.020 |
| | | | 215 | | Six-month GOSE | 5 | 43 (20%) | | 0.020 |
| | | | 215 | | Six-month GOSE | 6 | 23 (11%) | | 0.020 |
| | | | 215 | | Six-month GOSE | 7 | 14 (7%) | | 0.020 |
| | | | 215 | | Six-month GOSE | 8 | 11 (5%) | | 0.020 |
| | | | 215 | 6 | Baseline functional prognosis | Pr(GOSE>3) | 53.9 (35.7–72.4) | 19.7 (9.5–31.6) | 0.003 |
| | | | 215 | 6 | Baseline functional prognosis | Pr(GOSE>4) | 39.2 (24.8–55.0) | 13.2 (5.6–23.6) | 0.002 |
| | | | 215 | 6 | Baseline functional prognosis | Pr(GOSE>5) | 20.2 (11.7–31.2) | 6.1 (2.8–11.6) | 0.002 |
| | | | 215 | 6 | Baseline functional prognosis | Pr(GOSE>6) | 11.9 (6.1–17.6) | 3.5 (2.1–7.1) | 0.003 |
| | | | 215 | 6 | Baseline functional prognosis | Pr(GOSE>7) | 5.6 (2.5–8.7) | 1.0 (0.5–3.6) | 0.001 |
| TIL-ICPHR | Day 5 | ICP/CPP | 193 | 53 | Centre distribution* | | 20 | 17 | 0.001 |
| | | | 193 | 53 | TILmax | | 11.0 (7.0–15.0) | 7.0 (5.0–9.0) | 0.000 |
| | | | 193 | 53 | TILmedian | | 6.0 (4.0–10.0) | 3.0 (2.0–5.0) | 0.000 |
| | | | 188 | 51 | TIL24 | | 6.0 (3.8–11.0) | 3.0 (1.0–4.0) | 0.000 |
| TIL-ICPHR | Day 5 | Physician concerns of ICP/CPP | 191 | 55 | Centre distribution* | | 17 | 14 | 0.000 |
| TIL-ICPHR | Day 6 | TIL24 | 204 | 4 | Baseline functional prognosis | Pr(GOSE>3) | 53.3 (35.7–70.8) | 17.8 (6.7–30.1) | 0.032 |
| | | | 204 | 4 | Baseline functional prognosis | Pr(GOSE>4) | 38.9 (24.9–54.6) | 11.7 (3.9–21.4) | 0.028 |
| | | | 204 | 4 | Baseline functional prognosis | Pr(GOSE>5) | 19.6 (11.8–29.9) | 4.9 (1.8–10.6) | 0.033 |
| | | | 204 | 4 | Baseline functional prognosis | Pr(GOSE>6) | 11.5 (6.1–16.8) | 2.6 (1.2–5.7) | 0.042 |
| | | | 204 | 4 | Baseline functional prognosis | Pr(GOSE>7) | 5.4 (2.5–8.4) | 0.6 (0.4–1.7) | 0.012 |
| TIL-ICPHR | Day 6 | ICP/CPP | 170 | 63 | Centre distribution* | | 20 | 15 | 0.000 |
| | | | 170 | 63 | Refractory intracranial hypertension | | 35 (21%) | 5 (8%) | 0.038 |
| | | | 170 | 63 | TILmax | | 11.0 (8.0–16.0) | 7.0 (5.0–10.0) | 0.000 |
| | | | 170 | 63 | TILmedian | | 6.5 (4.0–10.0) | 4.0 (3.0–6.0) | 0.000 |
| | | | 168 | 60 | TIL24 | | 6.0 (4.0–10.0) | 3.0 (1.0–6.0) | 0.000 |
| TIL-ICPHR | Day 6 | Physician concerns of ICP/CPP | 183 | 50 | Centre distribution* | | 17 | 16 | 0.000 |
| TIL-ICPHR | Day 7 | ICP/CPP | 130 | 96 | Centre distribution* | | 20 | 18 | 0.013 |
| | | | 130 | 96 | Refractory intracranial hypertension | | 28 (22%) | 10 (10%) | 0.042 |
| | | | 117 | 85 | Baseline functional prognosis | Pr(GOSE>1) | 82.5 (63.8–94.0) | 85.9 (70.3–94.0) | 0.042 |
| | | | 130 | 96 | TILmax | | 12.0 (8.0–16.0) | 8.0 (5.0–11.0) | 0.000 |
| | | | 130 | 96 | TILmedian | | 7.0 (4.1–11.0) | 4.5 (3.0–8.0) | 0.000 |
| | | | 128 | 93 | TIL24 | | 6.0 (4.0–10.0) | 3.0 (1.0–6.0) | 0.000 |
| TIL-ICPHR | Day 7 | Physician concerns of ICP/CPP | 171 | 55 | Centre distribution* | | 17 | 16 | 0.000 |

Abbreviations: Baseline GCS=Glasgow Coma Scale at ICU admission, from 3 to 15, GOS=Glasgow Outcome Scale, ICP=intracranial pressure, $ICP_{EH}$=end-hour ICP, $ICP_{HR}$=high-resolution ICP, Marshall CT=Marshall computerised tomography classification, Pr(GOSE>•)="probability of GOSE greater than • at six months post-injury" as previously calculated from the first 24 hours of admission,[27] TIL=Therapy Intensity Level scale, $TIL_{24}$=TIL score of calendar day in ICU, $TIL_{max}$=maximum $TIL_{24}$ over first week of ICU stay, $TIL_{median}$=median $TIL_{24}$ over first week of ICU stay. Data are median (IQR) for numeric characteristics and *n* (% of column group) for categorical characteristics, unless otherwise indicated.

*The values for centre distribution represent the number of unique centres for the non-missing value and missing value cohorts, respectively. For statistical testing, centre affiliation was treated as a categorical variable with 52 possible values.

†*p*-values, comparing patients in non-missing value group to those in the missing value group, are derived from with Welch's *t*-test for numeric variables and $\chi^2$ contingency table test for categorical variables.

**Supplementary Table S2. Study population representation of each TIL item and sub-item.**

| ICP-treatment modality | Item | TIL validation population | | |
|---|---|---|---|---|
| | Sub-item | Overall (*n*=873, 52 centres) | TIL-$ICP_{EH}$ (*n*=832, 51 centres) | TIL-$ICP_{HR}$ (*n*=255, 21 centres) |
| Positioning | Head elevation for ICP control or nursed flat (180°) for CPP management* | 827 (94.7%) | 793 (95.3%) | 245 (96.1%) |
| Sedation and neuromuscular blockade | Sedation* | 851 (97.5%) | 819 (98.4%) | 253 (99.2%) |
| | Low dose sedation (as required for mechanical ventilation) | 592 (67.8%) | 579 (69.6%) | 198 (77.6%) |
| | Higher dose sedation for ICP control (but not aiming for burst suppression) | 479 (54.9%) | 464 (55.8%) | 148 (58.0%) |
| | High dose propofol or barbiturates for ICP control (metabolic suppression) | 309 (35.4%) | 291 (35.0%) | 64 (25.1%) |
| | Neuromuscular blockade (paralysis)* | 305 (34.9%) | 291 (35.0%) | 78 (30.6%) |
| CSF drainage | CSF drainage volume* | 273 (31.3%) | 256 (30.8%) | 66 (25.9%) |
| | Low (<120 ml/day) | 208 (23.8%) | 197 (23.7%) | 50 (19.6%) |
| | High (≥120 ml/day) | 186 (21.3%) | 174 (20.9%) | 44 (17.3%) |
| CPP management | Fluid loading for maintenance of cerebral perfusion* | 517 (59.2%) | 503 (60.5%) | 172 (67.5%) |
| | Vasopressor therapy required for management of cerebral perfusion* | 738 (84.5%) | 716 (86.1%) | 220 (86.3%) |
| Ventilatory management | Hypocapnia for ICP control ($P_aCO_2$ in mmHg)* | 561 (64.3%) | 540 (64.9%) | 191 (74.9%) |
| | Mild ($35 \leq P_aCO_2 < 40$) | 496 (56.8%) | 478 (57.5%) | 177 (69.4%) |
| | Moderate ($30 \leq P_aCO_2 < 35$) | 214 (24.5%) | 208 (25.0%) | 74 (29.0%) |
| | Intensive ($P_aCO_2 < 30$) | 57 (6.5%) | 57 (6.9%) | 17 (6.7%) |
| Hyperosmolar therapy | Mannitol* | 238 (27.3%) | 232 (27.9%) | 69 (27.1%) |
| | ≤2g/kg/24h | 224 (25.7%) | 218 (26.2%) | 67 (26.3%) |
| | >2g/kg/24h | 43 (4.9%) | 42 (5.0%) | 5 (2.0%) |
| | Hypertonic saline* | 350 (40.1%) | 341 (41.0%) | 113 (44.3%) |
| | ≤0.3g/kg/24h | 312 (35.7%) | 306 (36.8%) | 105 (41.2%) |
| | >0.3g/kg/24h | 115 (13.2%) | 111 (13.3%) | 35 (13.7%) |
| Temperature control | Temperature control* | 575 (65.9%) | 555 (66.7%) | 190 (74.5%) |
| | Fever control (>38 or spontaneous <34.5) | 484 (55.4%) | 465 (55.9%) | 163 (63.9%) |
| | Cooling for ICP control (≥35) | 128 (14.7%) | 125 (15.0%) | 50 (19.6%) |
| | Hypothermia (<35) | 94 (10.8%) | 93 (11.2%) | 22 (8.6%) |
| Surgery for intracranial hypertension | Intracranial operation for progressive mass lesion, NOT scheduled on admission* | 147 (16.8%) | 144 (17.3%) | 52 (20.4%) |
| | Decompressive craniectomy* | 196 (22.5%) | 189 (22.7%) | 56 (22.0%) |

Abbreviations: CPP=cerebral perfusion pressure, CSF=cerebrospinal fluid, ICP=intracranial pressure, $ICP_{EH}$=end-hour ICP, $ICP_{HR}$=high-resolution ICP, $P_aCO_2$=partial pressure of carbon dioxide in arterial blood, PILOT=Paediatric Intensity Level of Therapy scale,[7] T=body temperature

in degrees Celsius, TIL=Therapy Intensity Level scale,[8,9] $TIL^{(1987)}$=original Therapy Intensity Level scale published in 1987,[6] $TIL^{(Basic)}$=condensed TIL scale,[8] uwTIL=unweighted TIL scale in which sub-item scores are replaced by the ascending rank index within the item.
Data are number of unique patients (% of column population) who received the item or sub-item therapy during the first week of their ICU stays.
*Data in these rows represent the number of unique patients (% of column population) who received any of the sub-item therapies for a given item during the first week of their ICU stays.

**Supplementary Table S3. Performance of refractory intracranial hypertension status detection at each threshold of $TIL_{max}$, $TIL_{median}$, $TIL^{(Basic)}_{max}$, and $TIL^{(Basic)}_{median}$.**

| **Scale summary metric** | **Threshold** (≥) | **Sensitivity** (%) | **Specificity** (%) | **Youden's J** (%) |
|---|---|---|---|---|
| $TIL_{max}$ | 0 | 100 (100–100) | 0 (0–0) | 0 (0–0) |
| | 1 | 100 (99–100) | 0 (0–1) | 0 (-1–1) |
| | 2 | 99 (98–100) | 1 (0–1) | -0 (-2–1) |
| | 3 | 99 (98–100) | 3 (2–4) | 2 (0–4) |
| | 4 | 99 (98–100) | 8 (6–9) | 7 (4–9) |
| | 5 | 98 (97–100) | 15 (13–17) | 13 (10–16) |
| | 6 | 96 (94–98) | 23 (20–25) | 19 (15–22) |
| | 7 | 94 (92–98) | 31 (28–33) | 25 (21–29) |
| | 8 | 94 (91–97) | 39 (36–42) | 33 (29–37) |
| | 9 | 91 (88–95) | 45 (42–48) | 36 (32–40) |
| | 10 | 88 (84–92) | 53 (50–56) | 41 (36–46) |
| | 11 | 83 (78–87) | 61 (58–64) | 43 (38–49) |
| | 12 | 77 (72–82) | 67 (64–70) | 43 (38–49) |
| | 13 | 73 (68–78) | 73 (71–76) | 46 (41–53) |
| | **14** | **68 (62–74)** | **79 (77–81)** | **47 (41–53)** |
| | 15 | 64 (58–70) | 83 (81–85) | 47 (40–53) |
| | 16 | 56 (50–62) | 86 (84–88) | 42 (36–48) |
| | 17 | 50 (44–56) | 90 (88–91) | 39 (33–46) |
| | 18 | 44 (38–50) | 93 (91–94) | 37 (31–43) |
| | 19 | 38 (32–44) | 94 (93–96) | 32 (26–38) |
| | 20 | 32 (27–38) | 96 (95–97) | 29 (23–34) |
| | 21 | 27 (22–33) | 97 (96–98) | 24 (19–30) |
| | 22 | 24 (19–29) | 98 (97–99) | 22 (17–27) |
| | 23 | 21 (16–26) | 98 (98–99) | 19 (14–24) |
| | 24 | 14 (10–18) | 99 (98–99) | 13 (9–17) |
| | 25 | 10 (6–14) | 99 (99–100) | 9 (6–13) |
| | 26 | 7 (4–10) | 100 (99–100) | 6 (3–9) |
| | 27 | 5 (2–7) | 100 (100–100) | 4 (2–7) |
| | 28 | 3 (1–4) | 100 (100–100) | 3 (1–4) |
| | 29 | 2 (0–3) | 100 (100–100) | 2 (0–3) |
| | 30 | 1 (0–2) | 100 (100–100) | 1 (0–2) |
| | 31 | 1 (0–1) | 100 (100–100) | 1 (0–1) |
| $TIL_{median}$ | 0 | 100 (100–100) | 0 (0–0) | 0 (0–0) |
| | 0.5 | 99 (98–100) | 4 (3–5) | 4 (2–5) |
| | 1 | 99 (98–100) | 5 (3–6) | 4 (2–5) |
| | 1.5 | 98 (97–100) | 10 (8–11) | 8 (5–10) |
| | 2 | 98 (97–100) | 10 (8–12) | 8 (6–11) |
| | 2.5 | 97 (95–99) | 18 (16–20) | 15 (12–18) |
| | 3 | 97 (95–99) | 18 (16–21) | 15 (12–18) |
| | 3.5 | 95 (93–98) | 32 (29–34) | 27 (24–31) |

| **Scale summary metric** | **Threshold** (≥) | **Sensitivity** (%) | **Specificity** (%) | **Youden's J** (%) |
|---|---|---|---|---|
| | 4 | 95 (93–98) | 32 (30–35) | 28 (24–32) |
| | 4.5 | 92 (89–96) | 45 (42–48) | 37 (33–42) |
| | 5 | 92 (89–95) | 45 (42–48) | 37 (33–42) |
| | 5.5 | 88 (84–92) | 59 (56–62) | 47 (42–52) |
| | 6 | 88 (84–92) | 59 (56–62) | 47 (43–52) |
| | 6.5 | 84 (81–88) | 68 (66–69) | 52 (49–56) |
| | 7 | 84 (80–89) | 67 (65–70) | 52 (46–57) |
| | **7.5** | **81 (77–87)** | **72 (70–75)** | **54 (48–60)** |
| | 8 | 81 (76–86) | 72 (70–75) | 53 (47–59) |
| | 8.5 | 73 (68–79) | 79 (77–80) | 52 (47–58) |
| | 9 | 73 (68–79) | 78 (76–81) | 52 (46–58) |
| | 9.5 | 65 (58–72) | 82 (81–84) | 48 (41–55) |
| | 10 | 65 (59–71) | 82 (80–85) | 47 (41–54) |
| | 10.5 | 59 (53–65) | 87 (85–89) | 46 (40–53) |
| | 11 | 59 (53–65) | 87 (86–89) | 47 (40–53) |
| | 11.5 | 52 (46–57) | 91 (90–93) | 43 (36–48) |
| | 12 | 52 (46–58) | 91 (90–93) | 43 (36–49) |
| | 12.5 | 44 (38–50) | 94 (93–96) | 38 (31–44) |
| | 13 | 43 (37–49) | 94 (93–96) | 37 (31–43) |
| | 13.5 | 36 (30–41) | 96 (95–97) | 32 (25–37) |
| | 14 | 35 (29–40) | 96 (95–97) | 31 (25–36) |
| | 14.5 | 31 (26–37) | 97 (96–98) | 28 (23–34) |
| | 15 | 31 (25–36) | 97 (96–98) | 28 (22–34) |
| | 15.5 | 24 (19–29) | 98 (98–99) | 22 (17–27) |
| | 16 | 23 (18–28) | 98 (98–99) | 22 (16–27) |
| | 16.5 | 20 (15–23) | 99 (99–100) | 19 (15–23) |
| | 17 | 19 (14–24) | 99 (99–100) | 18 (14–23) |
| | 17.5 | 13 (9–18) | 100 (99–100) | 13 (9–17) |
| | 18 | 13 (9–18) | 100 (100–100) | 13 (9–17) |
| | 18.5 | 9 (7–11) | 100 (100–100) | 9 (7–11) |
| | 19 | 8 (4–11) | 100 (100–100) | 7 (4–11) |
| | 19.5 | 7 (6–9) | 100 (100–100) | 7 (6–9) |
| | 20 | 6 (3–8) | 100 (100–100) | 6 (3–8) |
| | 20.5 | 6 (2–8) | 100 (100–100) | 5 (2–8) |
| | 21 | 5 (1–7) | 100 (100–100) | 5 (1–7) |
| | 21.5 | 6 (6–6) | 100 (100–100) | 6 (6–6) |
| | 22 | 5 (2–7) | 100 (100–100) | 5 (2–7) |
| | 22.5 | 3 (1–5) | 100 (100–100) | 3 (1–4) |
| | 23 | 3 (1–5) | 100 (100–100) | 3 (1–5) |
| | 24 | 1 (0–2) | 100 (100–100) | 1 (0–2) |
| | 25 | 0 (0–0) | 100 (100–100) | 0 (0–0) |
| | 27 | 1 (1–1) | 100 (100–100) | 1 (1–1) |

| **Scale summary metric** | **Threshold** (≥) | **Sensitivity** (%) | **Specificity** (%) | **Youden's J** (%) |
|---|---|---|---|---|
| | 28 | 0 (0–0) | 100 (100–100) | 0 (0–0) |
| $TIL^{(Basic)}_{max}$ | 0 | 100 (100–100) | 0 (0–0) | 0 (0–0) |
| | 1 | 100 (99–100) | 0 (0–1) | 0 (-1–1) |
| | 2 | 99 (98–100) | 3 (2–4) | 2 (0–3) |
| | 3 | 94 (91–97) | 32 (29–35) | 26 (22–30) |
| | **4** | **85 (81–89)** | **49 (45–52)** | **33 (28–39)** |
| | 5 | 0 (0–0) | 100 (100–100) | 0 (0–0) |
| $TIL^{(Basic)}_{median}$ | 0 | 100 (100–100) | 0 (0–0) | 0 (0–0) |
| | 0.5 | 99 (98–100) | 5 (4–6) | 5 (3–6) |
| | 1 | 99 (98–100) | 6 (4–7) | 5 (3–7) |
| | 1.5 | 98 (97–100) | 16 (14–18) | 15 (12–17) |
| | 2 | 98 (97–100) | 18 (15–20) | 16 (13–18) |
| | 2.5 | 80 (75–85) | 67 (64–69) | 47 (41–52) |
| | 3 | 79 (74–84) | 67 (65–70) | 46 (41–52) |
| | **3.5** | **67 (61–72)** | **81 (79–84)** | **48 (42–54)** |
| | 4 | 65 (60–71) | 81 (79–84) | 47 (41–52) |
| | 5 | 0 (0–0) | 100 (100–100) | 0 (0–0) |

Abbreviations: ICP=intracranial pressure, ICU=intensive care unit, TIL=Therapy Intensity Level scale,[8,9] $TIL^{(Basic)}$=condensed TIL scale.[8] The numeric definition of each scale is listed in Table 1, and the calculation of maximum (e.g., $TIL_{max}$) and median ($TIL_{median}$) scores is described in the Methods. The 95% confidence intervals of performance metrics were derived from bootstrapping with 1,000 resamples of unique patients over 100 missing value imputations. Refractory intracranial hypertension was defined as recurrent, sustained (i.e., of at least ten minutes) increases of ICP above 20 mmHg despite medical ICP management during ICU stay. This information was recorded by attending physicians in patient discharge summaries. The rows in bold designate the thresholds which maximise Youden's J statistic for each scale summary metric.

**Supplementary Table S4. Performance of detecting day of surgical ICP control at each threshold of $TIL_{24}$.**

| **$TIL_{24}$ threshold** (≥) | **Sensitivity** (%) | **Specificity** (%) | **Youden's J** (%) |
|---|---|---|---|
| 0 | 100% (100–100%) | 0% (0–0%) | 0% (0–0%) |
| 1 | 100% (100–100%) | 7% (6–7%) | 7% (6–7%) |
| 2 | 100% (100–100%) | 12% (11–13%) | 12% (11–13%) |
| 3 | 100% (100–100%) | 21% (19–22%) | 21% (19–22%) |
| 4 | 100% (100–100%) | 32% (30–33%) | 32% (30–33%) |
| 5 | 100% (100–100%) | 42% (41–44%) | 42% (41–44%) |
| 6 | 97% (95–99%) | 53% (51–55%) | 50% (47–53%) |
| 7 | 95% (93–98%) | 62% (60–64%) | 57% (54–60%) |
| 8 | 93% (90–96%) | 68% (66–70%) | 61% (58–64%) |
| **9** | **87% (83–91%)** | **74% (72–76%)** | **61% (57–65%)** |
| 10 | 79% (75–84%) | 79% (78–81%) | 58% (54–63%) |
| 11 | 72% (67–76%) | 84% (83–86%) | 56% (51–61%) |
| 12 | 66% (61–70%) | 88% (86–89%) | 53% (49–58%) |
| 13 | 57% (53–62%) | 91% (90–92%) | 48% (43–53%) |
| 14 | 50% (45–55%) | 93% (92–94%) | 43% (39–48%) |
| 15 | 43% (39–48%) | 95% (94–96%) | 38% (33–43%) |
| 16 | 36% (32–41%) | 97% (96–97%) | 33% (28–38%) |
| 17 | 30% (26–35%) | 98% (97–98%) | 28% (24–32%) |
| 18 | 24% (20–28%) | 98% (98–99%) | 22% (18–27%) |
| 19 | 20% (16–23%) | 99% (99–99%) | 19% (15–22%) |
| 20 | 15% (12–18%) | 99% (99–100%) | 14% (11–17%) |
| 21 | 12% (9–15%) | 100% (100–100%) | 12% (9–14%) |
| 22 | 10% (8–13%) | 100% (100–100%) | 10% (7–12%) |
| 23 | 8% (6–10%) | 100% (100–100%) | 8% (6–10%) |
| 24 | 6% (4–7%) | 100% (100–100%) | 6% (4–7%) |
| 25 | 4% (2–5%) | 100% (100–100%) | 4% (2–5%) |
| 26 | 2% (1–3%) | 100% (100–100%) | 2% (1–3%) |
| 27 | 1% (1–2%) | 100% (100–100%) | 1% (1–2%) |
| 28 | 1% (0–1%) | 100% (100–100%) | 1% (0–1%) |
| 29 | 1% (0–1%) | 100% (100–100%) | 1% (0–1%) |
| 30 | 1% (0–1%) | 100% (100–100%) | 1% (0–1%) |
| 31 | 0% (0–0%) | 100% (100–100%) | 0% (0–0%) |
| 33 | 0% (0–0%) | 100% (100–100%) | 0% (0–0%) |

Abbreviations: ICP=intracranial pressure, ICU=intensive care unit, TIL=Therapy Intensity Level scale,[8,9] $TIL^{(Basic)}$=condensed TIL scale.[8] The numeric definition of TIL is listed in Table 1, and the calculation of daily (e.g., $TIL_{24}$) scores is described in the Methods. The 95% confidence intervals of performance metrics were derived from bootstrapping with 1,000 resamples of unique patients over 100 missing value imputations. If a decompressive craniectomy was performed as a last resort for refractory intracranial hypertension, each of the days following the operation were also considered days of surgical ICP control. The row in bold designates the threshold which maximises Youden’s J statistic.
